# Exploring novel risk loci for heart failure and the shared genetic etiology with blood lipids, blood pressure, and blood glucose: a large-scale multi-trait association analysis

**DOI:** 10.1101/2023.12.20.23300280

**Authors:** Yanchen Zhu, Yahui Wang, Zhaorui Cui, Fani Liu, Jiqiang Hu

**Affiliations:** Cardiology Department, Dongfang Hospital Beijing University of Chinese Medicine, Beijing, China

**Keywords:** Heart failure, Blood lipids, Blood pressure, Blood glucose, Shared genetics, Multi-trait association analysis

## Abstract

**Background:** The comprehensive exploration of genomic risk loci for heart failure (HF) remains constrained, and the genetic role of blood lipids (BL), blood pressure (BP) and blood glucose (BG) in HF has not been fully characterized.

**Methods:** We first assessed the global and local genetic correlations between HF and the quantitative traits of BL, BP, and BG. We then employed multi-trait association analysis and multi-trait colocalization analysis to identify novel and pleiotropic genomic risk loci for HF. Furthermore, we explored potential genes, pathways, tissues, and cells associated with HF involving BL, BP, and BG. Lastly, we investigated potential therapeutic targets for HF.

**Findings:** We found extensive global and local genetic correlations between HF and the traits of BL, BP, and BG. Multi-trait association analysis successfully identified 154 novel genomic risk loci for HF. Multi-trait colocalization analysis further revealed 46, 35, and 14 co-localized loci shared by HF with BL, BP, and BG, respectively. We found that the loci shared by HF with these traits rarely overlapped, indicating distinct shared mechanisms. Gene-mapping, gene-based, and transcriptome-wide association analyses prioritized noteworthy candidate genes (such as LPL, GRK5, and TNNC1) for HF. In enrichment analysis, HF exhibited comparable characteristics with cardiovascular traits and metabolic correlated to BL, BP, and BG. We provided genetic evidence for putative drugs, and highlighted 33 robust potential protein targets.

**Interpretation:** These findings will provide biological insights into the pathogenesis for HF, and benefit the development of preventive or therapeutic drugs for HF.

## Introduction

Heart failure (HF) is a prominent public health issue. Global statistics indicate that HF impacts over 64 million individuals, which contributes the major cause of cardiovascular hospitalization rates, mortality, and healthcare expenditures(1, 2). As the terminal state of cardiac diseases, HF is a polygenic disease and presents a more intricate genetic structure than other cardiometabolic disorders. Therefore, investigating the underlying genetic mechanisms of HF could enhance our etiological understanding of HF and facilitate the development of potential targets for intervention. However, the exploration of the genetic mechanisms of HF is still insufficient.

Previous studies have identified several genomic risk loci associated with HF(3, 4). However, further identification of risk loci for HF remains challenging due to the limited sample size. Therefore, it is necessary to employ advanced statistical genetics methods to investigate the association of potential loci with HF. Multi-trait joint analysis can borrow relevant information from multiple related traits and has become an effective statistical method to improve statistical power to identify novel genomic risk loci for target traits(5). In the clinical practice, dyslipidemia, elevated blood pressure (BP), and blood glucose (BG), are main risk factors for HF, and these abnormal statuses are highly prevalent among HF patients, exerting a substantial influence on disease progression(6).

Despite HF is an irreversible progressive disease, the abnormalities in blood lipids (BL), BP, and BG can be identified early and treated effectively with medications. Previous studies have reported that BL, BP, and BG have potential shared genomic loci with HF(7, 8). In addition, the present genetics association study of BL, BP, and BG exhibits a substantial sample size, demonstrating significant polygenic heritability and greater multitude of genomic loci in contrast to prior research. Therefore, their multi-trait joint analysis with HF can not only borrow information from these traits to deeply explore HF-related genetic variations, but also identify pleiotropic loci shared by HF with BL, BP, and BG.

In this study, we utilized the largest publicly available genome-wide association study (GWAS) summary statistics to perform multi-trait analysis between HF and the quantitative traits of BL, BP, and BG. Our study aims to achieve two primary objectives. Firstly, we aim to discover potential novel genomic risk loci for HF, elucidate its genetic mechanism, and explore potential drug targets. Secondly, we aim to investigate the shared genetic etiology basis for HF with BL, BP, and BG, and characterize the genetic roles of BL, BP, and BG in HF.

## Methods

### 1. Population samples and ethics

Figure 1 presents a schematic overview of our study. In this study, we utilized GWAS summary statistics for a total of 12 traits. For each trait, we utilized the most recent and largest publicly available GWAS summary statistics from European ancestry individuals. The GWAS summary statistics for HF were obtained from the Heart Failure Molecular Epidemiology for Therapeutic Targets (HERMES) consortium, which combined data from 26 cohort-level GWAS comprising 47,309 cases and 930,014 controls(3). The Global Lipids Genetics Consortium (GLGC) consortium provided summary statistics [high-density lipoprotein cholesterol (HDL-C), low-density lipoprotein cholesterol (LDL-C), total cholesterol (TC), and triglyceride (TG)] (N= 1,320,016) of BL(9), and additional summary statistics [apolipoprotein A1(APOA1) (N= 393,193) and apolipoprotein B(APOB) (N= 439,214)] for BL traits were from UK Biobank(10). The GWAS summary statistics for BP traits [systolic blood pressure (SBP), diastolic blood pressure (DBP), and pulse pressure (PP)] were used from a meta-analysis study of UK Biobank (N= 458,577) and the International Consortium of Blood Pressure genetics (ICBP) (N= 299,024, across 77 cohorts)(11). For BG, the summary statistics [fasting glucose (FG) (N=314,916) and hemoglobin A1c (HbA1c) (N=344,182)] were obtained from the UK Biobank-Neale lab(12). The supplementary Table 1 provides access to the specific details of each GWAS summary statistics.

**Fig. 1.**
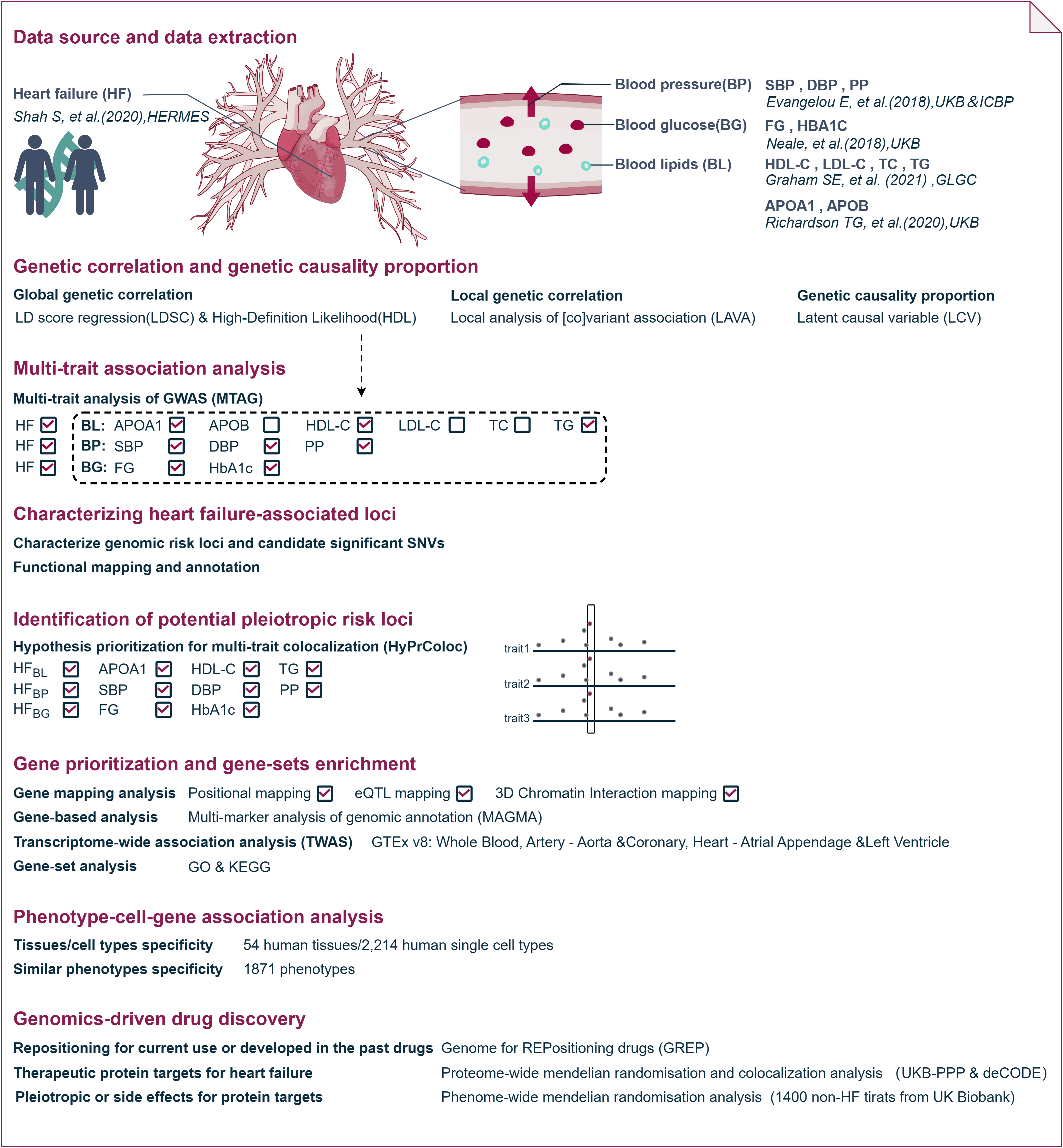
Design schematic of the present study. HDL-C: high-density lipoprotein cholesterol, LDL-C: low-density lipoprotein cholesterol, TC: total cholesterol, TG: triglyceride, APOA1: apolipoprotein A1, APOB: apolipoprotein B, SBP: systolic blood pressure, DBP: diastolic blood pressure, PP: Pulse pressure, FG: Fasting glucose, HbA1c: Hemoglobin A1c.

### 2. Statistical analysis

The GWAS summary statistics underwent genotypic quality control measures. Supplement Methods provide more comprehensive description of these analytical procedures.

Both linkage disequilibrium score regression (LDSC)(13) and high-Definition Likelihood (HDL)(14) were conducted to evaluate heritability of each trait and the genetic correlation between HF and the quantitative traits of BL, BP, and BG. Latent causal variable model (LCV)(15) was employed to further evaluate the genetic causality proportion between HF and these traits. Given the intricate genetic structure of each region, we employed local analysis of [co]variant association (LAVA)(16) to examine local genetic correlation. To correct for multiple testing for above statistical analyses, we applied the Benjamini-Hochberg false discovery rate (FDR) approach, with the threshold set at 0.05.

We utilized multi-trait analysis of GWAS (MTAG)(5) to identify novel genomic risk loci for HF by leveraging the interrelationships among pairwise traits exhibiting significant genetic correlation. To assess the overall inflation resulting from the violation of the homogeneous assumption in MTAG analysis, we computed the maximum FDR (maxFDR), which evaluate the overall inflation due to violation of the homogeneous assumption. The genome-wide significance threshold for HF_MTAG_ was determined to be P < 5×10^−8^. We further employed the Functional Mapping and Annotation of Genetic Associations (FUMA)(17) to characterize significant genomic loci. FUMA identified independent significant single nucleotide variants (SNVs) with a genome-wide significant and a linkage disequilibrium (LD) measure of r^2^□<□0.6. Lead SNVs were determined by selecting independent significant SNVs that were not in LD with each other at r^2^□<□0.1. Risk loci were defined by combining lead SNVs that physically overlapped or had LD blocks within 250□kb apart. FUMA additionally provided functional annotations such as ANNOVAR analysis, combined annotation dependent depletion (CADD) scores, and RegulomeDB scores. Variants with a CADD score exceeding 12.37 were deemed potentially deleterious. To identify shared causal variants within each genomic locus across traits, hypothesis prioritization in multi-trait colocalization (HyPrColoc)(18) analysis was conducted based on genomic risk loci of HF_MTAG_. The colocalized locus was considered if the posterior probability exceeded 0.7.

Based on MTAG results, we conducted a comprehensive investigation into the underlying shared biological mechanisms for HF with BL, BP, and BG. In order to identify potential genes associated with HF, a combination of gene-mapping, multi-marker analysis of genomic annotation (MAGMA)(19), and transcriptome-wide association analysis (TWAS)(20) methods were employed. The threshold for FDR.P correcting multiple testing was set at 0.05. Furthermore, we elucidated the biological pathways by conducting gen-set enrichment analyses using the GO and KEGG databases. In addition, we used the DESE (driver tissue estimation by selective expression) approach implemented in phenotype-cell-gene association analysis (PCGA)(21–24) website to further explore the tissue/cell types specificity and similar phenotypes for HF_MTAG_.

### 3. Genomics-driven drug discovery

To further enhance gene-driven drug discovery for HF, we utilized the Genome for Repositioning drugs (GREP)(25) software to determine the clinical indication categories and the enrichment of candidate effector genes in drug repositioning. GREP conducts Fisher’s exact tests to detect enrichment of a gene set within genes targeted by drugs currently in use or previously developed for the specific clinical indication category (Anatomical Therapeutic Chemical Classification System [ATC]). Furthermore, proteome-wide mendelian randomization (MR) was utilized to identify potential therapeutic plasma protein targets for HF [UK Biobank Pharma Proteomics Project (UKB-PPP)(26), and deCODE genetics(27)]. The primary analysis was employed using the inverse variance weighted method, and all results underwent FDR correction (FDR.P<0.05). The steiger test was conducted to assess whether the MR results were affected by potential reverse causality. The MR-Egger regression intercept test and Cochran’s Q test were used to respectively assess the presence of pleiotropy and heterogeneity of the results. Bayesian colocalization analysis was utilized to examine whether the observed significant protein-disease pairs were driven by single causal SNV in the LD region(28). Specifically, we set the threshold of posterior probability hypotheses 4(the protein and HF were driven by single causal SNV) at 0.7. Based on MR and colocalization, phenome-wide MR analysis was conducted to investigate the pleiotropic or side effects of the protein targets (FDR.P<0.05).

## Results

### 1. Genetic correlation and genetic causality proportion

LDSC results showed that BG (FG, HbA1c), BP (SBP, DBP, and PP), and TC exhibited significant positive genetic correlations with HF. Conversely, HDL-C and APOA1 displayed significant negative genetic correlations (Supplementary Table 2; Fig. 2). The results pertaining to HDL exhibit a notable level of congruity with the findings of LDSC (Supplementary Table 3; Fig. 2). LAVA discovered a total of 366 significant bivariate local genetic correlations (FDR.P<0.05) for HF with BL, BP, and BG at 224 specific regions (Supplementary Table 4; Fig. 2). The present study reveals a general pattern in the local genetic correlations for HF with HDL-C, APOA1, and APOB, indicating a notable inclination towards negative correlations. Conversely, other traits exhibit a tendency towards positive correlations, aligning with the direction of their respective global genetic correlations. It should be noted that the coexistence of both negative and positive local genetic correlations suggests a multifaceted impact.

**Fig. 2.**
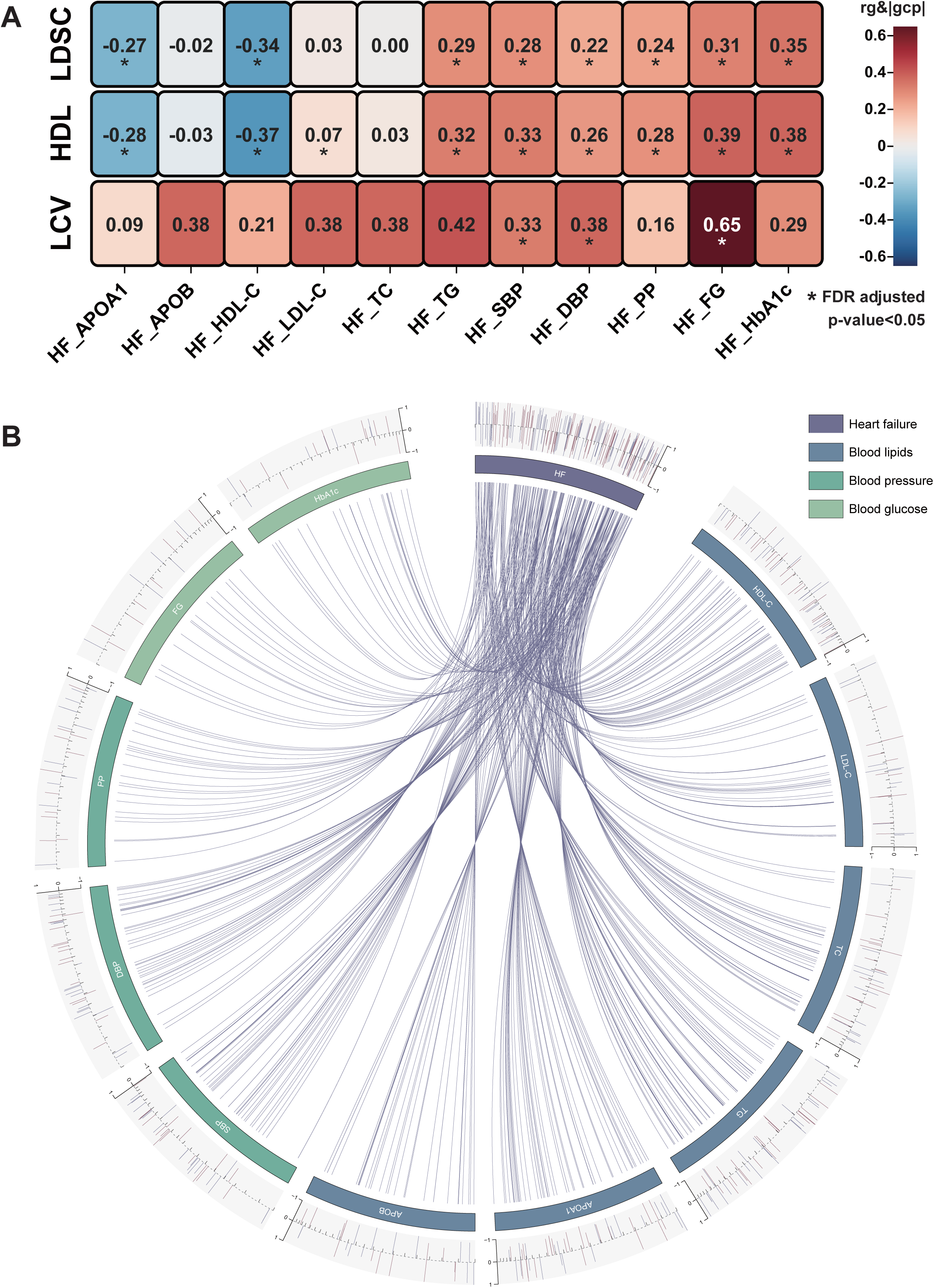
Genetic correlation and genetic causality proportion between HF and the quantitative traits of BL, BP, and BG. **a**, The correlation heat-map shows the global genetic correlation and genetic causality proportion between HF and the quantitative traits of BL, BP, and BG. b, Chord diagram shows the local genetic correlation between HF and these traits. *LDSC: linkage disequilibrium score regression, HDL: high-Definition Likelihood, LCV: latent causal variable model, rg: genetic correlation estimate, gcp: genetic causality proportion, HF: heart failure, HDL-C: high-density lipoprotein cholesterol, LDL-C: low-density lipoprotein cholesterol, TC: total cholesterol, TG: triglyceride, APOA1: apolipoprotein A1, APOB: apolipoprotein B, SBP: systolic blood pressure, DBP: diastolic blood pressure, PP: Pulse pressure, FG: Fasting glucose, HbA1c: Hemoglobin A1c*.

The LCV analyses demonstrated significant associations between HF and FG (|GCP| = 0.65, FDR.P = 0.015), SBP (|GCP| = 0.33, FDR.P = 0.002), DBP (|GCP| = 0.38, FDR.P = 7.91×10^−5^) (Supplementary Table 5; Fig. 2), suggesting a potential genetic causal association for HF with FG, SBP, and DBP.

### 2. Multi-trait association analysis

Multi-trait association analyses combining HF with BL, BP, and BG, respectively, significantly increased the effective sample sizes and heritability of HF. Specifically, the polygenic heritability exhibited an impressive 2.02-, 1.83-, and 1.3-fold escalation for each respective HF_MTAG_ consideration. Accompanied by max-FDR values of 0.046, 0.032, and 0.029 (Table 1), respectively, showed no indication of inflation, thereby confirming the reliability of our methodology and findings. We successfully identified 93, 48 and 40 genomic risk loci from 486, 109 and 97 independent genome-wide significant SNVs for HF_BL_, HF_BP_, and HF_BG_, of which 85, 41, and 31 are novel risk loci for HF (Supplementary Table 6-12; Fig. 3). The HyPrColoc further identified 46, 35, and 14 pleiotropic loci that exhibited evidence of colocalization, indicating the widely distribution of shared genominc risk loci for HF with BL, BP, and BG (Supplementary Table 13-15).

**Fig. 3.**
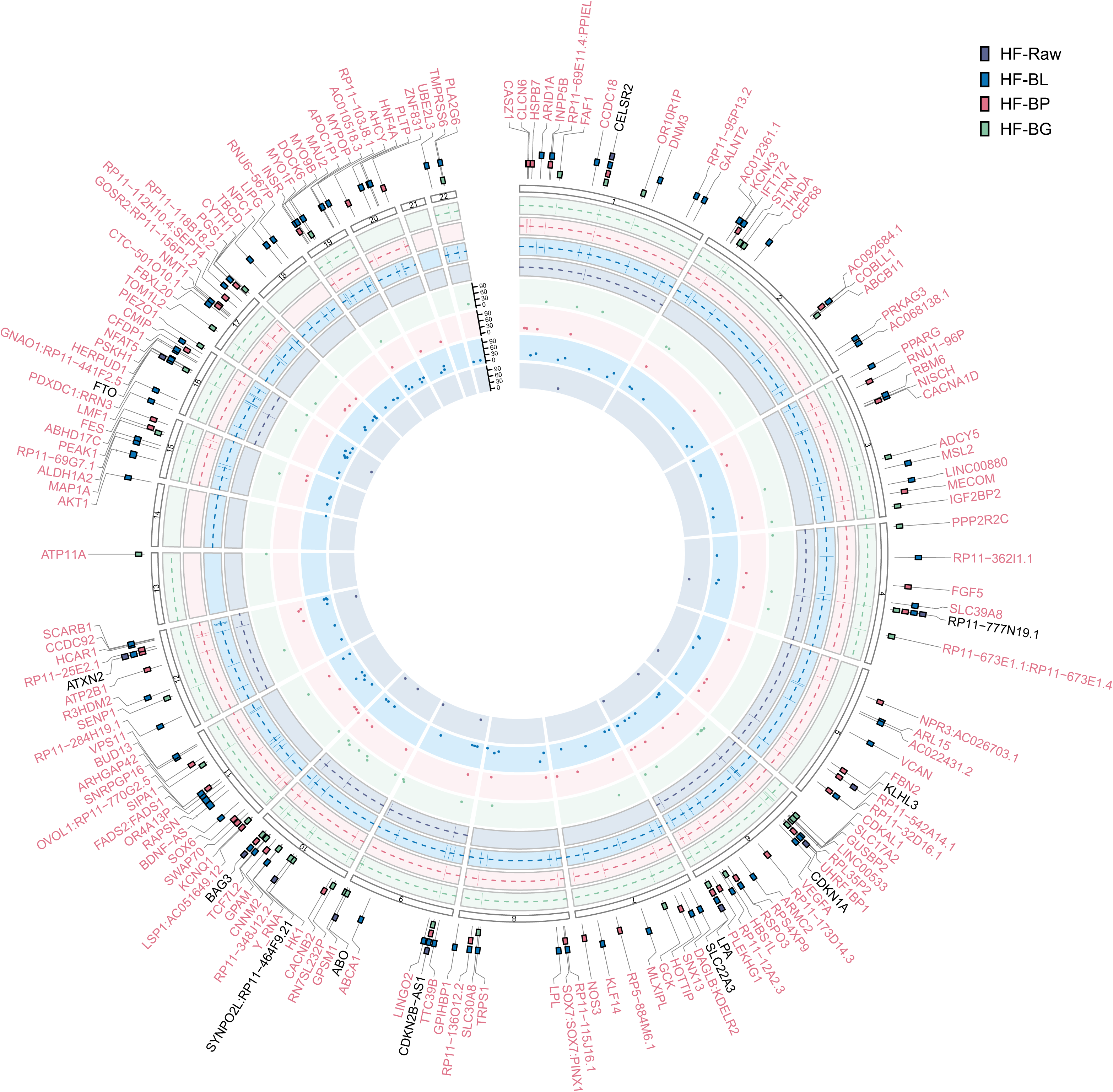
Circular plot of multi-trait analysis of GWAS results for HF_GWAS_, HF_BL_, HF_BP_, and HF_BG_. The dots in the inner circle represent the loci (index SNV) associated with each trait. The middle part presents the genomic risk loci. The outer part presents the nearest genes to the top signals of HF, and the associated traits are indicated by different coloured squares. Genes in red denote novel loci. The details of these risk loci in the traits of BL, BP, and BG are presented in Supplementary Table 10-12. *HF: heart failure, BL: blood lipids, BP: blood pressure, BG: blood glucose*.

**Table 1.** The summary of MTAG results for heart failure. MTAG: multi-trait analysis of GWAS; h2: Observed scale heritability (LD score regression); Se: standard error; gcov_int: genetic covariance intercept; λ*GC: the genomic control inflation factor based on the median; maxFDR: maximum FDR; CADD: combined annotation dependent depletion*.

| Trait | liability-scale h2 (Se) | $\lambda_{GC}$ | Mean Chi^2 | gcov_int (Se) | maxFDR_MTAG | N_sample size | N_risk loci | N_novel risk loci | N_genome-wide significant SNVs | N_exons | N_CADD score>12.37 |
| --- | --- | --- | --- | --- | --- | --- | --- | --- | --- | --- | --- |
| HFGWAS | 0.0304 (0.0024) | 1.127 | 1.159 | 1.006 (0.007) | NA | 964057 | 11 | - | 736 | 20 | 40 |
| HFBL | 0.0618 (0.0032) | 1.114 | 1.259 | 0.826 (0.010) | 0.046 | 1366841 | 93 | 85 | 4188 | 117 | 186 |
| HFBP | 0.0558 (0.0026) | 1.118 | 1.227 | 0.862 (0.009) | 0.032 | 1243259 | 48 | 41 | 1930 | 41 | 78 |
| HFBG | 0.0395 (0.0022) | 1.127 | 1.189 | 0.933 (0.008) | 0.029 | 1240601 | 40 | 31 | 934 | 13 | 28 |

Furthermore, ANNOVAR offered notable variants’ annotation information regarding these loci (Supplementary Table 16-18). For instance, we discovered 117 (2.8%) of 4188 genome-wide significant SNVs from HF_BL_ loci are exons, 77 of which are located in protein-coding genes (35 synonymous SNVs, 42 nonsynonymous SNVs). Further annotation by Combined Annotation-Dependent Depletion (CADD) scores predicted that 186 SNVs were deleterious (CADD score > 12.37). We noted several novel genomic risk loci and variants of interest. The most significant risk locus is located at region 8p21.3, and its index variant rs144958026 (P = 5.51×10^-65^) is an intronic variant of LPL gene which encoding lipoprotein lipase. The next significant locus is located at region 11q12.2, whose index variant rs174551 (P = 6.92×10^-50^) is located in the 5-UTR of genes FADS1 and FADS2. Fatty acid desaturase (FADS) genes encode the fatty acid desaturase enzyme that is mainly responsible for regulating the biosynthesis of unsaturated fatty acids in the fatty acid metabolic pathway. In addition, rs34312154 (P=3.38×10^-09^) in region 11p11.2 has the highest CADD score of 31. We also noted four index variants that fell in the exonic region, including rs61749613 (P = 3.62×10^-9^; gene = VCAN), rs55707100 (P = 2.61×10^-15^; gene = MAP1A), rs1800961 (P = 4.06×10^-16^; gene = HNF4A), and rs9935936 (P = 1.72×10^-9^; gene = GNAO1:RP11-441F2.5).

Notably, by combining MTAG analysis for HF with BL, BP and BG, we discovered a total of 165 HF risk loci, of which 154 of these identified loci were novel discoveries in relation to HF. By comparing locus reported in GWAS catalog (Supplementary Table 19-21), we found that a total of 143 loci have not been reported before. Interestingly, there was only a 6.1% overlap between the loci related to HF_BL_, HF_BP_, and HF_BG_, indicating that these three with different mechanisms that contribute to the effects on HF.

### 3. Gene-based and transcriptome-wide association analysis

FUMA mapped 1466 [including 1251 positional mapped genes, 327 expression quantitative trait loci (eQTL) mapped genes and 540 3D Chromatin Interaction mapped genes], 486 (including 406 positional mapped genes, 108 eQTL mapped genes and 145 3D Chromatin Interaction mapped genes), 500 (including 435 positional mapped genes, 113 eQTL mapped genes and 190 3D Chromatin Interaction mapped genes) genes for HF_BL_, HF_BP_, and HF_BG_, respectively (Supplementary Table 22-24). We obtained a total of 2444 candidate genes, of which only 5.8% overlapped.

The MAGMA gene-based analysis found a total of 910, 703, and 398 genes significantly associated with HF_BL_, HF_BP_, and HF_BG_ (FDR.P<0.05) (Supplementary Table 25-27). We obtained a total of 1539 candidate genes, of which only 23.8% overlapped.

Combined with tissue-specific eQTL data (GTEx v8)(29) for whole blood, artery-aorta, coronary, heart-atrial appendage, and left ventricle, TWAS analysis revealed 248, 71, 53 genes for HF_BL_, HF_BP_, and HF_BG_ (FDR.P<0.05) (Supplementary Table 28-30). Among a total of 359 candidate genes, only a minimal proportion of 3.34% exhibited overlapping associations.

In summary, the results obtained from the parallel gene-level analyses are consistent with the findings of the preceding SNV-level analyses, indicating a limited overlap among the genes associated with HF_BL_, HF_BP_, and HF_BG_ (Supplementary Fig. 1). This provides additional support to the concept that BL, BP, and BG exert their impact on HF through separate mechanisms.

### 4. Gene-set enrichment

The MAGMA gene-set analysis indicated that the predominant biological processes of 87 pathways implicated in the interaction between HF and BL primarily revolve around lipid metabolism. Furthermore, we identified 43 HF_BP_ and 16 HF_BG_ pathways (Supplementary Table 31-34; Supplementary Fig. 2). The results demonstrate that the biological processes involved in the association between HF and BP are significantly linked to vasculature and muscle development, while the association between HF and BG primarily revolves around hexokinase activity and lipid metabolism.

### 5. Phenotype-cell-gene specificity association analysis

We found that arterial, adipose, and lung tissues displayed significant associations with HF_MTAG_. The tissues that exhibited the top 3 strongest correlations with HF_BL_ were adipose-visceral omentum, artery-coronary, and adipose-subcutaneous (Supplementary Table 34-36; Supplementary Fig. 3). For HF_BP_, the top 3 significant associations were observed in artery-coronary, artery-tibial, and lung. And HF_BG_ demonstrated a strong correlation with artery-coronary, subcutaneous adipose, and artery-tibial. Among 2,214 human single cell types, macrophages were identified as pivotal cell types for HF_BL_, endothelial cells for HF_BP_, and ACE2-expressing AT2 cells exhibited the most notable association with HF_BG_ (Supplementary Table 37-39; Supplementary Fig.4). Through the examination of phenotypic similarities in 1,588 unique phenotypes, our findings align with established biological knowledge. As expected, HF_BL_, HF_BP_, and HF_BG_ exhibited comparable characteristics with cardiovascular traits and metabolic traits correlated to BL, BP, and BG (Supplementary Table 40-42; Supplementary Fig. 5).

### 6. Existing drug effects for heart failure

Using GREP software for enrichment of significant HF-related genes obtained from gene-mapping, MAGMA, and TWAS analyses, we identified significant enrichment genes (ABCA1, APOB, LPL, PPARD, PPARG, and THRA) for HF_BL_ in drug targets of the cardiovascular lipid-modifying drug targets (ATC C10 drugs), which encode targets for probucol, mipomersen, clofibrate, gemfibrozil, bezafibrate, and dextrothyroxine (Supplementary Table 43). HF_BP_ genes demonstrated significant enrichment within the cardiovascular beta-blocking drug targets (ATC C07 drugs), incorporating ADRA1B and KCNH2 genes that encode targets for labetalol and sotalol (Supplementary Table 43). And HF_BG_ genes exhibited a significant enrichment within drug targets for systemically used antivirals (ATC J05 drugs), which included the ADORA2B and CES1 genes, the encoding targets for vidarabine and oseltamivir (Supplementary Table 43).

### 7. Plasma protein targets for heart failure

We performed two-sample MR using plasma protein quantitative trait loci (pQTL) statistics from the UK Biobank Pharma Proteomics Project (UKB-PPP)(26) and deCODE(27) genetics with HF_MTAG_ statistics to evaluate protein targets for primary prevention for HF. We identified a total of 33 protein targets that satisfied the statistical significance of both MR (FDR.P<0.05) and colocalization (PP.H4>0.7) support in at least one cohort, and there was no reverse causal association by steiger testing (Fig. 4). Sensitivity analyses also showed that there was no pleiotropy or obvious heterogeneity. Among the 4907 pQTL for drug target proteins from deCODE, our results identified a total of 16 plasma proteins with potential causal associations with HF risk (Supplementary Table 44-46). These proteins were supported by MR (FDR.P < 0.05) and colocalization (PP.H4>0.7) analyses, and no reverse causal association was indicated by steiger test. Among them, GSTM4, NADK, and NPPB were replicated in the UKB-PPP cohort (FDR.P < 0.05; PP.H4>0.7; Steiger.P < 0.05) and had consistent effect directions. GSTA1 and LILRA5 were supported by colocalization in the UKB-PPP cohort (PP.H4>0.7), and the MR results showed nominal significance (P<0.05). INHBC, despite having a consistent direction of effect in the UKB-PPP cohort, was not supported by MR and colocalization.

**Fig. 4.**
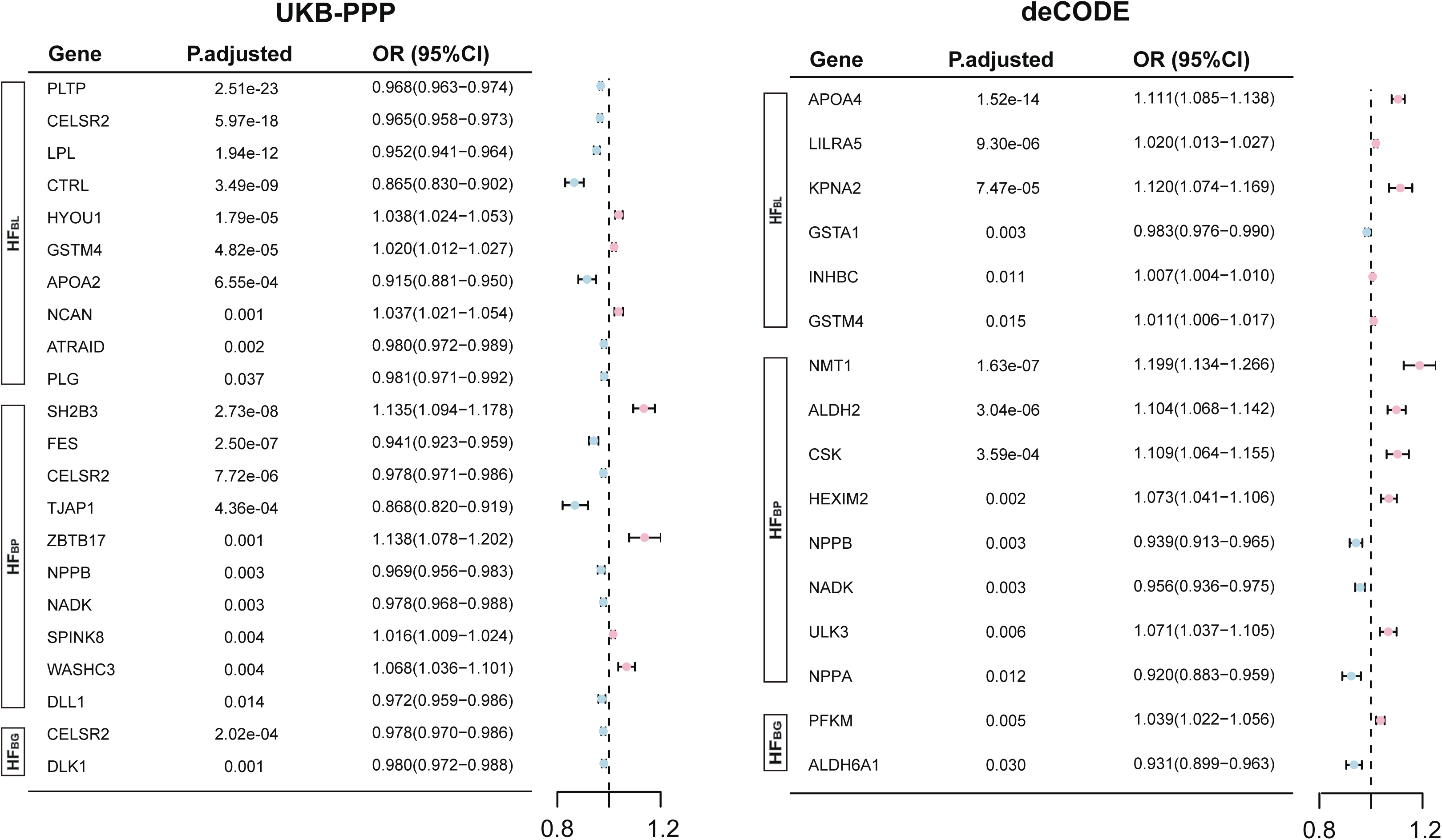
Forest plot of protein targets for heart failure indicated by mendelian randomization analysis. The forest plot only displayed 33 specific protein targets that satisfied the statistical significance of both mendelian randomization (FDR.P<0.05) and colocalization (PP.H4>0.7) in at least one cohort. The square symbolizes the odds ratio for mendelian randomization, while the horizontal line represents the 95% confidence interval. *UKB-PPP: UK Biobank Pharma Proteomics Project; deCODE: deCODE genetics; HF: heart failure; BL: blood lipids; BP: blood pressure; BG: blood glucose; P.adjusted: false discovery rate (FDR) adjusted p-value; OR: odds ratio; CI: confidence interval*.

The remaining proteins (APOA4, KPNA2, ALDH2, CSK, HEXIM2, NMT1, NPPA, ULK3, ALDH6A1, PFKM) are only available in the deCODE cohort due to strict instrumental variable filtering conditions. For 2941 pQTL in UKB-PPP, additional 17 protein targets (FDR.P < 0.05; PP.H4>0.7; Steiger.P < 0.05) were identified (Supplementary Table 47-49). Among them, NCAN, PLTP, and DLL1 received colocalization support in the previous deCODE cohort (PP.H4>0.7), and the MR results all showed nominal significance (P<0.05). PLG did not find MR significance in the deCODE cohort but was supported by colocalization. DLK1 was not supported by MR and colocalization. The remaining proteins (APOA2, ATRAAID, CELSR2, CTRL, HYOU1, LPL, CELSR2, FES, SH2B3, SPINK8, TJAP1, WASHC3, ZBTB17) could not be replicated in the deCODE cohort due to lack of instrumental variables. In addition, further phenome-wide MR analysis revealed that no side effects for the majority of identified protein targets were observed, while simultaneously indicating potential therapeutic benefits for other significant medical conditions (Supplementary Results; Supplementary Fig. 6-7).

## Discussion

Multi-trait association analysis for HF with BL, BP, and BG significantly improved the statistical power in identification of novel genomic risk loci for HF. We discovered 154 novel HF loci, of which 143 have not been reported previously. We further explored the shared genetic etiology, including potential genes, pathways, tissues, and cells for HF with these traits. The novel loci we have identified hold promise as potential targets for drug development or therapeutic interventions. Furthermore, bioinformatics analyses provide genetic evidence for putative drug effects and novel protein targets of HF (Supplementary Discussion).

In this study, our findings suggest that there is minimal overlap between the risk loci or genes associated with HF_BL_, HF_BP_, and HF_BG_, indicating that the risk of HF caused by these factors appears to be independent, with distinct biological mechanisms. Consequently, the combination therapeutic strategy targeting HF with BL, BP, and BG may offer additive and diverse benefits.

The 154 risk loci include some noteworthy HF-related genes. Lipoprotein lipase (LPL)-mediated hydrolysis of circulating lipoproteins into fatty acids (FA) is thought to be the primary source of FA utilization by the heart(30). Studies have shown that mice with cardiac LPL deficiency develop HF as they age and are unable to respond normally to increased afterload. In addition, LPL-deficient mice result in a reduction in HDL-C of more than 50%(30). The significance of G protein-coupled receptor kinase 5 (GRK5) as a key regulator of pathological cardiac hypertrophy has been documented, rendering it a potential therapeutic target for HF. GRK5 was found to be upregulated in the myocardium of individuals with HF, as well as to promote maladaptive cardiac hypertrophy in animal models(31). Furthermore, an experimental and pharmacogenomic research showed the pharmacogenomic interaction between GRK5 and beta-blocker therapy, wherein the presence of the GRK5-Leu41 polymorphism was linked to a reduction in mortality rates among African American individuals afflicted with HF or cardiac ischemia(32). TNNC1 (Troponin C1, Slow Skeletal and Cardiac Type) is a protein coding gene. Diseases associated with TNNC1 include cardiomyopathy, familial hypertrophic and cardiomyopathy(33). TNNC1 is mainly related to striated muscle contraction. The inotropic drug levosimendan, which targets TNNC1, is a common drug for HF.

We acknowledge some limitations. Our analyzes were limited to individuals of European ancestry and the results may not be generalizable to other ancestry. Additionally, we excluded all rare variants (MAF < 1%) from the MTAG analysis and therefore from all subsequent analyses. Therefore, we may not be able to identify rare variants with large effects. In MR analyses, we exclusively utilized cis-regulatory regions as instrumental variables, which may mitigate horizontal pleiotropy to some extent and result in reduced statistical efficacy. Lastly, our inquiry was restricted to identifying and validating pQTL data utilizing accessible instrumental variables, potentially neglecting alternative therapeutic targets.

## Supporting information

Supplementary Results

Supplementary Discussion

## Data Availability

All data produced in the present study are available upon reasonable request to the corresponding author.

## Conclusion

In conclusion, this multi-trait association study provides important insights to the risk loci for HF and the shared genetic etiology for HF with BL, BP, and BG. Additionally, it highlights existing drugs and potential novel protein targets for HF therapy. These findings will provide biological insights into the pathogenesis for HF, and benefit the development of preventive or therapeutic drugs for HF.

## Contributors

YZ, Conceptualization, Data curation, Formal analysis, Methodology, Writing-original draft; YW Conceptualization, Methodology, Writing-original draft; ZC and FL, Conceptualization, Methodology, Writing-review, and editing; JH, Conceptualization, Data curation, Funding acquisition, Writing-review, and editing. All authors critically edited the manuscript, followed by reading and approving the final version. All authors had full access to all the data in the study and had final responsibility for the decision to submit for publication.

## Data sharing statement

The GWAS summary statistics for heart failure and blood pressure used in this study are deposited in the GWAS Catalog (https://www.ebi.ac.uk/gwas/) and the accession codes are as follows: HF (GCST009541),SBP (GCST006624), DBP (GCST006630), and PP (GCST006629).Summary-level data from Global Lipids Genetics Consortium(GLGC) for blood lipids are available as follows: https://csg.sph.umich.edu/willer/public/glgc-lipids2021, and additional lipids traits ( APOA1 [ieu-b-107] and APOB [ieu-b-108]) are available through the IEU Open GWAS database. Data for blood glucose can be obtained from UK Biobank-Neale lab via http://www.nealelab.is/uk-biobank. The pQTL summary statistics are acquired from the UK Biobank Pharma Proteomics Project (UKB-PPP) through the website https://www.synapse.org, and from deCODE genetics via http://www.decode.com. Phenome-wide summary statistics for the UK Biobank can be accessed at https://www.leelabsg.org/resources.

## Code availability

No novel custom computer code or mathematical algorithm was employed in generating the results pivotal to the derived conclusions.

## Acknowledgements

We thank Figdraw (www.figdraw.com) for its help in creating the figures. We would like to express our gratitude to all researchers and participants involved in GWAS studies, for their generous sharing their data.

## Supplementary Materials

**Supplementary Methods**

**Supplementary Results**

**Supplementary Discussion**

**Supplementary Tables 1-49**: Table of contents, S1-S49.

**Supplementary Fig. 1 Wayne diagrams of the results indicated by gene base and transcriptome-wide association analyses.** a, Plots shows the results of gene-mapping analysis. b, Plots shows the results of MAGMA and TWAS analysis. *MAGMA: multi-marker analysis of genomic annotation, TWAS: transcriptome-wide association analysis*.

**Supplementary Fig. 2 Significantly enriched in GO and EKGG gene sets of heart failure indicated by MAGMA analysis.** a, gene-set enrichment of HF_BL_. b, gene-set enrichment of HF_BP_. c, gene-set enrichment of HF_BG_. *GO: Gene Ontology, KEGG: Kyoto Encyclopedia of Genes and Genomes, MAGMA: multi-marker analysis of genomic annotation, HF: heart failure, BL: blood lipids, BP: blood pressure, BG: blood glucose*.

**Supplementary Fig. 3 Enrichment of tissue specificity associated with heart failure.** a, associated tissues of HF_BL_. b, associated tissues of HF_BP_. c, associated tissues of HF_BG_. *HF: heart failure, BL: blood lipids, BP: blood pressure, BG: blood glucose*.

**Supplementary Fig. 4 Enrichment of cell-type specificity associated with heart failure.** a, associated cell types of HF_BL_. b, associated cell types of HF_BP_. c, associated cell types of HF_BG_. *HF: heart failure, BL: blood lipids, BP: blood pressure, BG: blood glucose*.

**Supplementary Fig. 5 Enrichment of similar phenotype specificity associated with heart failure.** a, associated similar phenotypes of HF_BL_. b, associated similar phenotypes of HF_BP_. c, associated similar phenotypes of HF_BG_. *HF: heart failure, BL: blood lipids, BP: blood pressure, BG: blood glucose*.

**Supplementary Fig. 6 The exploration of non-HF pleiotropic or side effects for the protein targets from UKB-PPP.** Manhattan plot utilizes black circles to highlight the statistically significant results of the FDR correction. *UKB-PPP: UK Biobank Pharma Proteomics Project*.

**Supplementary Fig. 7 The exploration of non-HF pleiotropic or side effects for the protein targets from deCODE.** Manhattan plot utilizes black circles to highlight the statistically significant results of the FDR correction. *deCODE: deCODE genetics*.

