## Supplementary Methods for "Exploring novel risk loci for heart failure and the shared genetic etiology with blood lipids, blood pressure, and blood glucose: a large-scale multi-trait association analysis"

**1.Genotypic quality control**

The GWAS summary statistics were underwent genotypic quality control measures by, (i) excluding insertions or deletions polymorphisms, (ii) removing rare variants with the minor allele frequency (MAF) less than 0.01, (iii) excluding palindromic single nucleotide polymorphisms (SNVs), (iv) removing SNVs within the major histocompatibility complex region due to its complex linkage disequilibrium structure, (v) restricting our summary statistics to 1000 Genomes Project Phase 3 European reference SNVs to ensure consistency(1). In subsequent analyses, corresponding data processing programs were added to adapt for the requirements of different analytical methods.

**2.Global genetic correlation analysis**

We conducted both linkage disequilibrium score regression (LDSC)(2) and high-Definition Likelihood (HDL)(3) to evaluate heritability of each trait and the genetic correlation between HF and the quantitative traits of BL, BP, and BG. Following the requirements of the LDSC approach, we used pre-computed LD scores from European reference panel in 1000 Genomes Project Phase 3 and retained only about 1.2 million well-imputed HapMap3 SNVs in LDSC analysis. We did not restrict the intercept term because of unknown population stratification or sample overlap. The HDL approach improves precision of genetic correlation estimates compared to LDSC approach, which is due to its account of full LD information and the fact that it is a likelihood-based method instead of a moments-based method. We used a 1,029,876 QCed UK Biobank imputed HapMap3 SNVs reference panel that pre-developed based on HDL approach. Lastly, we used the FDR correction as multiple testing correction for the results of above analyses.

**3.Local genetic correlation**

Considering the complex variability of genetic regions, global genetic correlations may underestimate pleiotropic correlations among traits. In particular, inconsistencies in the direction of genetic correlations in different regions may have antagonistic effects and lead to insignificant overall genetic correlations. Therefore we used local analysis of [co]variant association (LAVA) to assess local genetic correlation between HF and the quantitative traits of BL, BP, and BG(4). We used the genetic covariance intercept obtained from the LDSC analysis to correct for potential sample overlap. We tested local genetic correlation in 2495 genomic regions and applied the FDR method for multiple testing.

**4.Genetic causality proportion**

We employed the latent causal variable model (LCV) to evaluate the genetic causality proportion (GCP) between HF and these quantitative traits(5). The LCV approach defines a latent variable serves as a mediator for the genetic correlation between two traits. LCV aims to assess whether the latent variable exhibits a stronger association with one trait compared to the other. GCP values range from -1 to 1, as a negative value indicates complete genetic causality of trait 2 on trait 1 and vice versa. In the LCV analysis, the LD scores used in the analysis were pre-provided by the LDSC method. The statistical significance GCP were identified with FDR-corrected p-values below 0.05.

**5.Multi-trait association analysis**

The multi-trait analysis of GWAS (MTAG) analysis utilizes a generalized inverse-variance-weighted meta-analysis approach to identify novel genetic associations for each trait by leveraging the interrelationships among these traits. For summary statistics included, the MTAG analysis enables correction for potential sample overlap, which was calculated by LDSC approach. A key homogeneity assumption of the MTAG method is that all SNVs share the same variance-covariance matrix. However, the estimator of MTAG can be still consistent even if this assumption is violated. The max FDR (the FDR in the worst case) is thus used to evaluate the overall inflation due to violation of the homogeneous assumption. MTAG analysis produces trait-specific effect estimates for each SNV, from which novel summary statistics is generated for each trait.

Initially, we performed MTAG analysis for HF with all quantitative traits of BL, BP, and BG. However， the maxFDR was calculated at 0.215, which indicates the unreliability of the results. Consequently, subsequent MTAG analyses were conducted for HF with BL, BP, and BG respectively. And we recalculated the maxFDR to ensure that the analyses did not result in the overall inflation due to violation of the homogeneous assumption. In MTAG analysis, the genome-wide significant threshold was set at 5×10^−8^.

**6.Functional annotation**

We employed the Functional Mapping and Annotation of Genetic Associations (FUMA)(6) SNP2GENE function v1.6.1 to characterize significant genomic loci. FUMA identified independent significant SNV with a genome-wide significant (5 ×10^−8^) and a linkage disequilibrium (LD) measure of r^2^ < 0.6. Lead SNVs were determined by independent significant SNVs that were not in LD with each other at r^2^ < 0.1. Risk loci were defined by combining lead SNVs that physically overlapped or had LD blocks within 250 kb apart. FUMA additionally provided functional annotations including ANNOVAR (v2017-07-17) analysis, combined annotation dependent depletion (CADD) scores, and RegulomeDB scores. Variants with a CADD score exceeding 12.37 were deemed potentially deleterious.

**7.Multi-trait colocalization**

Hypothesis prioritization in multi-trait colocalization (HyPrColoc)(7) analysis was conducted to identify pleiotropic genomic risk loci and shared causal variants within the genomic locus across HF_MTAG_ and quantitative traits of BL_MTAG_, BP_MTAG_, and BG_MTAG_. The HyPrColoc is a Bayesian approach that can effectively colocalize multiple traits simultaneously. The HyPrColoc analysis was restricted to the region within ±500 kb from the top SNV of HF_MTAG_ risk loci in this study. We performed the HyPrColoc analysis with default settings, which included a prior probability of initial trait association of 1×10^−4^ and a conditional probability of subsequent traits having shared association of 0.02. The colocalized locus was considered if the posterior probability exceeded 0.7.

**8.Gene mapping analysis**

To map genome-wide significant SNVs from loci to specific genes, we performed the gene mapping procedure in FUMA v1.6.1(6). We used positional mapping (within ±250 kb from the locus), eQTL mapping (SNVs with FDR corrected eQTL P < 0.05 in whole bood or cardiovascular tissues) and 3D Chromatin Interaction Mapping (FDR.P < 10^−6^ in aorta and ventricles tissues) to identified candidate genes.

**9.Genome-wide gene-based and gene-set analyses**

Multi-marker analysis of genomic annotation (MAGMA) v1.08 analysis was performed by using FUMA(6). The default SNP-wise test in MAGMA were performed on 18,199 autosomal genes and the mean chi-square statistic were calculated for the variants annotated to each gene. The window for gene annotation was set to 5 kb based on NCBI build 37. In addition, we elucidated the biological pathways by conducting MAGMA gene-set enrichment analysis using the Gene Ontology (GO) and Kyoto Encyclopedia of Genes and Genomes (KEGG) databases. The p-value of the gene-based and gene-set analyses was corrected for multiple testing using FDR, and the statistical significance threshold was set at 0.05.

**10.Transcriptome-wide association analysis**

We conducted transcriptome-wide association studies (TWAS)(8) analysis using FUSION software to assess HF-related gene expression in the cardiovascular tissues. We performed TWAS analysis for HF_MTAG_ by using pre-computed predictive models from five tissue sources (cardiac-related whole blood, artery - aorta & coronary, heart - atrial appendage and left ventricle) in GTEx v.8(9). The results of TWAS were statistically significant as the FDR-corrected p-value was less than 0.05.

**11.Phenotype-cell-gene association analysis**

We used the DESE (driver tissue estimation by selective expression) approach implemented in phenotype-cell-gene association analysis (PCGA) website to further explore the associated tissue/cell types and genes of HF(10-13). DESE prioritizes disease-driving tissue/cell types by integrating GWAS summary statistics and gene expression profiles, with the underlying hypothesis that disease-associated genes tend to be selectively expressed in disease-driven tissues or cells. Combined with GWAS summary data for HF_MTAG_, we inspected the enrichment of genes related to HF in 54 tissues, 2214 human cell types and 1588 unique phenotypes.

**12.Genome for repositioning drugs**

We employed the Genome for Repositioning drugs (GREP)(14) software to ascertain clinical indication categories and the potential enrichment of candidate effector genes in drug repositioning. We utilized genes from gene mapping, as well as genes with the FDR-corrected p-value less than 0.05 in MAGMA and TWAS, to form the sets of disease risk genes. GREP conducts a sequence of Fisher's exact tests to detect enrichment of a gene set within genes targeted by drugs currently in use or previously developed from the specific clinical indication category (Anatomical Therapeutic Chemical Classification System [ATC]).

**13.Mendelian randomization and sensitivity analyses**

We performed two-sample mendelian randomization (MR) to identify novel potential causal plasma protein targets for HF. This study included two large-scale plasma protein quantitative trait loci (pQTL) summary statistics obtained from UK Biobank Pharma Proteomics Project (UKB-PPP)(15) and deCODE genetics(16). The latest published plasma pQTL statistics from the UKB-PPP contains 2,941 plasma proteins in 54,000 UK Biobank participants, of which 34,557 are European ancestry. The pQTL statistics were generated based on Olink Explore 3072. The largest sample size of plasma pQTL statistics based on European ancestry to date was from deCODE (by Ferkingstad, E. et al. in 2021), which included 35,559 Icelandic individuals. A total of 4,907 plasma proteins were measured using the SomaScan multiplex aptamer assay (version 4).

We extracted instrumental variants for each plasma protein in its cis-region (a 1Mb window around transcription start site of the protein-encoding gene) with the parameters r^2^<0.001, P< 5×10^−8^ and MAF>0.01. The strength of the instrumental variables was quantified by the F-statistic. An F-statistic greater than 10 indicated the absence of weak bias in the instrumental variables. LD calculations were based on European reference panel of the 1000 Genomes Project Phase 3. Lastly, the R package Mendelian Randomization (version 0.7.0) was employed to statistical analysis. We applied FDR correction for multiple testing of MR results (P.adjusted<0.05).

In the sensitivity analyses, we utilized steiger test to assess whether the MR results were affected by potential reverse causality. When the p-value of steiger test is less than 0.05, it indicates that the results are not susceptible to potential reverse causality. For the MR analyses of multiple genetic variants considered as instrumental variables, we used MR-Egger regression intercept test and Cochran's Q test to respectively assess the presence of pleiotropy and heterogeneity of the results.

**14.Bayesian colocalization analysis**

Colocalization analyses were conducted using the coloc R package (version 5.2.1) to further investigate the proteins-HF pairs were driven by single causal SNV(17). The protein loci were defined as the cis-regulatory regions encoding the protein genes. The bayesian colocalization model operates on the assumption that there is at most only one causal SNV is associated to each trait in the locus region. Consequently, five mutually exclusive hypotheses were developed: H0, proposing the absence of any SNV associated with both the protein and HF; H1, proposing the existence of only one SNV associated with the protein; H2, proposing the existence of only one SNV associated with HF; H3, proposing the existence of distinct SNVs associated with the protein and HF; and H4, proposing the existence of single SNV associated with both the protein and HF. The prior probabilities were set to 1×10^−4^ for P1 when a SNV was related only to trait one, 1×10^−4^ for P2 when a SNV was only related to trait two, and 1×10^−5^ for P12 when a SNV was related to both traits. If the posterior probability for H4 (PP.H4) exceeded 0.7, the protein-HF pairs were driven by single causal SNV in the LD region.

**15.Phenome-wide mendelian randomization**

We used the phenome-wide MR to study the non-HF potential pleiotropic or side effects of the protein targets. The protein targets identified by the above MR and colocalization analyses were set as exposure, and summary statistics of 1400 non-HF phenotypes were set as outcomes. The non-HF summary statistics utilized in this study were obtained from the UK Biobank datasets(18). We then performed the analysis using the same parameters as for MR and sensitivity analyses. Potential causal effects were considered statistically significant when the FDR-corrected p-value was less than 0.05.
