## Supplementary Results for "Exploring novel risk loci for heart failure and the shared genetic etiology with blood lipids, blood pressure, and blood glucose: a large-scale multi-trait association analysis"

**1. Pleiotropic medical indications or potential side effects for protein targets**

We conducted phenome-wide MR analysis using 1400 non-HF phenotype data from the UK Biobank, encompassing a total of 17 phenotype categories(1). This analytical approach aims to assess the pleiotropic medical indications and potential side effects for protein targets identified above. The findings of our study indicate that 30 of the 35 protein targets identified did not exhibit genetically predicted potential side effects, and many have additionally potential therapeutic benefits for other important clinical diseases. For instance, HF protective proteins LPL and CELSR2 have beneficial effects on atherosclerotic cardiovascular disease, hyperlipidemia, and hypercholesterolemia. The HF risk protein NMT1 is also an important risk protein for hypertension, left bundle branch block and asthma. However, we also observed that a total of 5 protein targets may have potential side effects. HF risk proteins CSK and ULK3 have potential protective effects against allergy unspecified not elsewhere classified and Intervertebral disc disorders. The risk protein for HF, INHBC, is also a risk protein for myocardial infarction and gout, but it has a potential protective effect on urinary organ cancer. The risk protein for HF, NCAN, was also a risk protein for disorders of lipoid metabolism but had a protective effect on chronic liver disease, cirrhosis, diabetes and adjustment reaction of the nervous system. The HF protective protein FES, although beneficial for atherosclerotic cardiovascular disease, hyperlipidemia, and hypercholesterolemia, increases the risk of carbuncle and furuncle.
