## Supplementary Discussion for "Exploring novel risk loci for heart failure and the shared genetic etiology with blood lipids, blood pressure, and blood glucose: a large-scale multi-trait association analysis"

In current use or developed in the past drugs, we discovered lipid modifying agents, beta blocking agents, and antivirals that have potential as therapeutic agents for HF. Beta-blockers have been the mainstay drugs for HF. We identified the pharmaceutical potential of lipid modifying agents such as mipomersen and clofibrate, and gemfibrozil. However, there is no direct evidence that these lipid-modifying drugs are beneficial for HF at present, and therefore may need to be interpreted with caution in clinical decision-making. In this study, we did not find convincing evidence of significant associations between statin-related loci or genes with HF in post-GWAS analyses. There have been different views on the effect of statin therapy on heart failure (1-3), and a previous MR study also reported that no significant association was observed between genetically predicted statin-related target and the risk of HF(4). Therefore, whether the widely used statins are beneficial for HF remains to be further evaluated at present. Among the antivirals for systemic use, we located the ADORA2B and CES1 genes, which encode targets for vidarabine and oseltamivir. A recent study revealed that the plasma N-acetylneuraminic acid level in patients with heart failure was significantly elevated, and the higher the index, the worse the prognosis. Interestingly, the commonly used anti-influenza drug oseltamivir can effectively inhibit this and improve cardiac dysfunction(5). Previous studies have also found the potent and selective inhibitory properties of vidarabine on AC5, as well as its efficacy in mitigating the advancement of cardiomyopathy in animal models. (6). In addition, we identified numerous novel potential protein targets for HF, the majority of which remain within the clinical discovery phase. Phenome-wide mendelian randomization analyses exhibited a promising reality where the majority of these protein targets incurred no significant adverse effects and possessed potential therapeutic implications for other important clinical diseases. Therefore, these proteins targets hold great potential for the development of novel drugs for effective primary prevention of HF in the future.
