## Supplementary figures and images for "Exploring novel risk loci for heart failure and the shared genetic etiology with blood lipids, blood pressure, and blood glucose: a large-scale multi-trait association analysis"

### Supplementary Figure 1

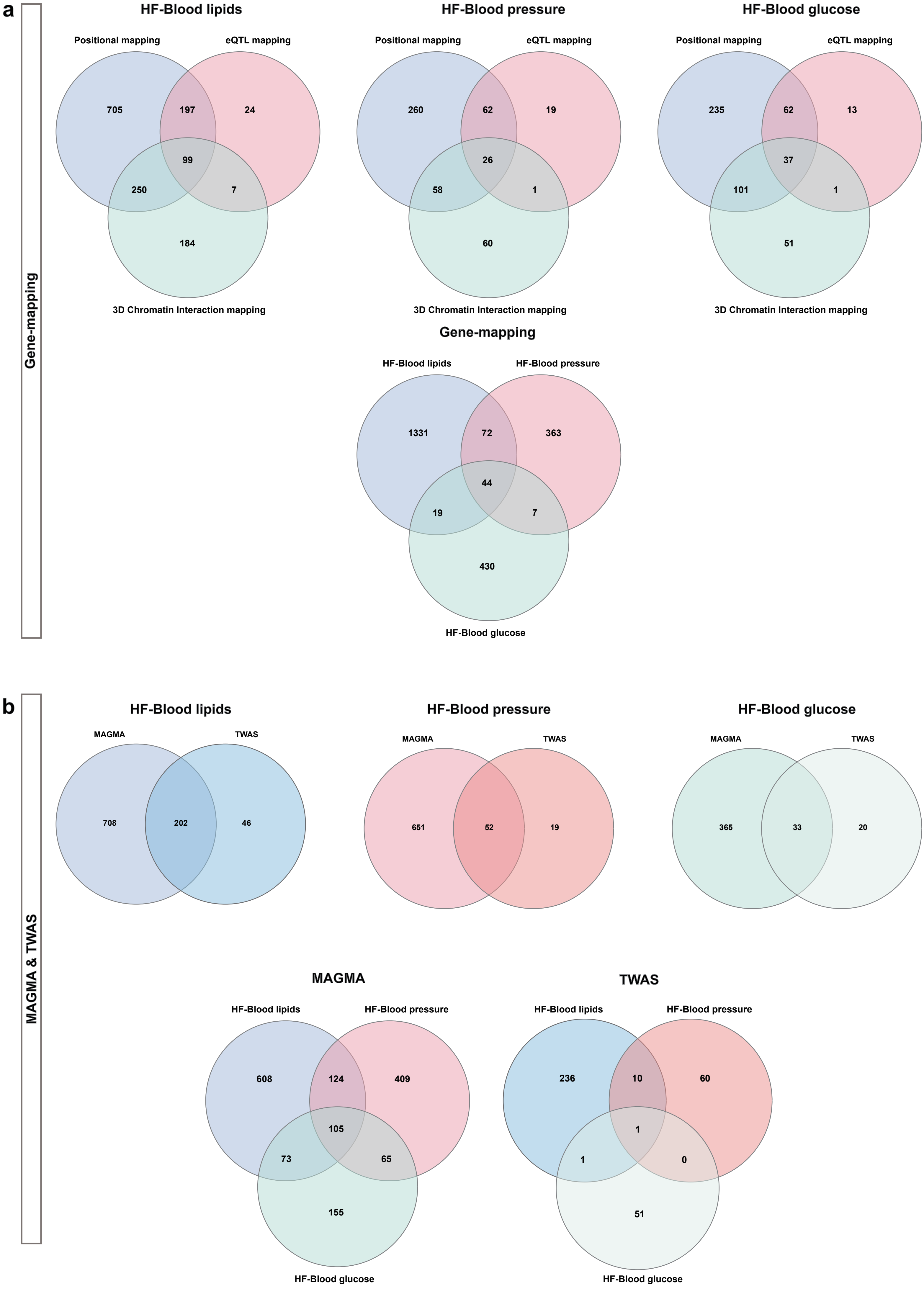

### Supplementary Figure 2

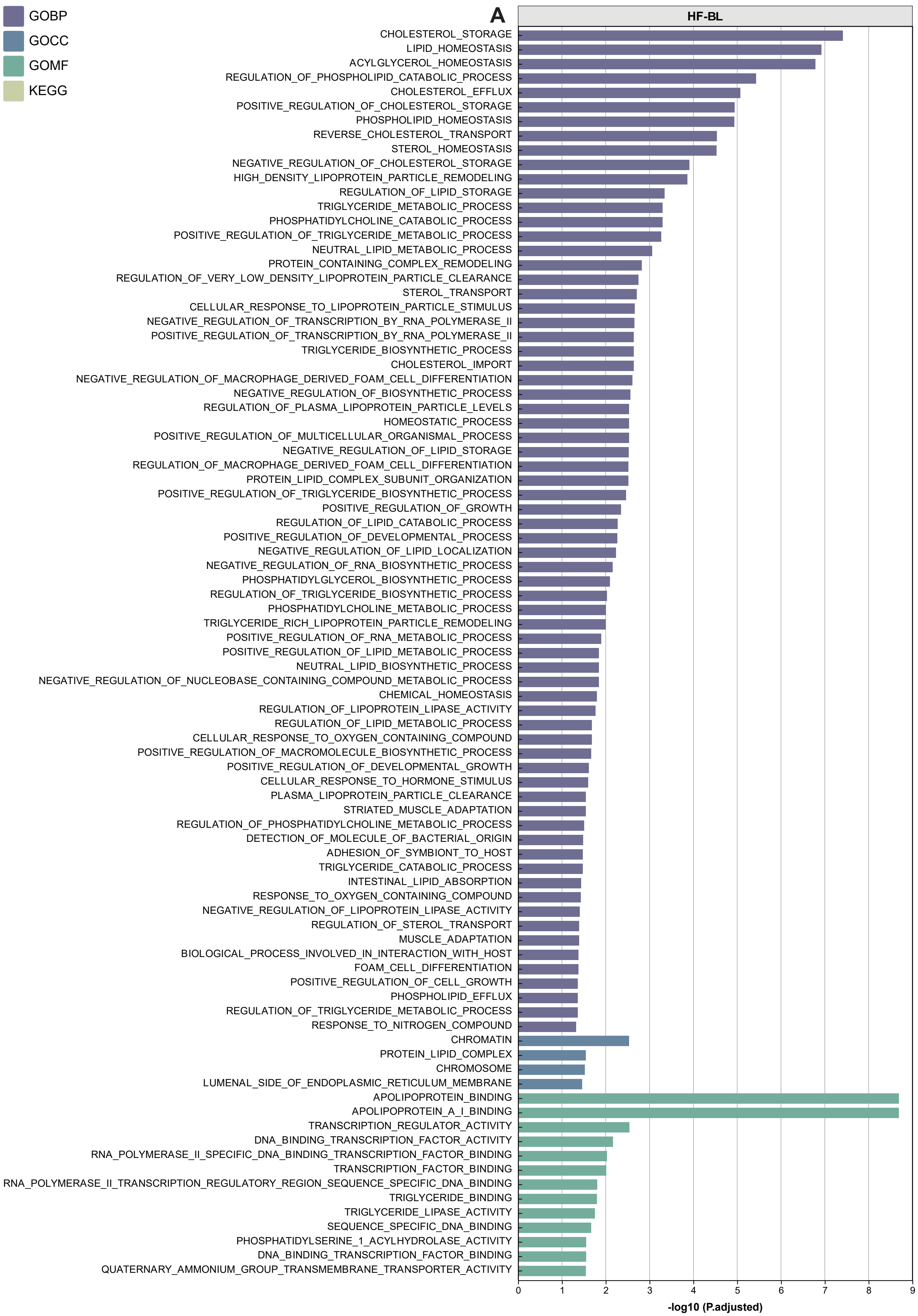

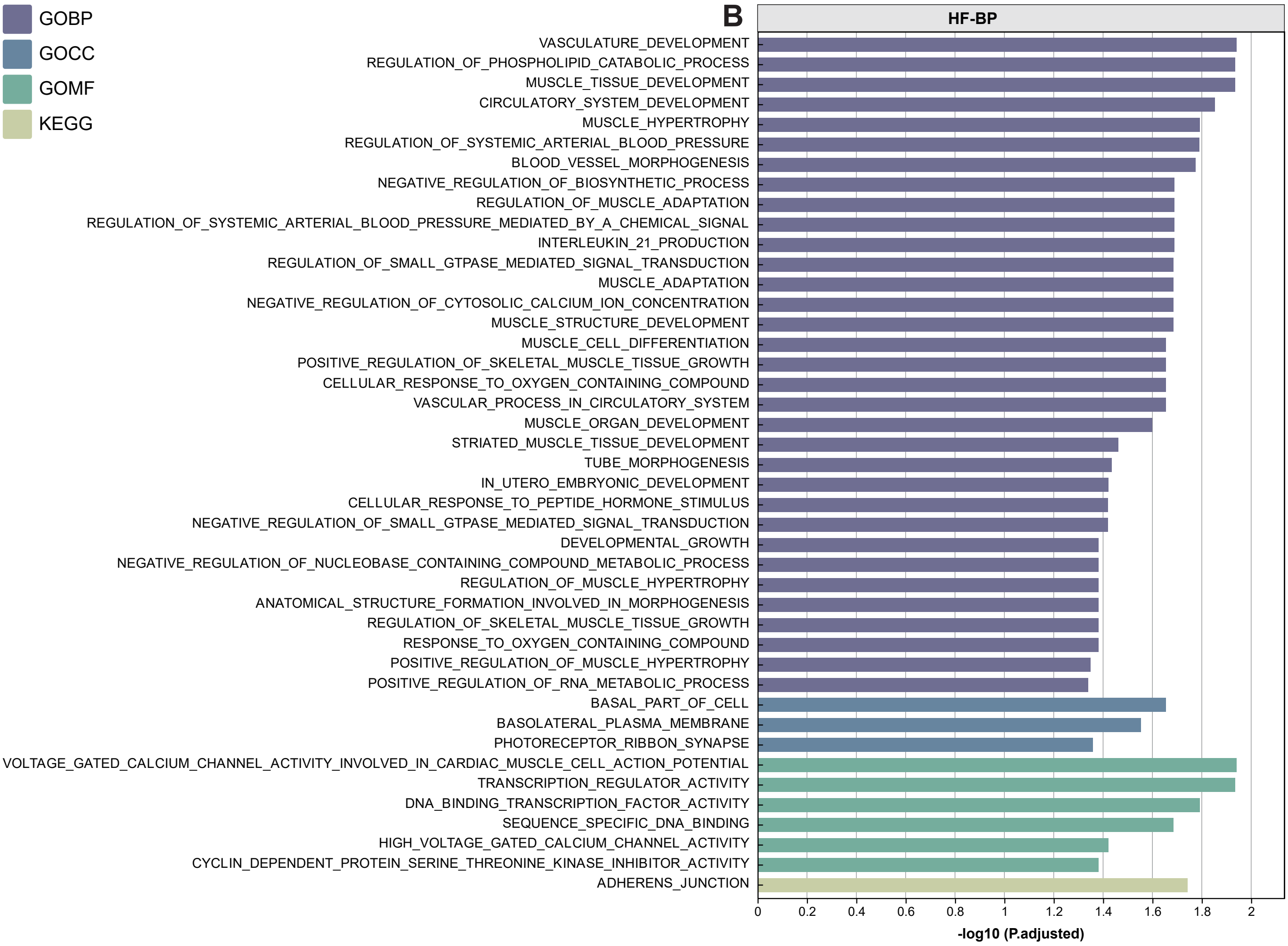

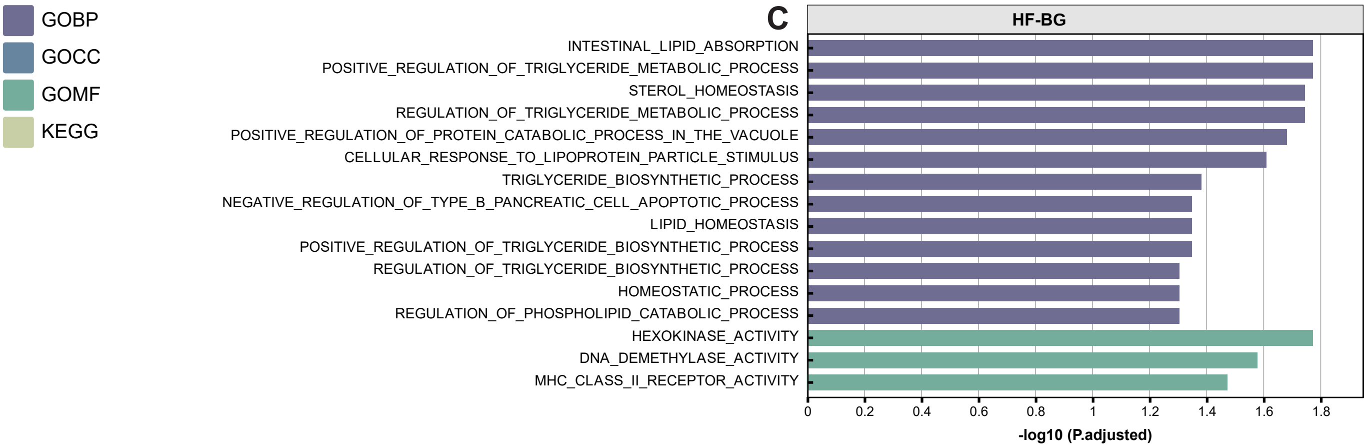

### Supplementary Figure 3

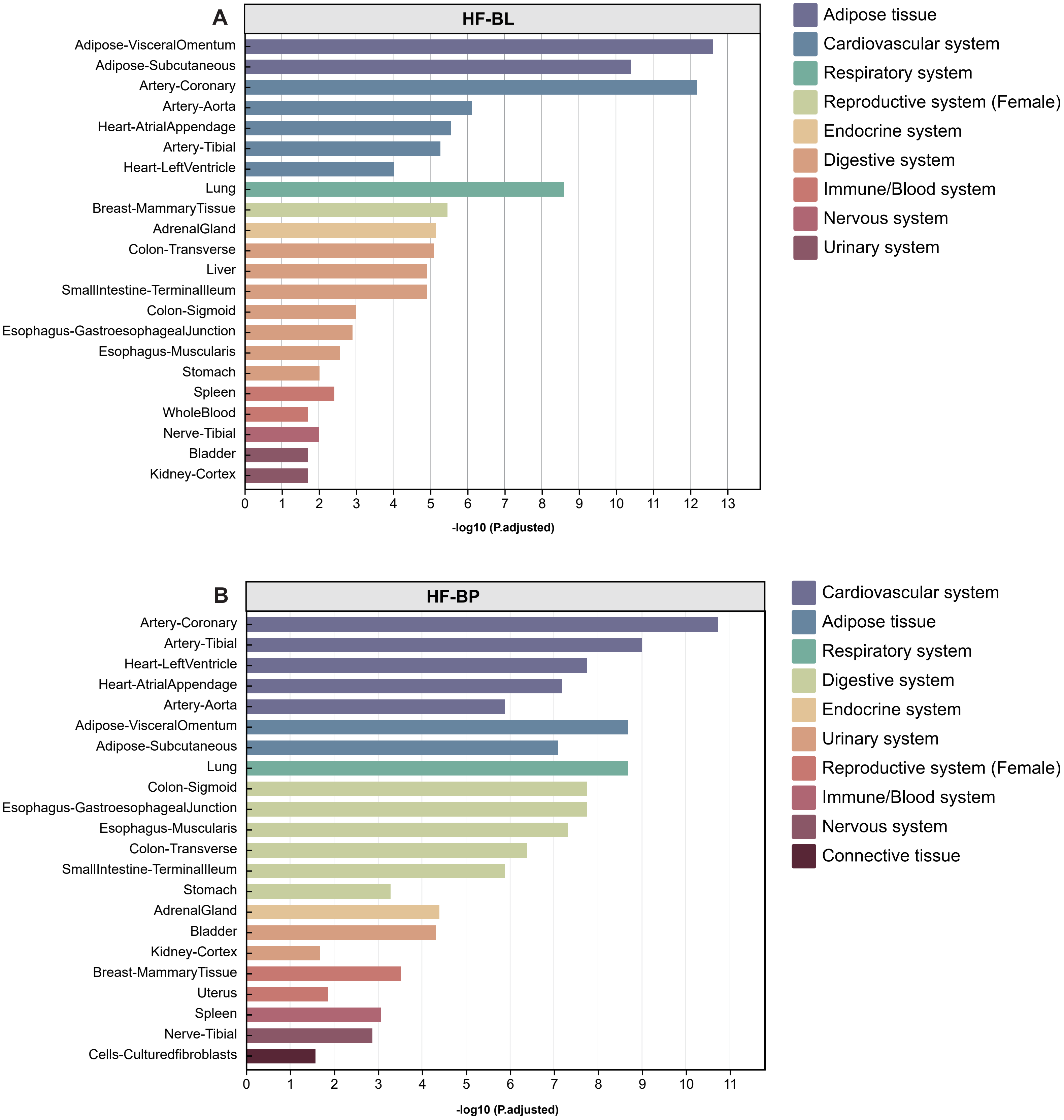

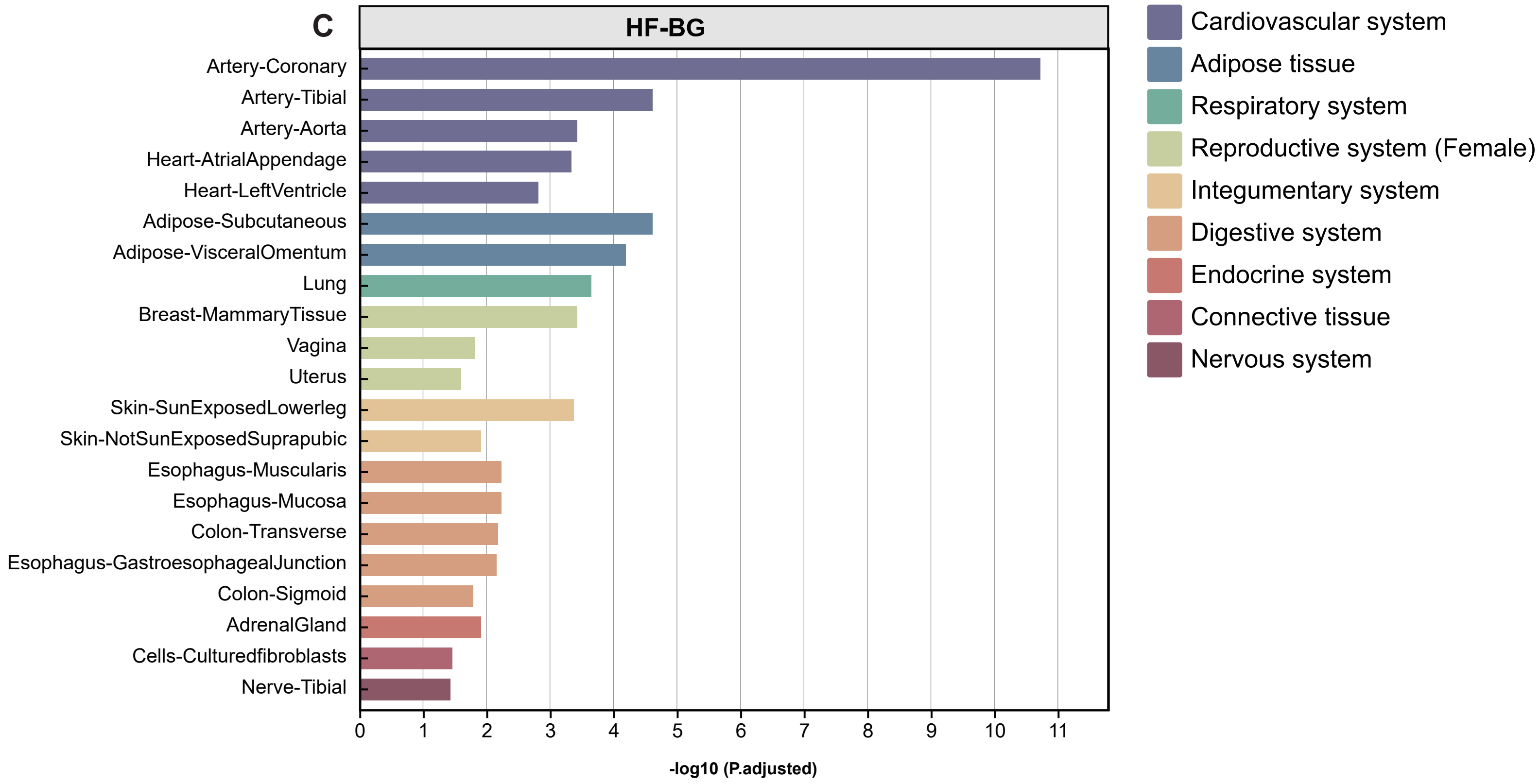

### Supplementary Figure 4

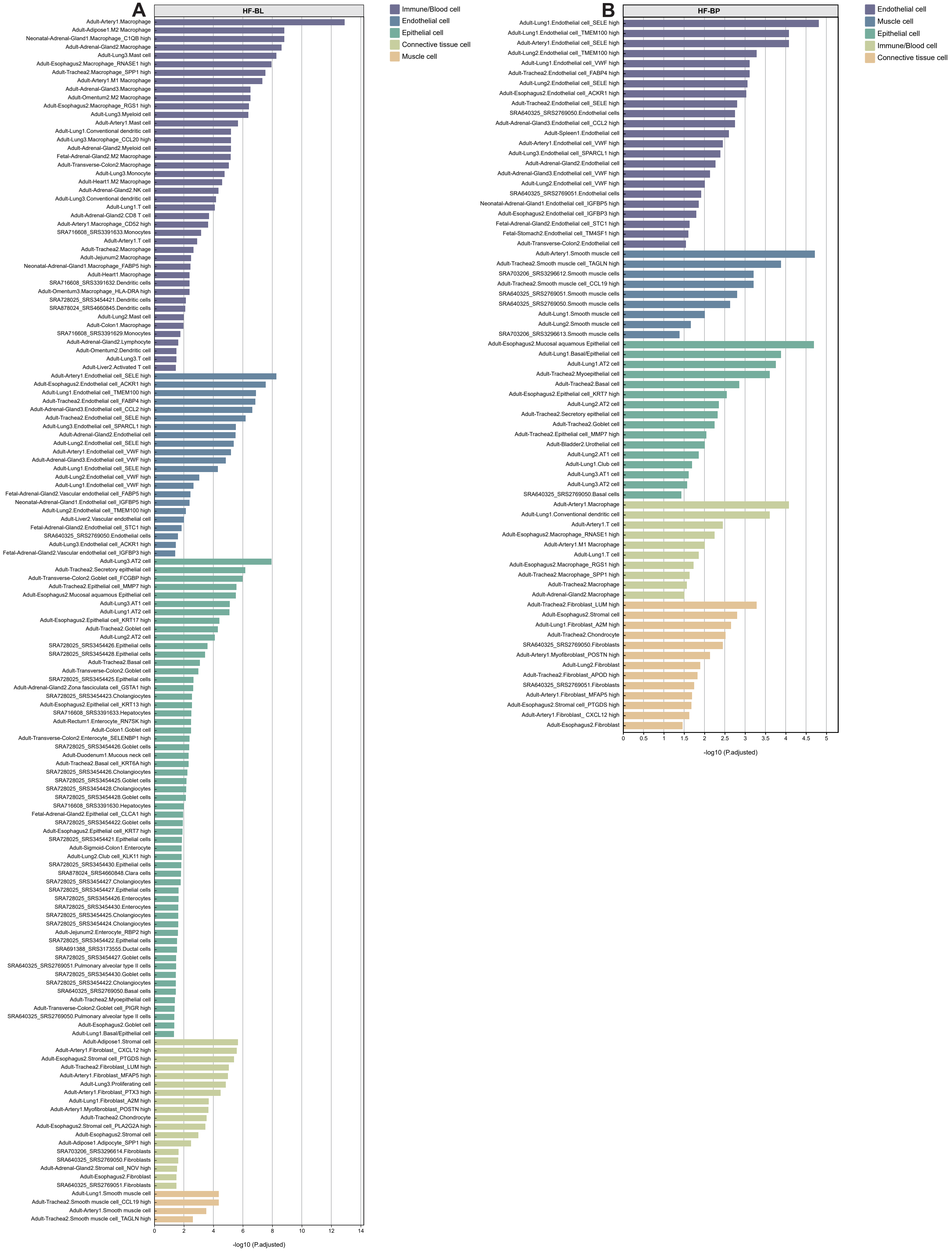

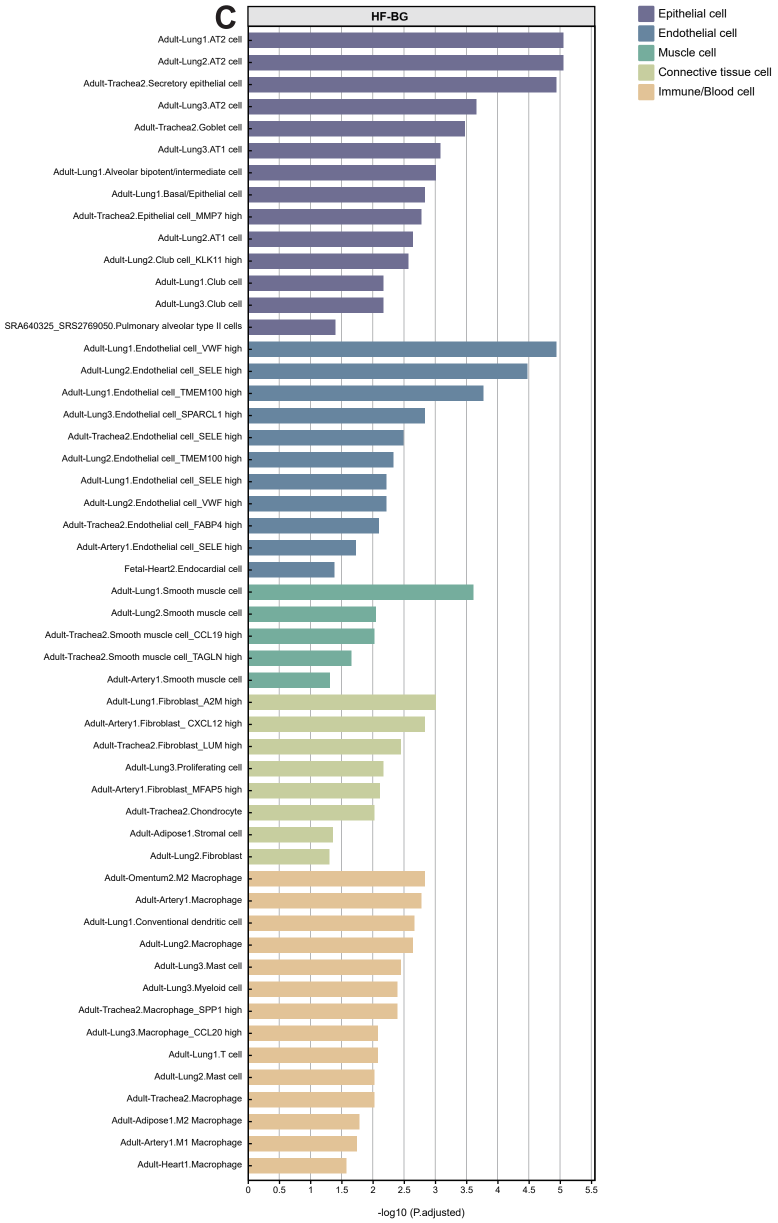

### Supplementary Figure 5

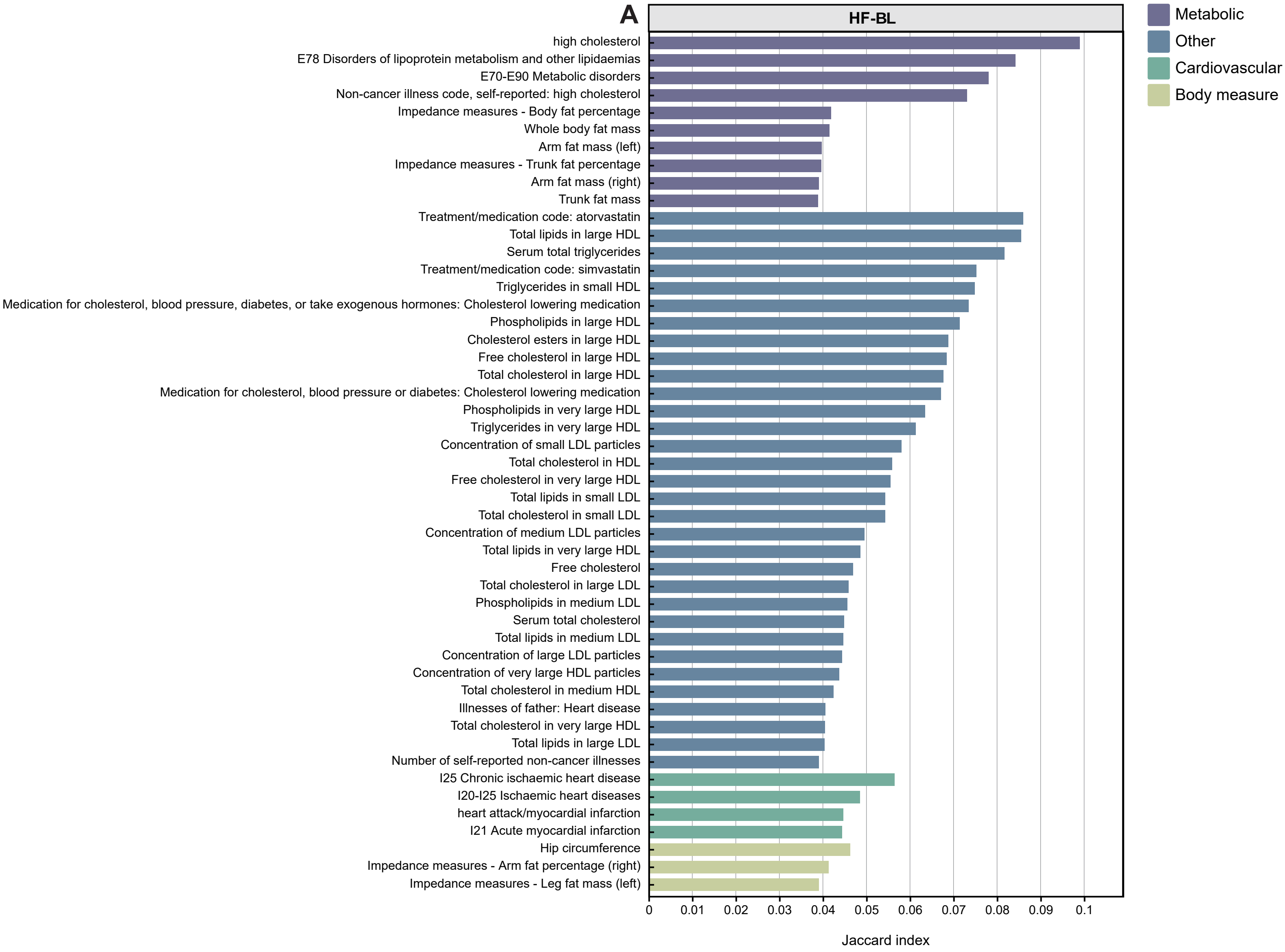

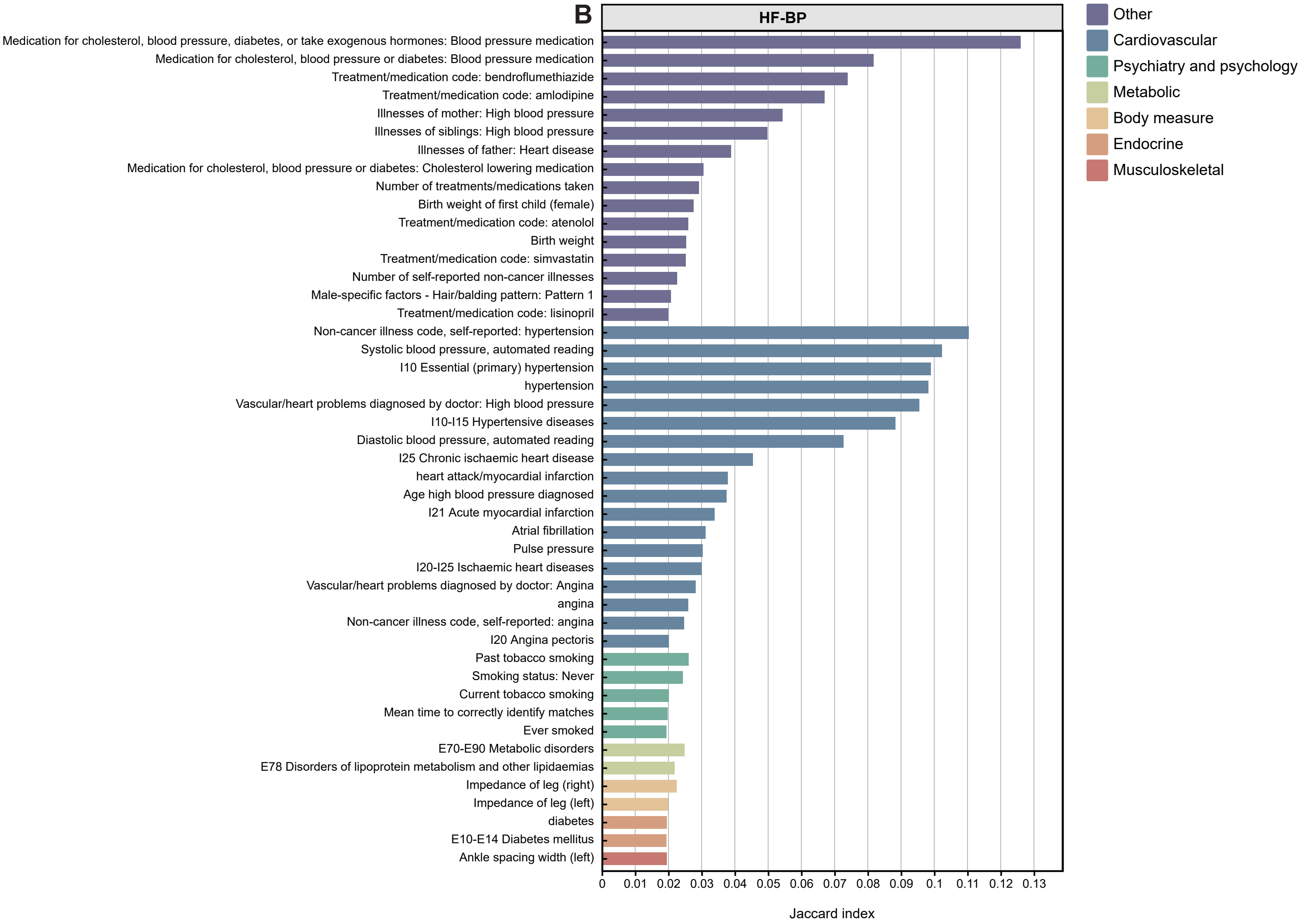

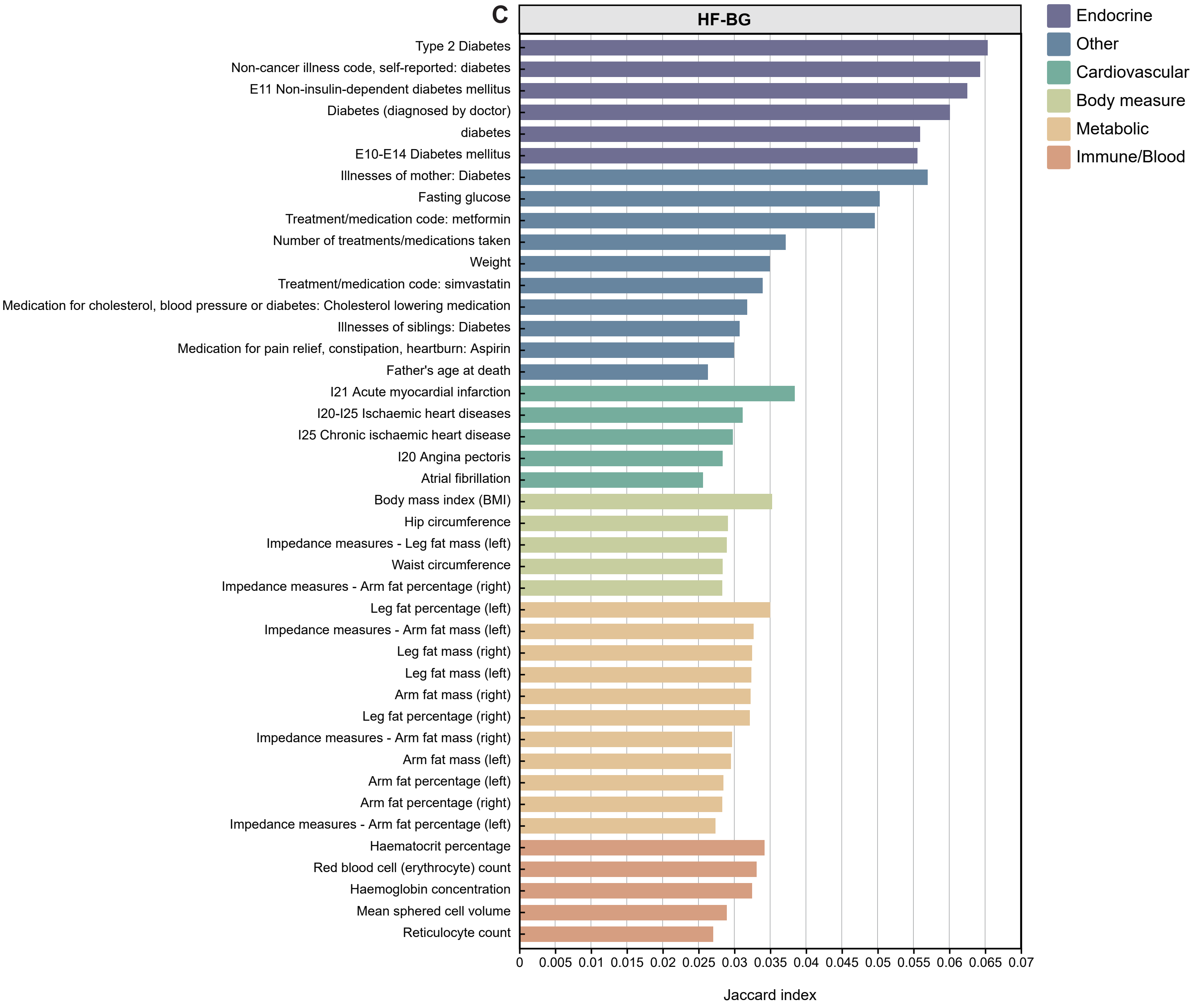
