## Supplementary Figure 6 for "Exploring novel risk loci for heart failure and the shared genetic etiology with blood lipids, blood pressure, and blood glucose: a large-scale multi-trait association analysis"

a

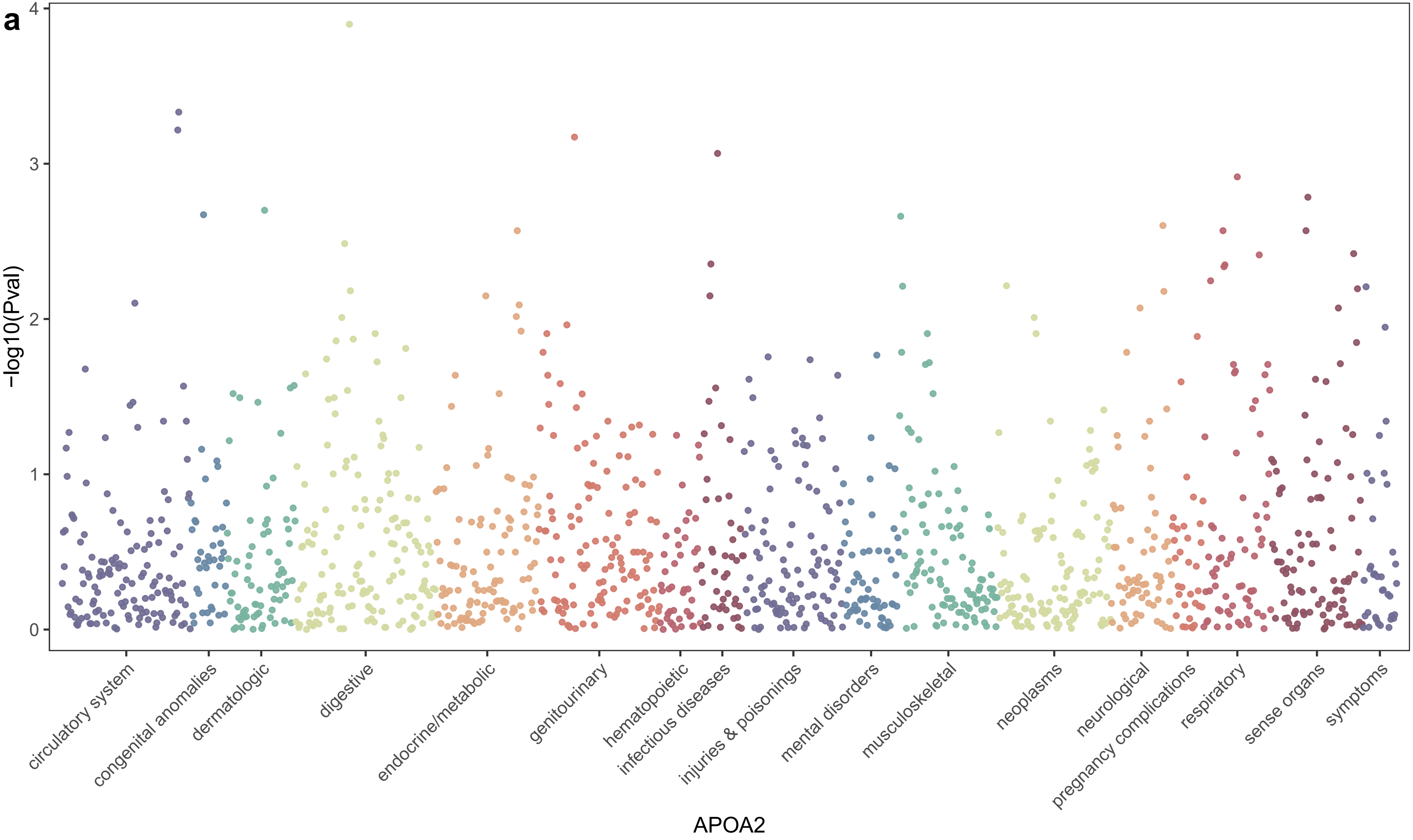

-log10(Pval)
0
1
2
3
4
circulatory system
congenital anomalies
dermatologic
digestive
endocrine/metabolic
genitourinary
hematopoietic
infectious diseases
injuries & poisonings
mental disorders
musculoskeletal
neoplasms
neurological
pregnancy complications
respiratory
sense organs
symptoms
APOA2

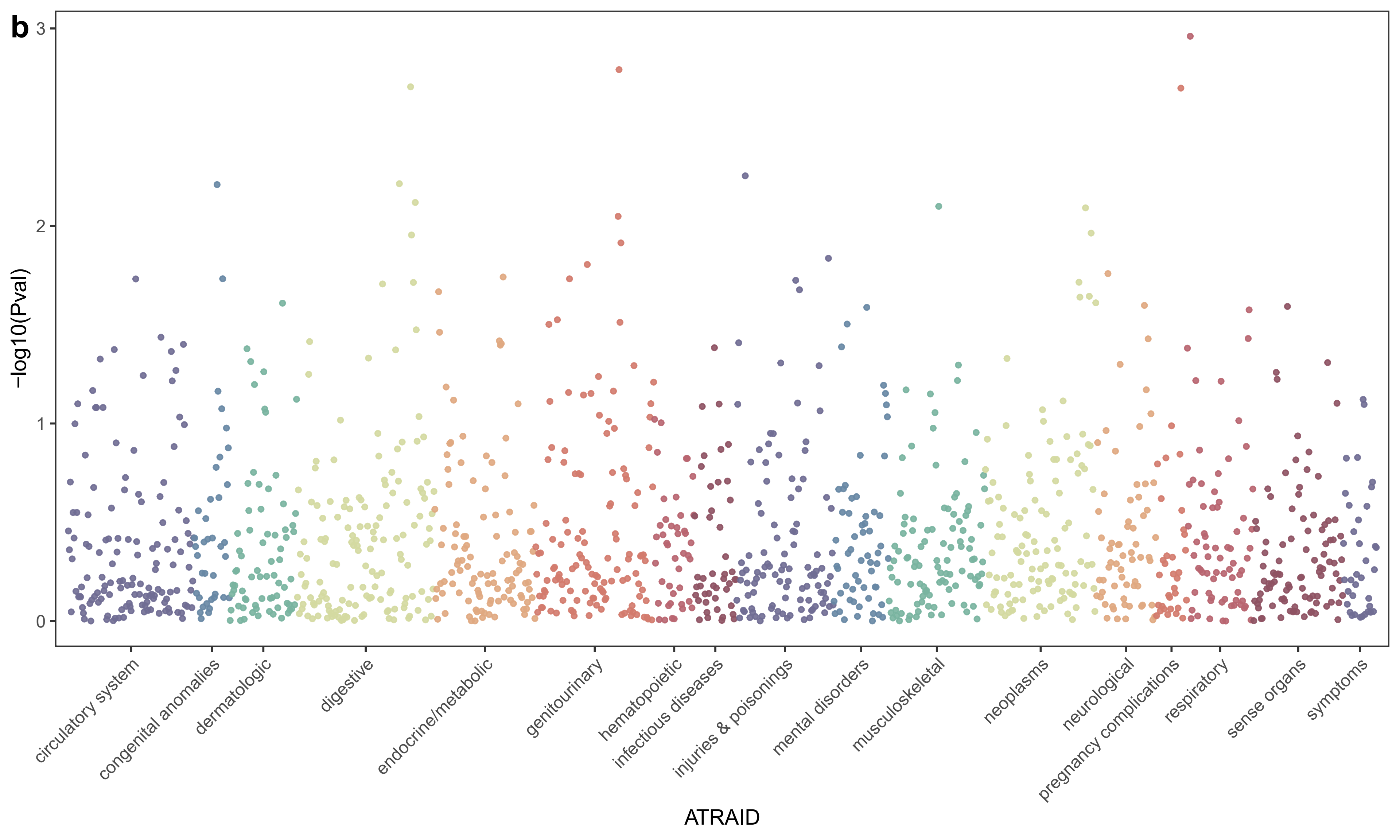

b
3
2
1
0
-log10(Pval)
circulatory system
congenital anomalies
dermatologic
digestive
endocrine/metabolic
genitourinary
hematopoietic
infectious diseases
injuries & poisonings
mental disorders
musculoskeletal
neoplasms
neurological
pregnancy complications
respiratory
sense organs
symptoms
ATRAID

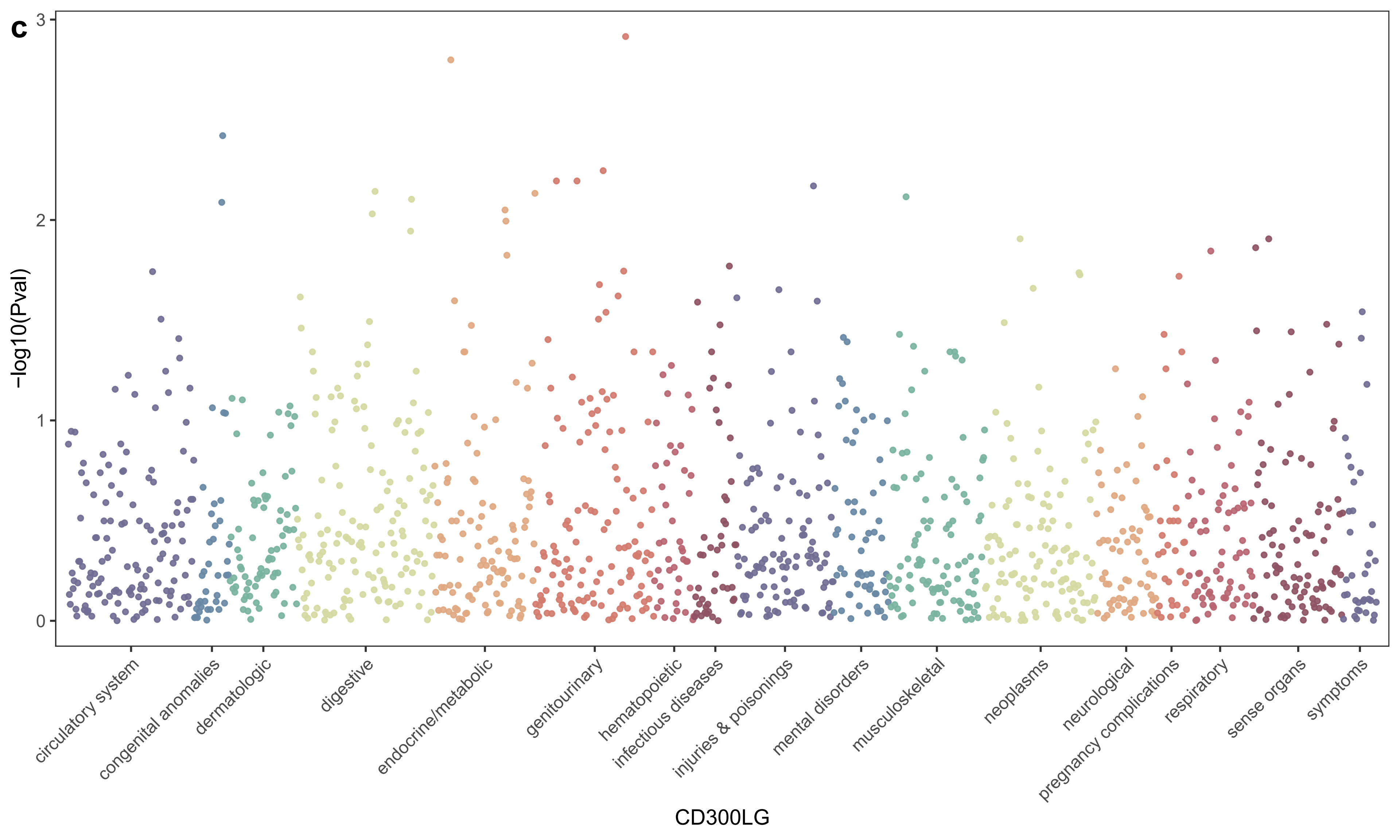
c
3
2
1
0
-log10(Pval)
circulatory system
congenital anomalies
dermatologic
digestive
endocrine/metabolic
genitourinary
hematopoietic
infectious diseases
injuries & poisonings
mental disorders
musculoskeletal
neoplasms
neurological
pregnancy complications
respiratory
sense organs
symptoms
CD300LG

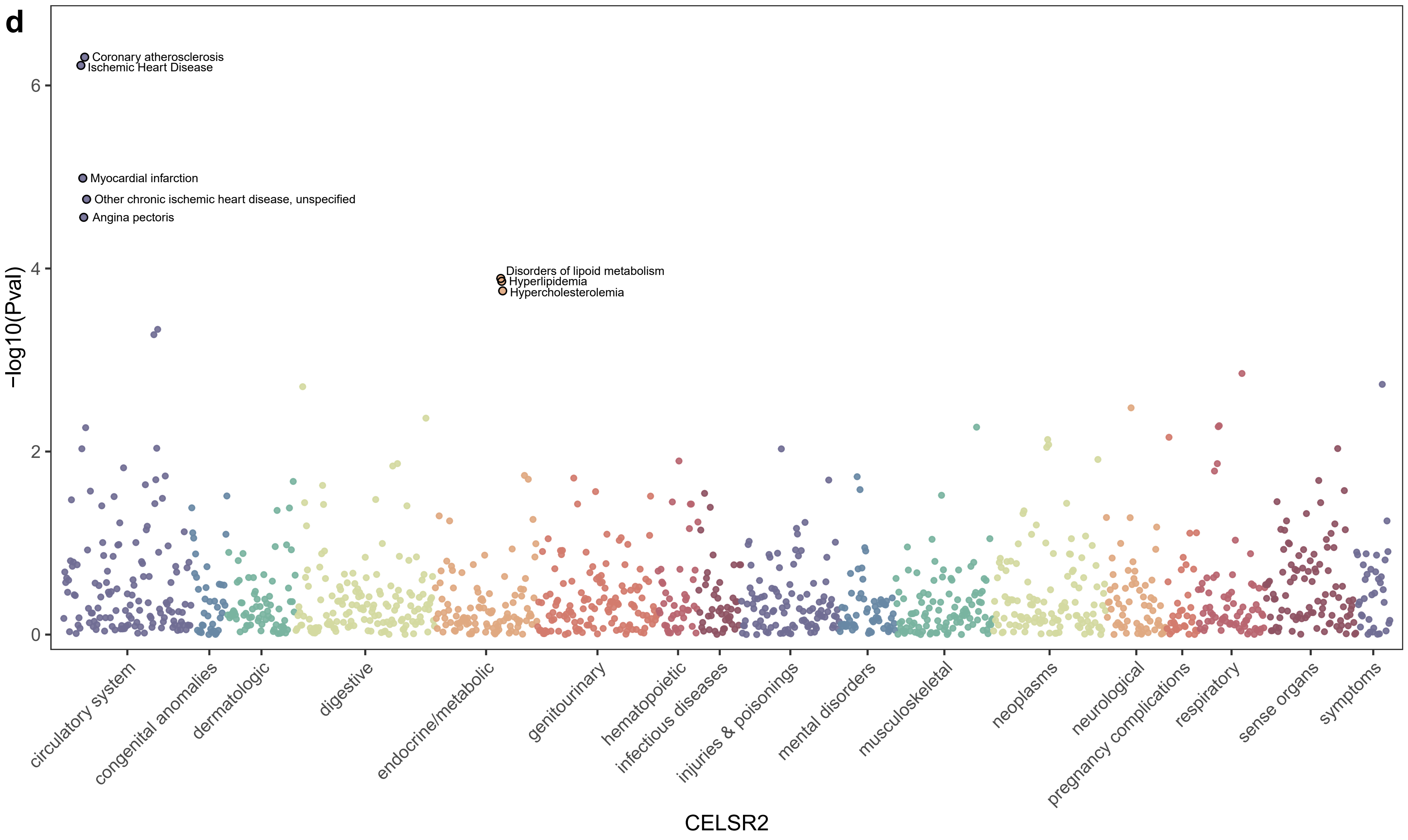
d
Coronary atherosclerosis
Ischemic Heart Disease
Myocardial infarction
Other chronic ischemic heart disease, unspecified
Angina pectoris
Disorders of lipoid metabolism
Hyperlipidemia
Hypercholesterolemia
-log10(Pval)
0
2
4
6
circulatory system
congenital anomalies
dermatologic
digestive
endocrine/metabolic
genitourinary
hematopoietic
infectious diseases
injuries & poisonings
mental disorders
musculoskeletal
neoplasms
neurological
pregnancy complications
respiratory
sense organs
symptoms
CELSR2

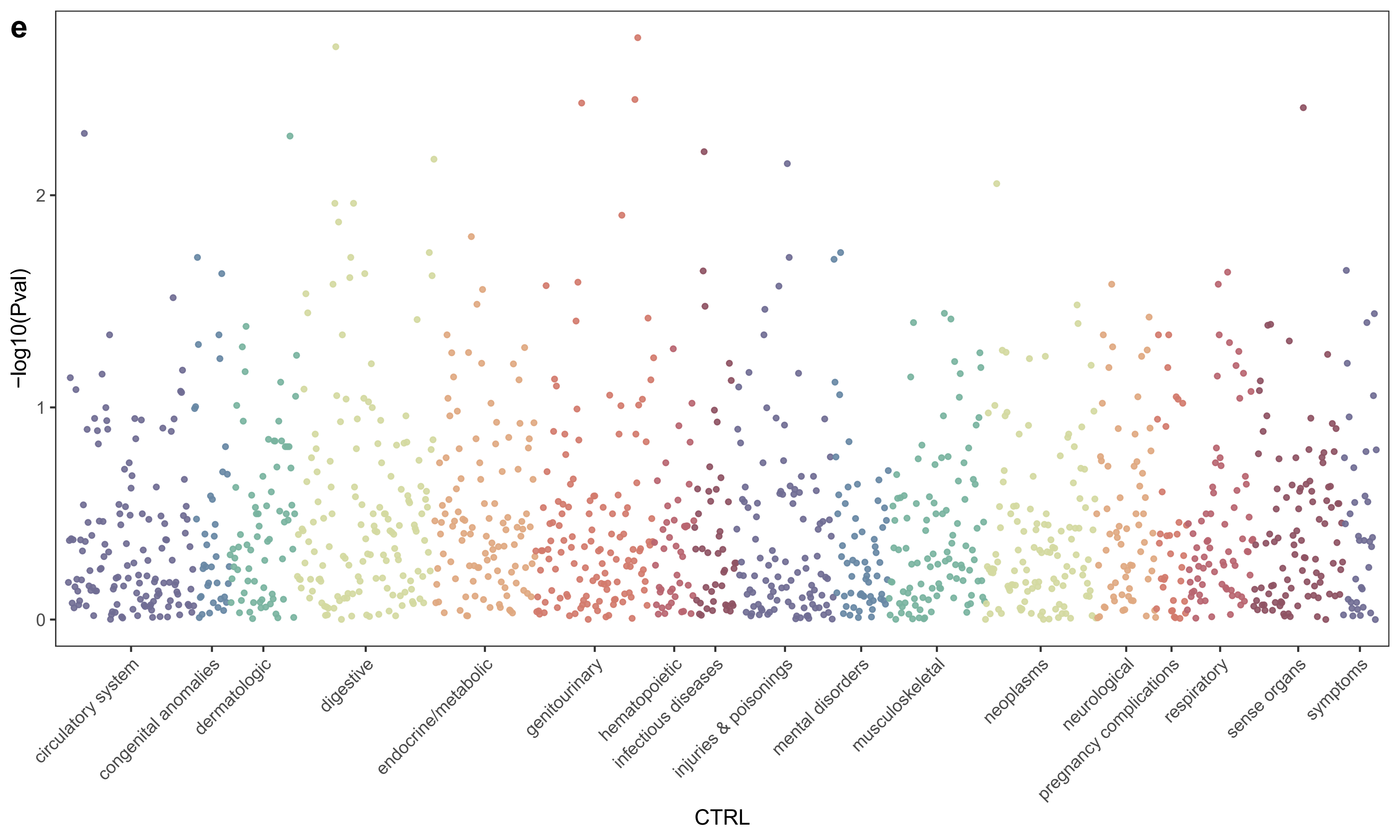

e
−log10(Pval)
0
1
2
circulatory system
congenital anomalies
dermatologic
digestive
endocrine/metabolic
genitourinary
hematopoietic
infectious diseases
injuries & poisonings
mental disorders
musculoskeletal
neoplasms
neurological
pregnancy complications
respiratory
sense organs
symptoms
CTRL

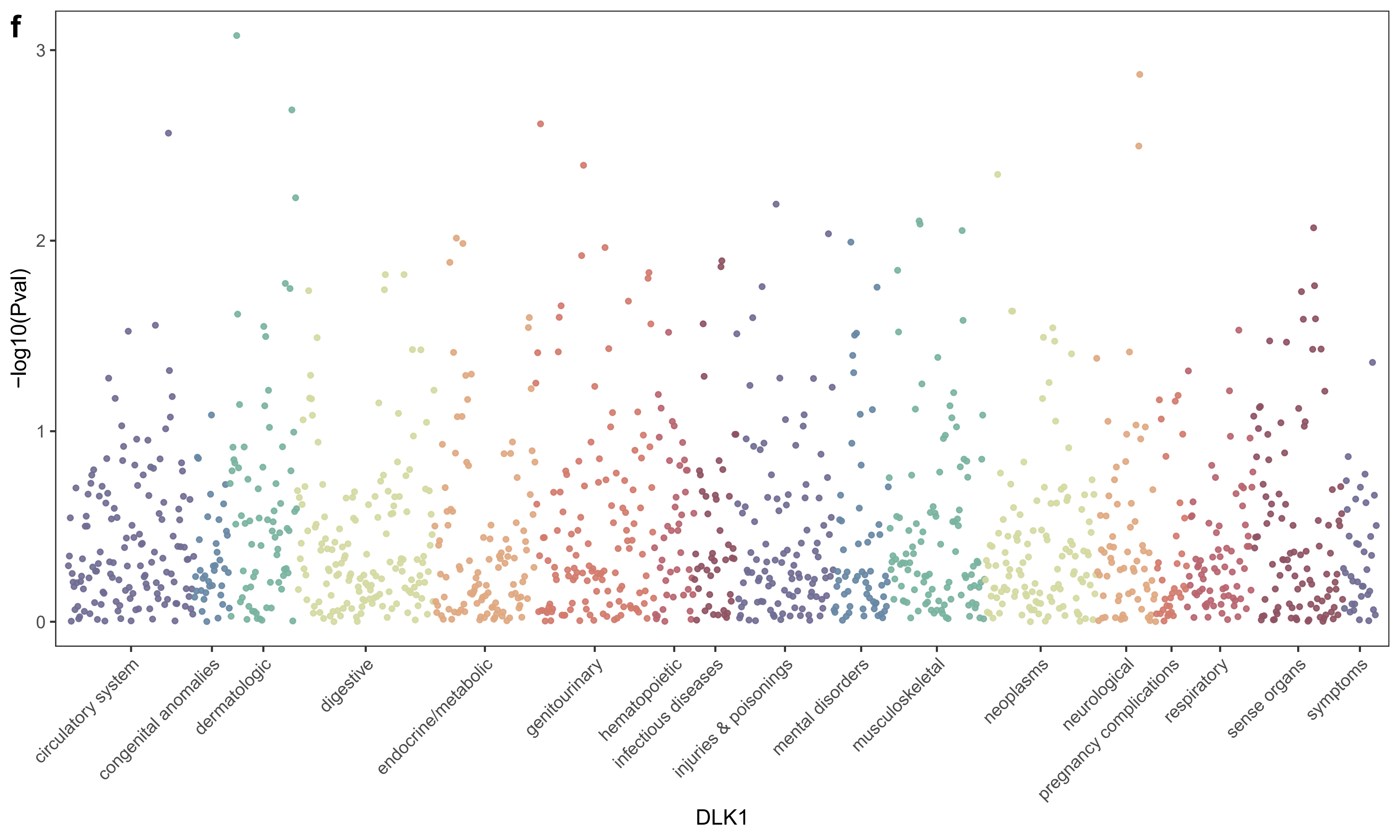

f
−log10(Pval)
3
2
1
0
circulatory system
congenital anomalies
dermatologic
digestive
endocrine/metabolic
genitourinary
hematopoietic
infectious diseases
injuries & poisonings
mental disorders
musculoskeletal
neoplasms
neurological
pregnancy complications
respiratory
sense organs
symptoms
DLK1

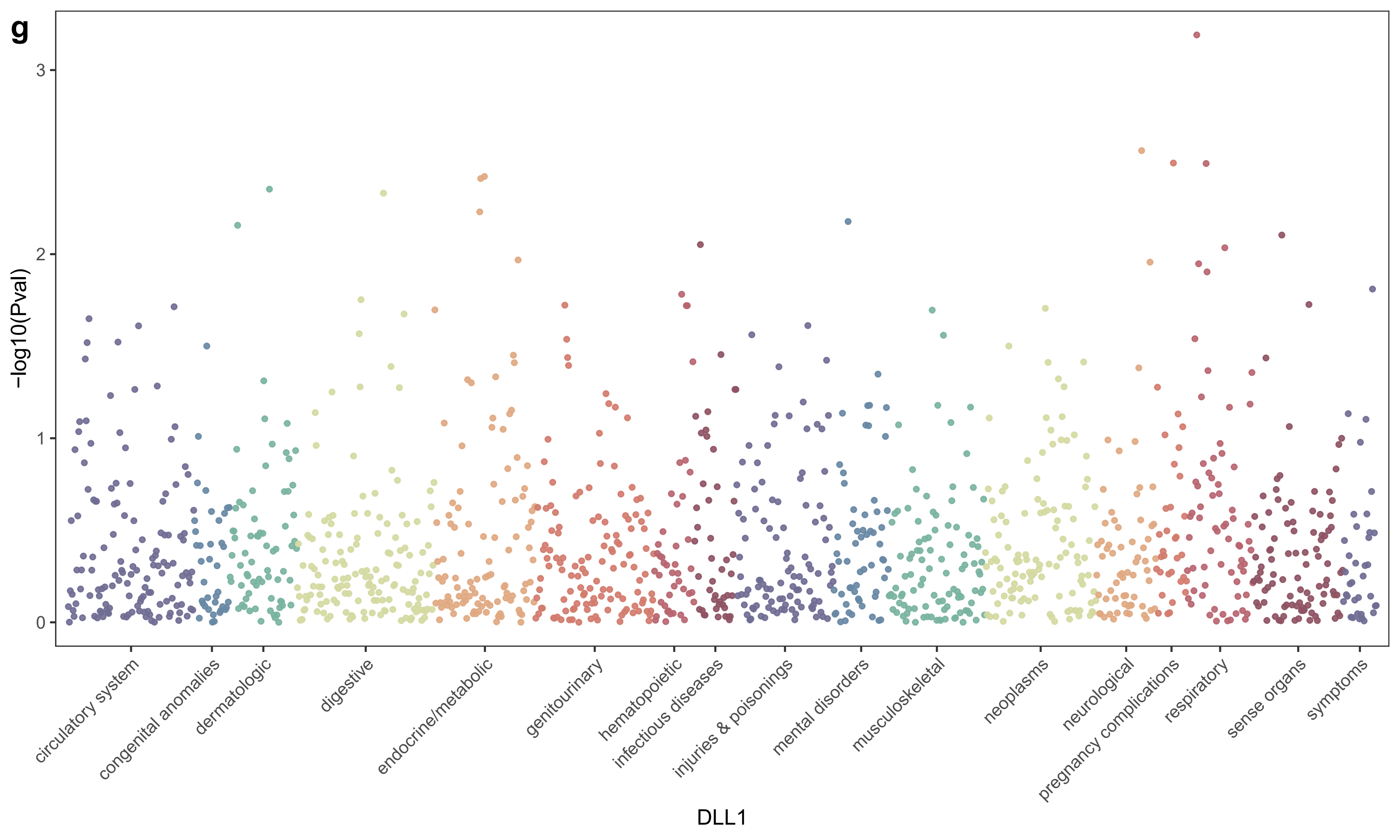
g
-log10(Pval)
0
1
2
3
circulatory system
congenital anomalies
dermatologic
digestive
endocrine/metabolic
genitourinary
hematopoietic
infectious diseases
injuries & poisonings
mental disorders
musculoskeletal
neoplasms
neurological
pregnancy complications
respiratory
sense organs
symptoms
DLL1

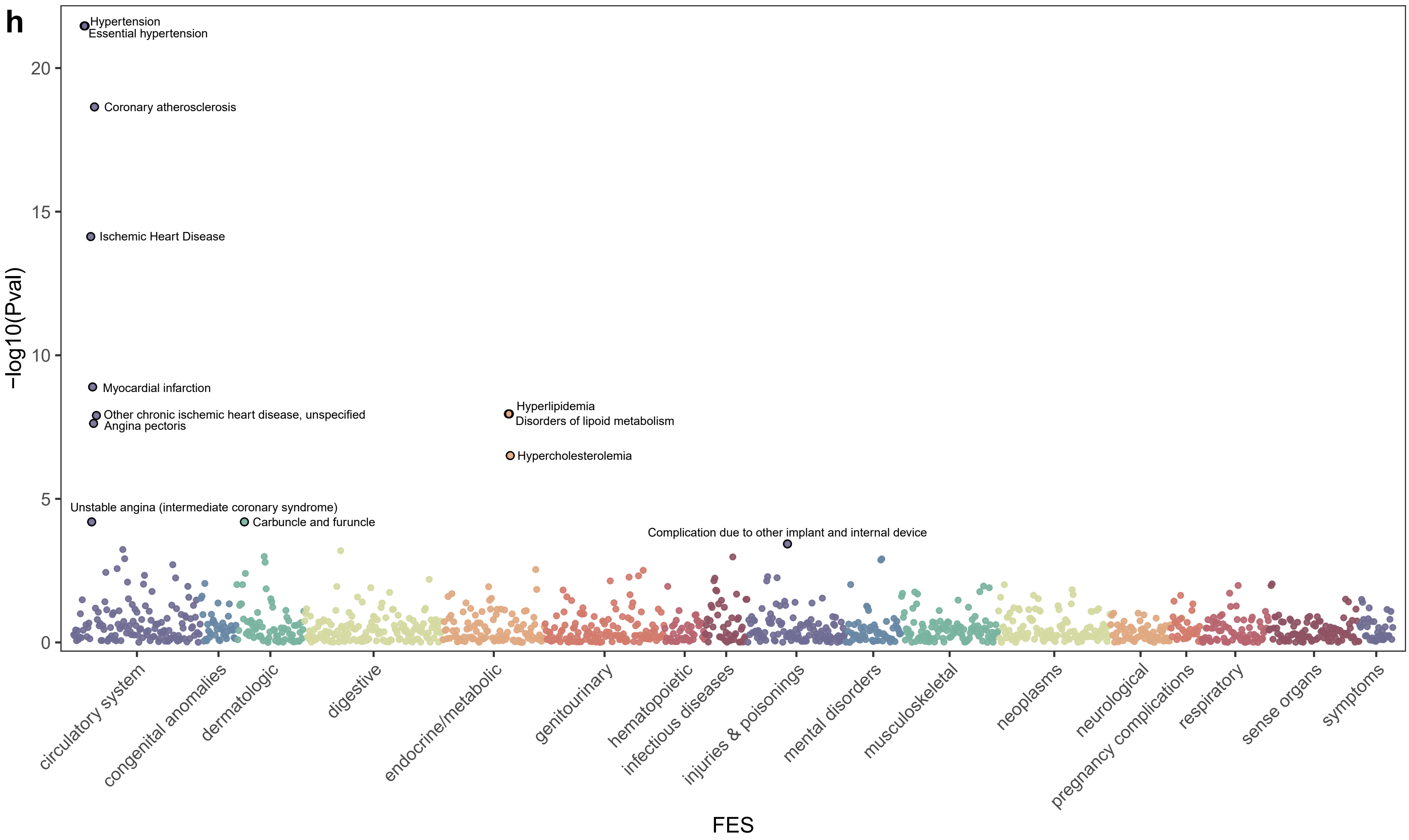
h
Hypertension
Essential hypertension
Coronary atherosclerosis
Ischemic Heart Disease
Myocardial infarction
Other chronic ischemic heart disease, unspecified
Angina pectoris
Hyperlipidemia
Disorders of lipoid metabolism
Hypercholesterolemia
Unstable angina (intermediate coronary syndrome)
Carbuncle and furuncle
Complication due to other implant and internal device
-log10(Pval)
0
5
10
15
20
circulatory system
congenital anomalies
dermatologic
digestive
endocrine/metabolic
genitourinary
hematopoietic
infectious diseases
injuries & poisonings
mental disorders
musculoskeletal
neoplasms
neurological
pregnancy complications
respiratory
sense organs
symptoms
FES

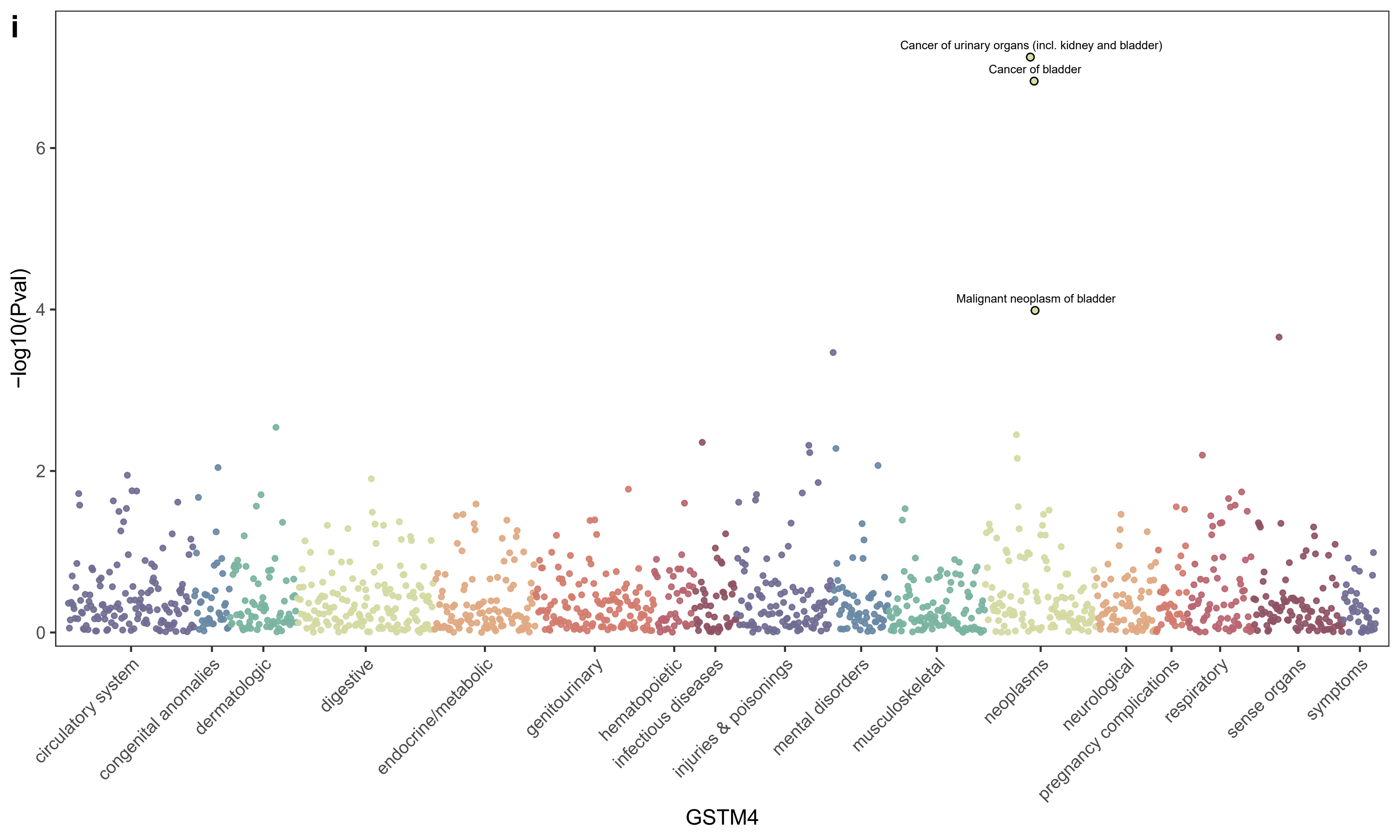

i
Cancer of urinary organs (incl. kidney and bladder)
Cancer of bladder
Malignant neoplasm of bladder
-log10(Pval)
0
2
4
6
circulatory system
congenital anomalies
dermatologic
digestive
endocrine/metabolic
genitourinary
hematopoietic
infectious diseases
injuries & poisonings
mental disorders
musculoskeletal
neoplasms
neurological
pregnancy complications
respiratory
sense organs
symptoms
GSTM4

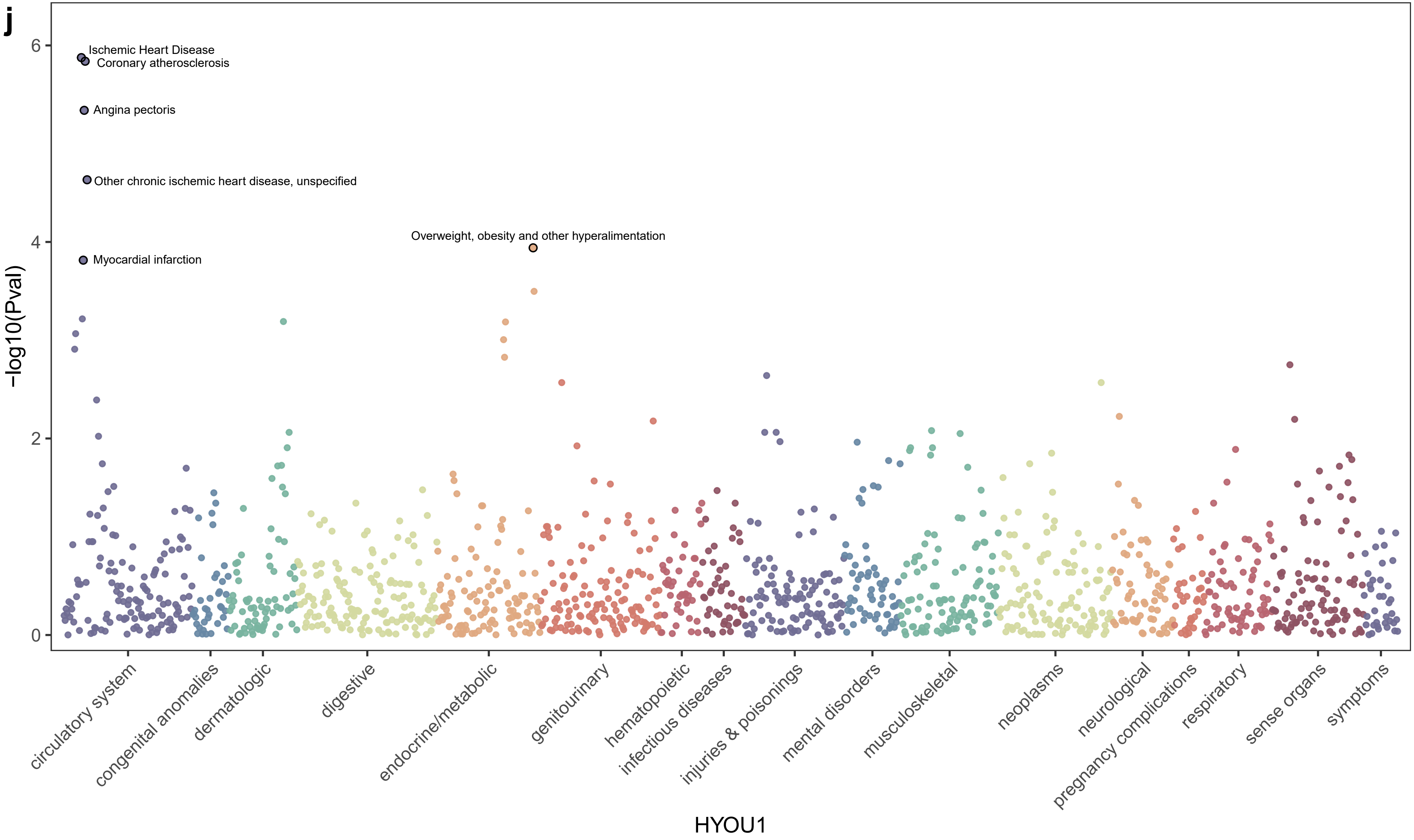
j
Ischemic Heart Disease
Coronary atherosclerosis
Angina pectoris
Other chronic ischemic heart disease, unspecified
Overweight, obesity and other hyperalimentation
Myocardial infarction
6
4
2
0
−log10(Pval)
circulatory system
congenital anomalies
dermatologic
digestive
endocrine/metabolic
genitourinary
hematopoietic
infectious diseases
injuries & poisonings
mental disorders
musculoskeletal
neoplasms
neurological
pregnancy complications
respiratory
sense organs
symptoms
HYOU1

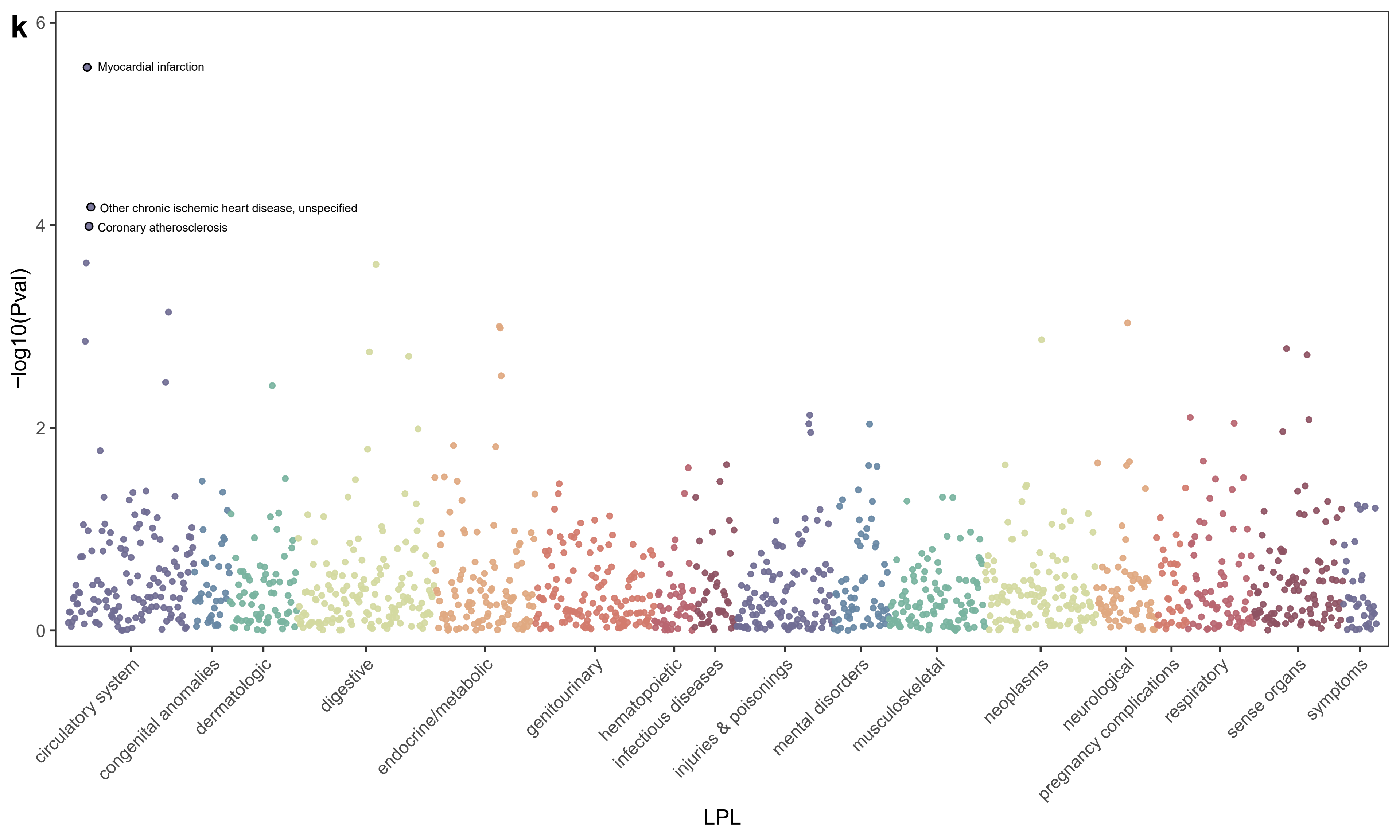
k
6
4
2
0
-log10(Pval)
Myocardial infarction
Other chronic ischemic heart disease, unspecified
Coronary atherosclerosis
circulatory system
congenital anomalies
dermatologic
digestive
endocrine/metabolic
genitourinary
hematopoietic
infectious diseases
injuries & poisonings
mental disorders
musculoskeletal
neoplasms
neurological
pregnancy complications
respiratory
sense organs
symptoms
LPL

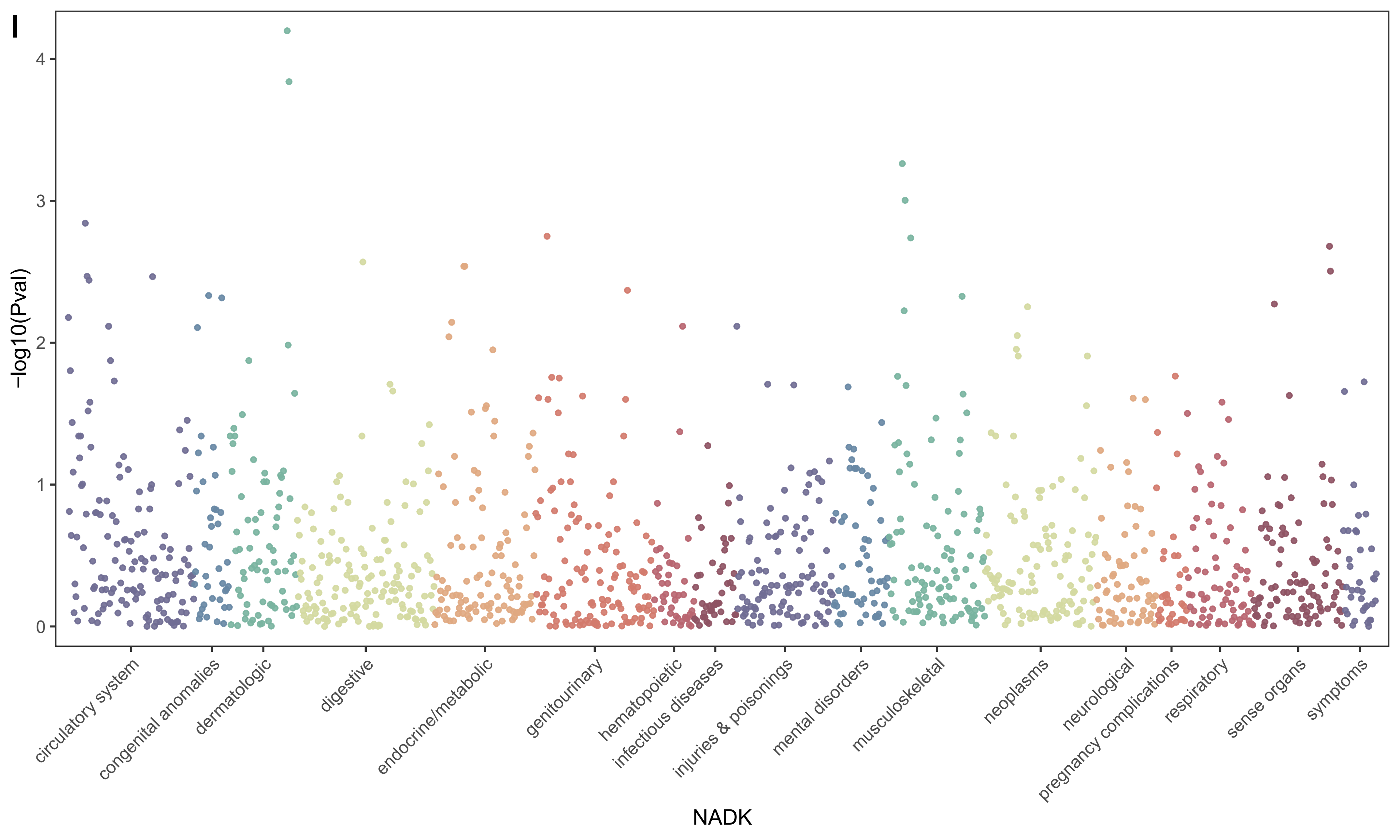
I
-log10(Pval)
4
3
2
1
0
circulatory system
congenital anomalies
dermatologic
digestive
endocrine/metabolic
genitourinary
hematopoietic
infectious diseases
injuries & poisonings
mental disorders
musculoskeletal
neoplasms
neurological
pregnancy complications
respiratory
sense organs
symptoms
NADK

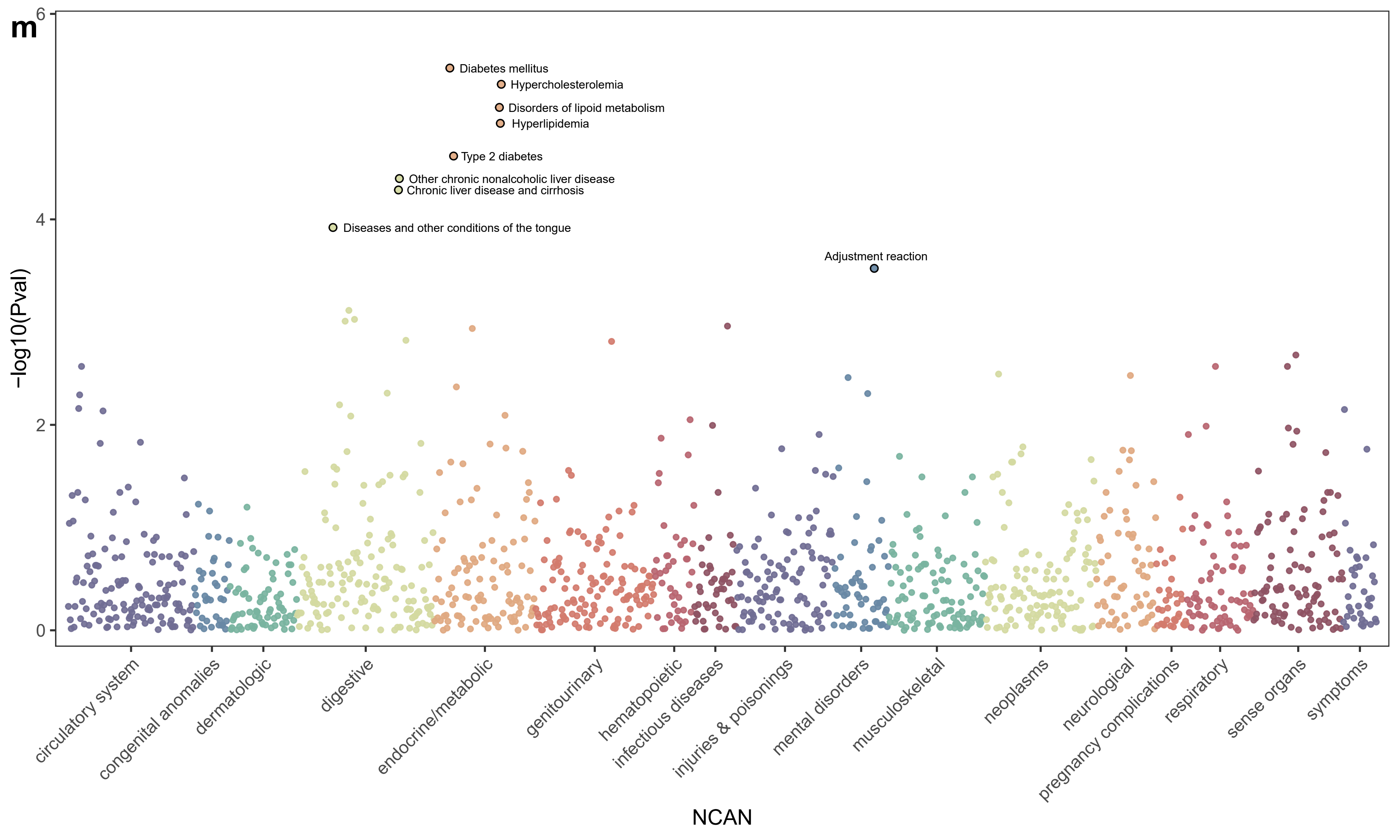
m
6
4
2
0
-log10(Pval)
Diabetes mellitus
Hypercholesterolemia
Disorders of lipoid metabolism
Hyperlipidemia
Type 2 diabetes
Other chronic nonalcoholic liver disease
Chronic liver disease and cirrhosis
Diseases and other conditions of the tongue
Adjustment reaction
circulatory system
congenital anomalies
dermatologic
digestive
endocrine/metabolic
genitourinary
hematopoietic
infectious diseases
injuries & poisonings
mental disorders
musculoskeletal
neoplasms
neurological
pregnancy complications
respiratory
sense organs
symptoms
NCAN

n

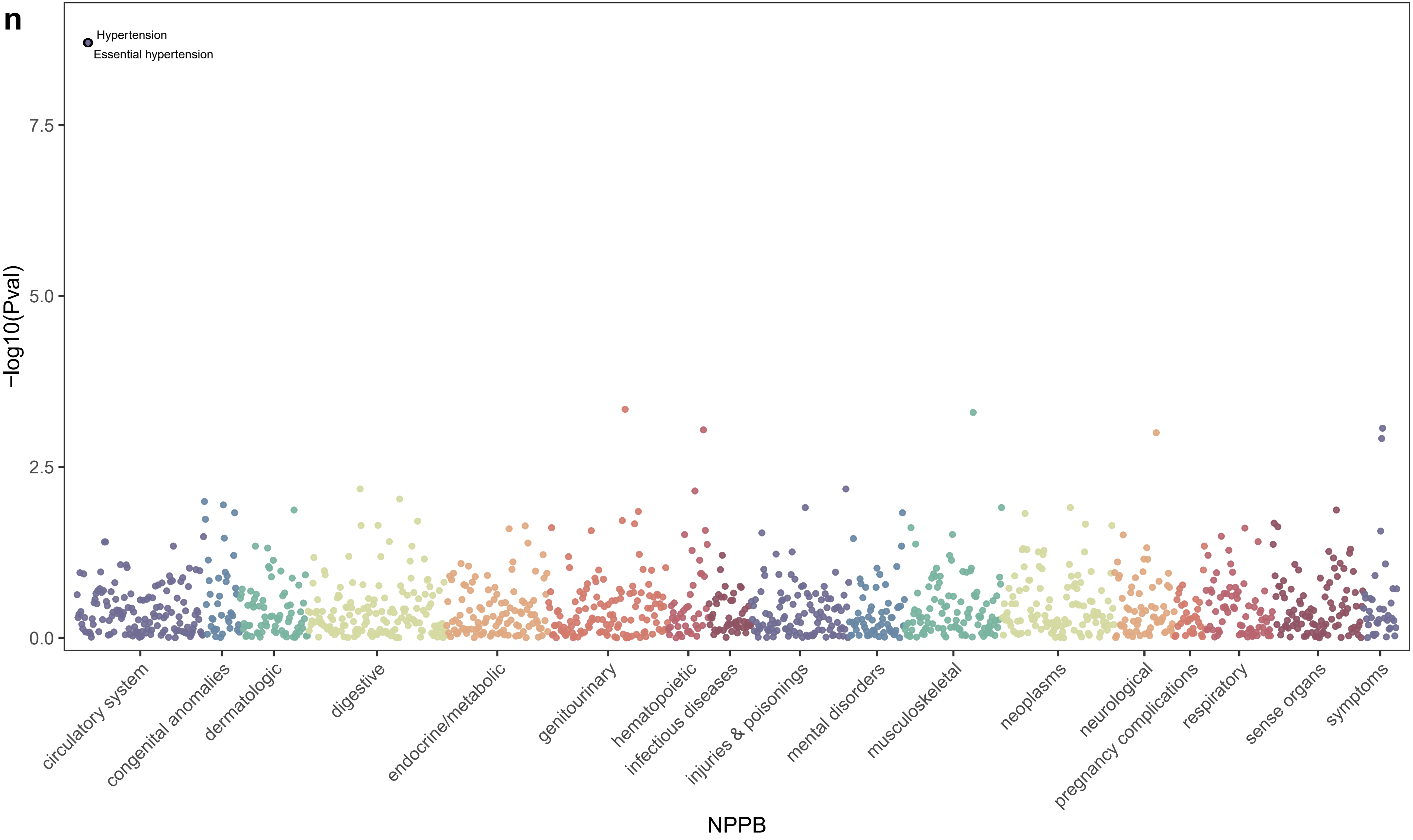
Hypertension
Essential hypertension
7.5
5.0
2.5
0.0
−log10(Pval)
circulatory system
congenital anomalies
dermatologic
digestive
endocrine/metabolic
genitourinary
hematopoietic
infectious diseases
injuries & poisonings
mental disorders
musculoskeletal
neoplasms
neurological
pregnancy complications
respiratory
sense organs
symptoms
NPPB

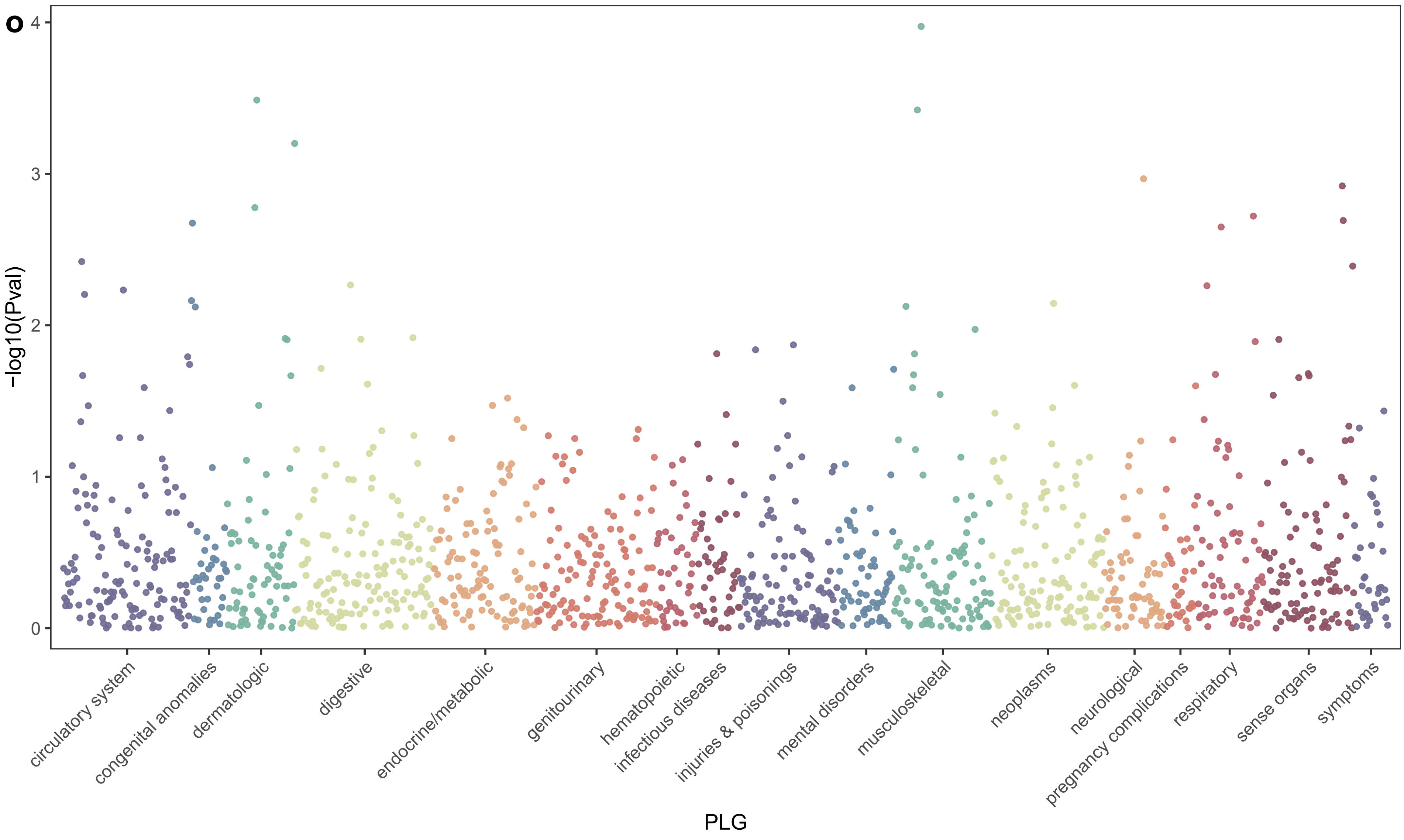
o
-log10(Pval)
0
1
2
3
4
circulatory system
congenital anomalies
dermatologic
digestive
endocrine/metabolic
genitourinary
hematopoietic
infectious diseases
injuries & poisonings
mental disorders
musculoskeletal
neoplasms
neurological
pregnancy complications
respiratory
sense organs
symptoms
PLG

**p**

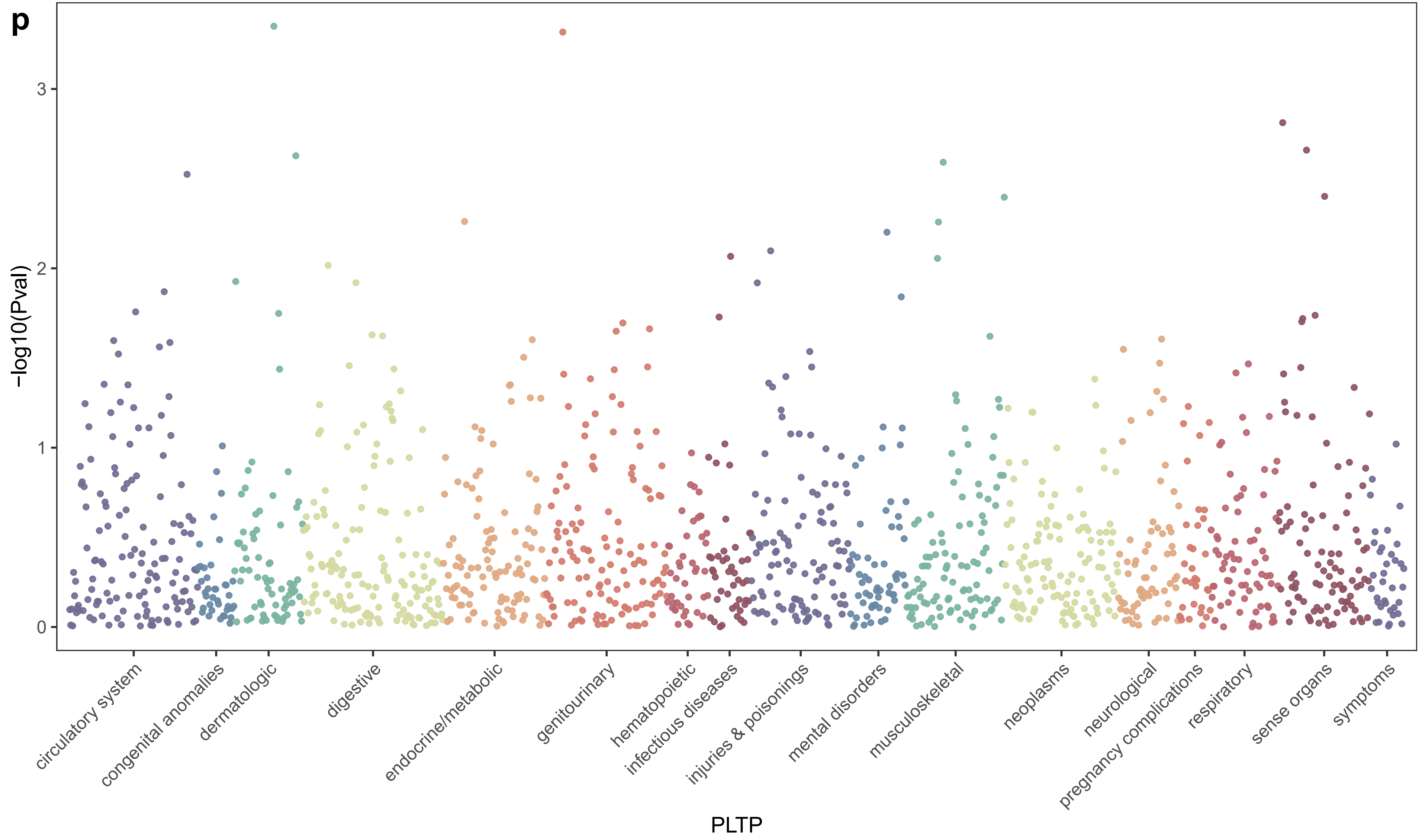

−log10(Pval)
0
1
2
3
circulatory system
congenital anomalies
dermatologic
digestive
endocrine/metabolic
genitourinary
hematopoietic
infectious diseases
injuries & poisonings
mental disorders
musculoskeletal
neoplasms
neurological
pregnancy complications
respiratory
sense organs
symptoms
PLTP

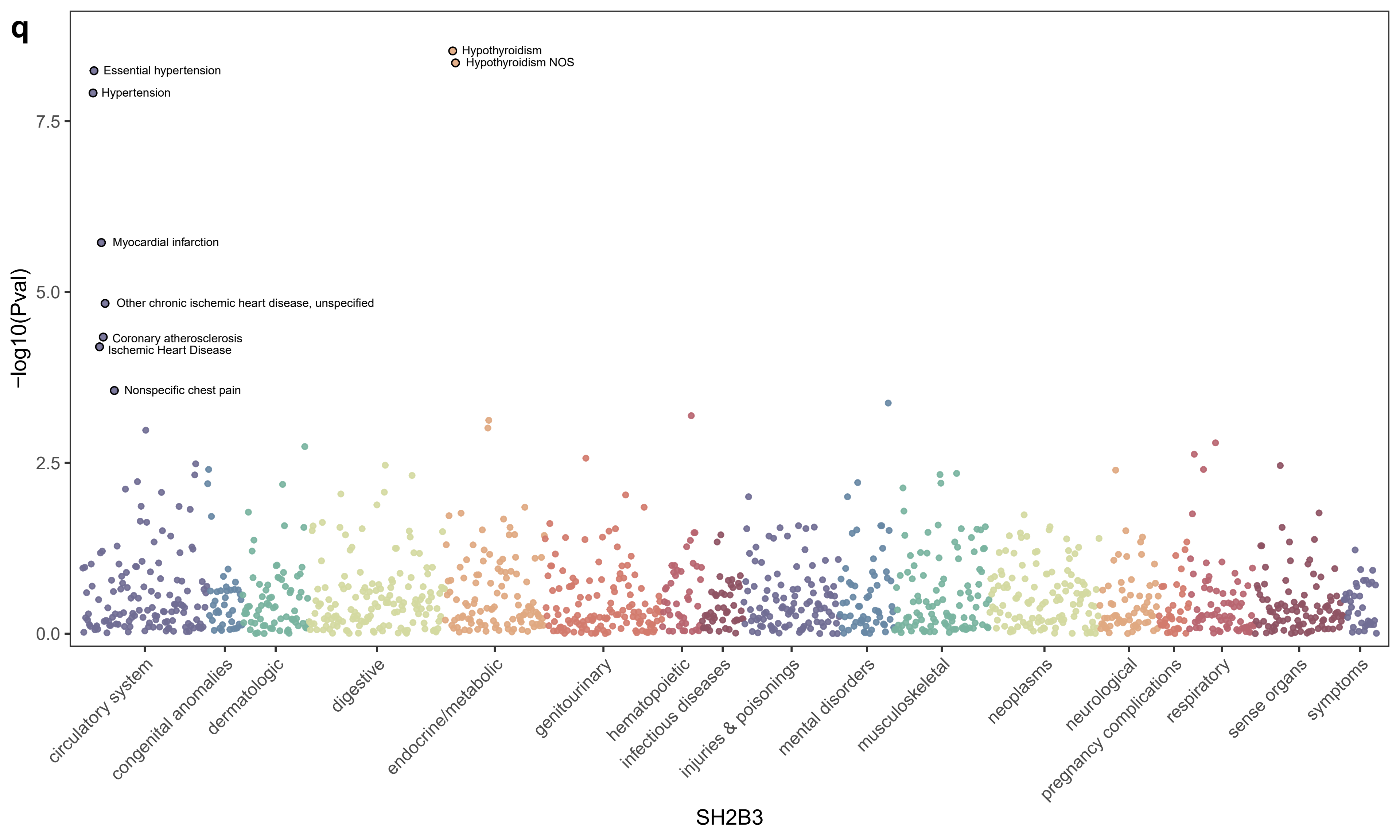
q
Hypothyroidism
Hypothyroidism NOS
Essential hypertension
Hypertension
Myocardial infarction
Other chronic ischemic heart disease, unspecified
Coronary atherosclerosis
Ischemic Heart Disease
Nonspecific chest pain
−log10(Pval)
7.5
5.0
2.5
0.0
circulatory system
congenital anomalies
dermatologic
digestive
endocrine/metabolic
genitourinary
hematopoietic
infectious diseases
injuries & poisonings
mental disorders
musculoskeletal
neoplasms
neurological
pregnancy complications
respiratory
sense organs
symptoms
SH2B3

r

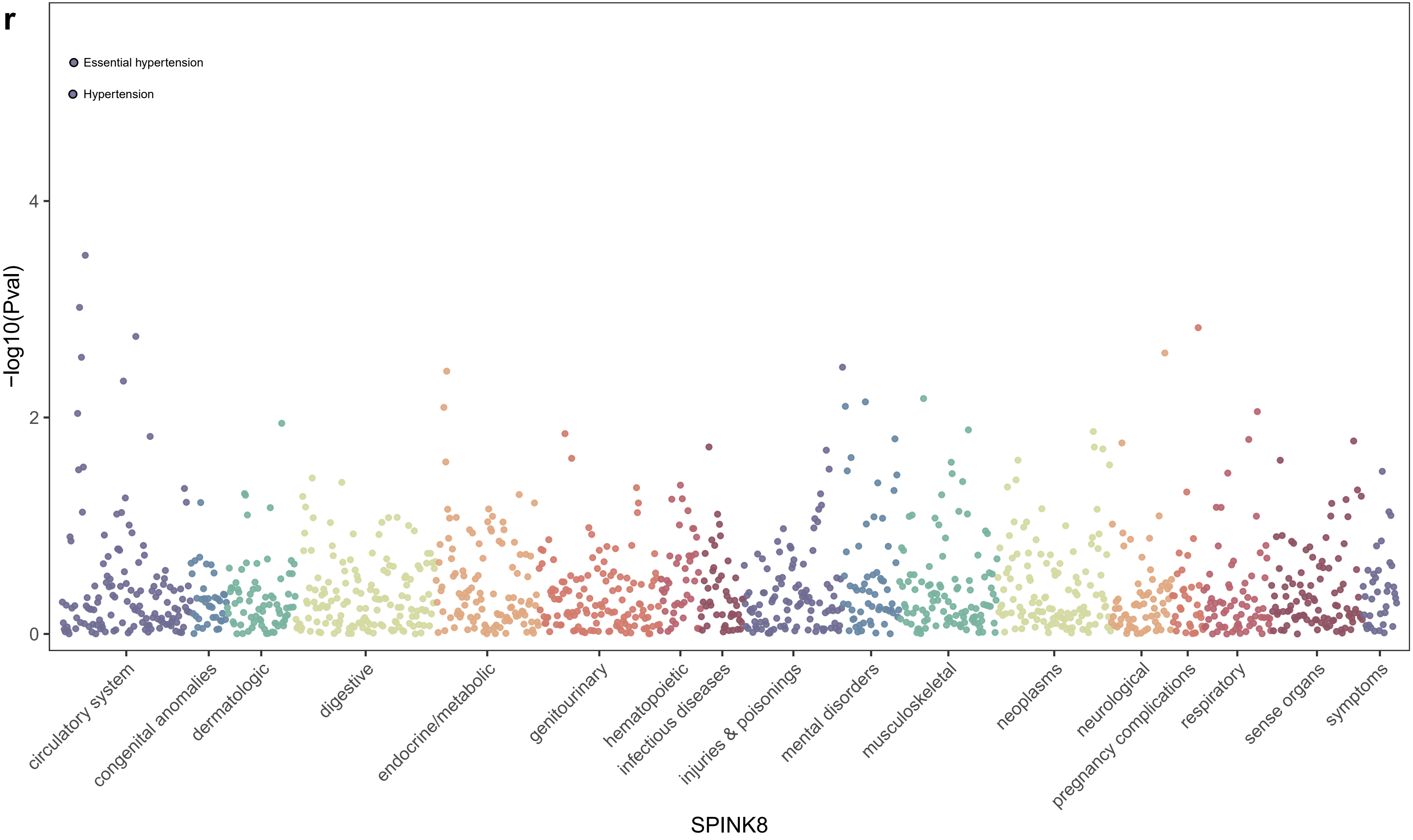
Essential hypertension
Hypertension
-log10(Pval)
0
2
4
circulatory system
congenital anomalies
dermatologic
digestive
endocrine/metabolic
genitourinary
hematopoietic
infectious diseases
injuries & poisonings
mental disorders
musculoskeletal
neoplasms
neurological
pregnancy complications
respiratory
sense organs
symptoms
SPINK8

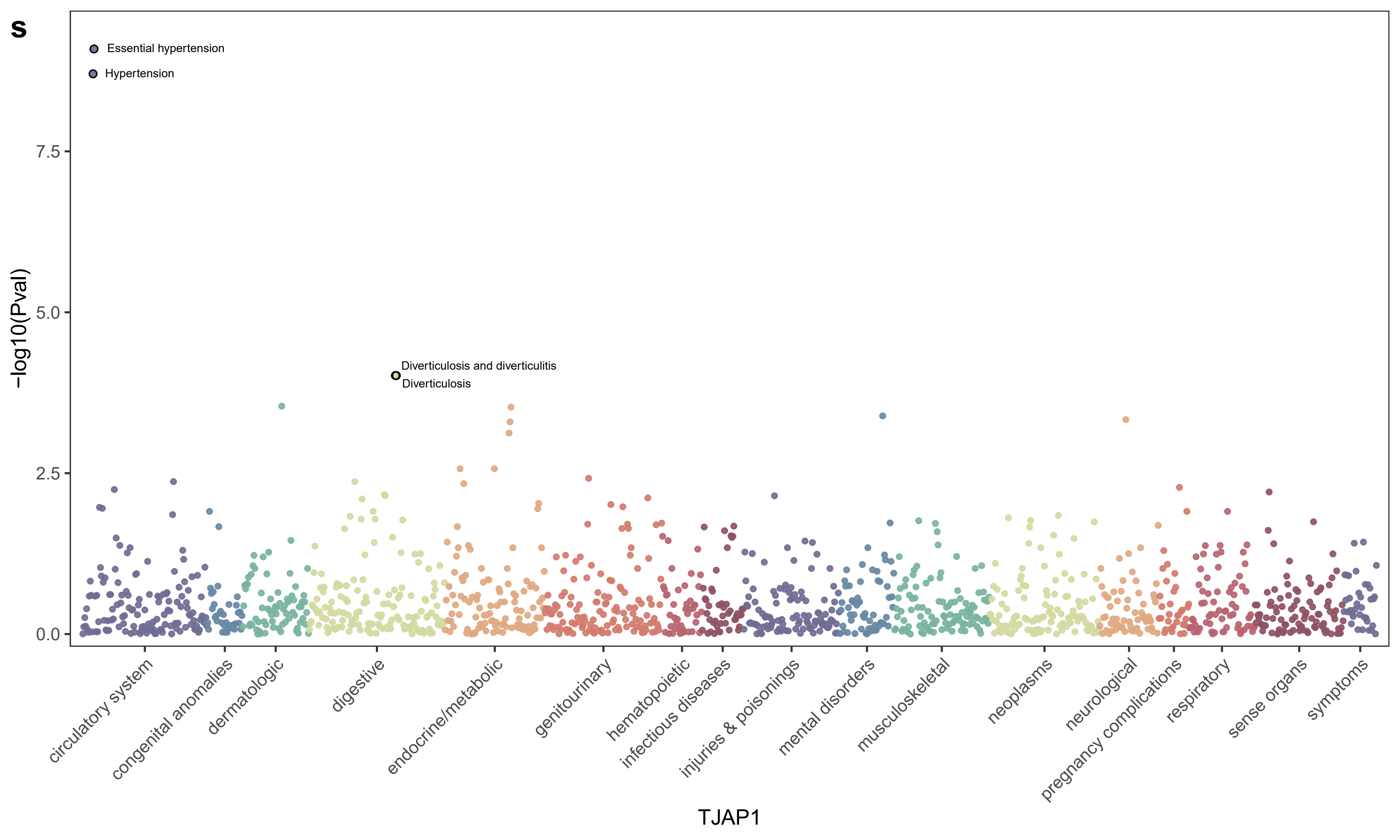
s
Essential hypertension
Hypertension
Diverticulosis and diverticulitis
Diverticulosis
-log10(Pval)
7.5
5.0
2.5
0.0
circulatory system
congenital anomalies
dermatologic
digestive
endocrine/metabolic
genitourinary
hematopoietic
infectious diseases
injuries & poisonings
mental disorders
musculoskeletal
neoplasms
neurological
pregnancy complications
respiratory
sense organs
symptoms
TJAP1

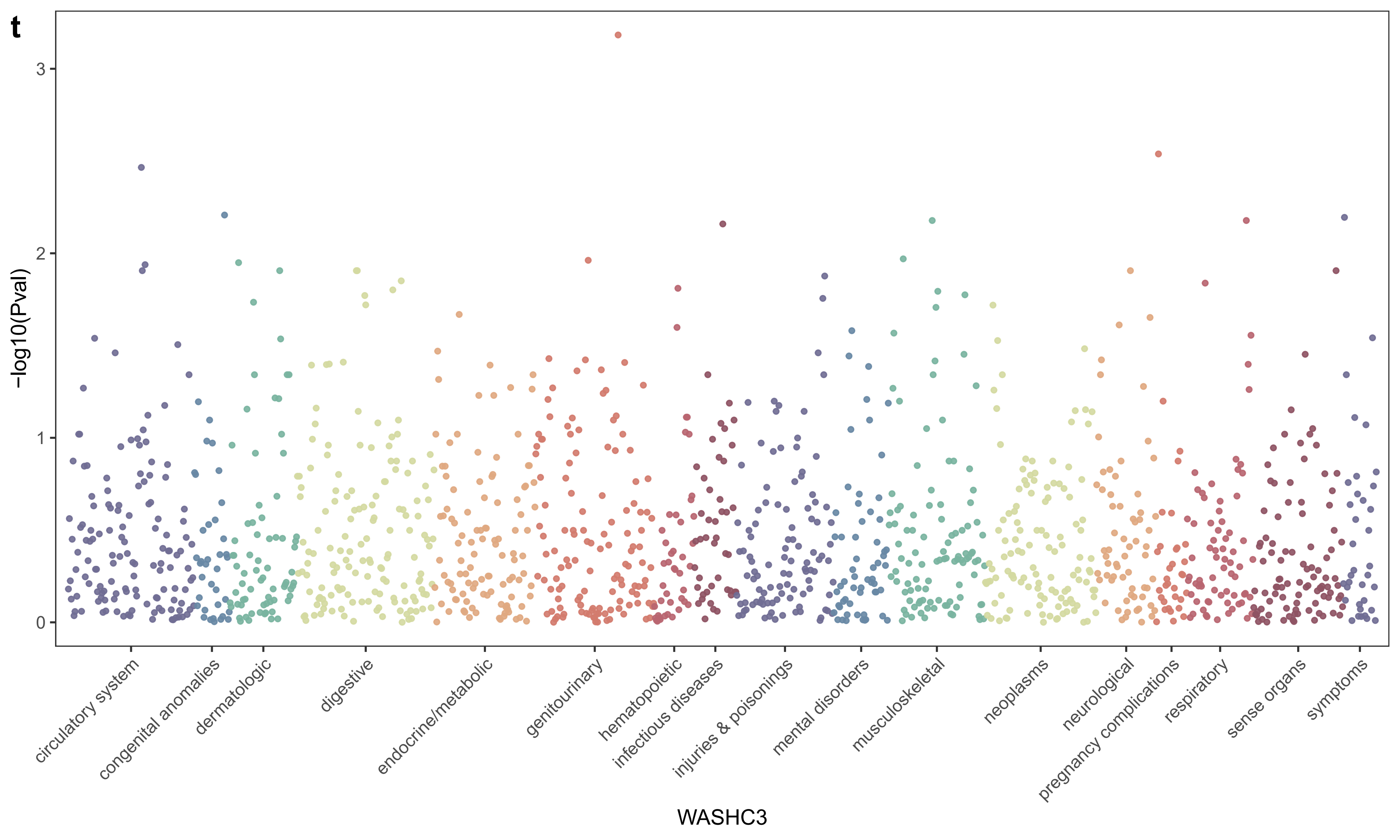
t
-log10(Pval)
0
1
2
3
circulatory system
congenital anomalies
dermatologic
digestive
endocrine/metabolic
genitourinary
hematopoietic
infectious diseases
injuries & poisonings
mental disorders
musculoskeletal
neoplasms
neurological
pregnancy complications
respiratory
sense organs
symptoms
WASHC3

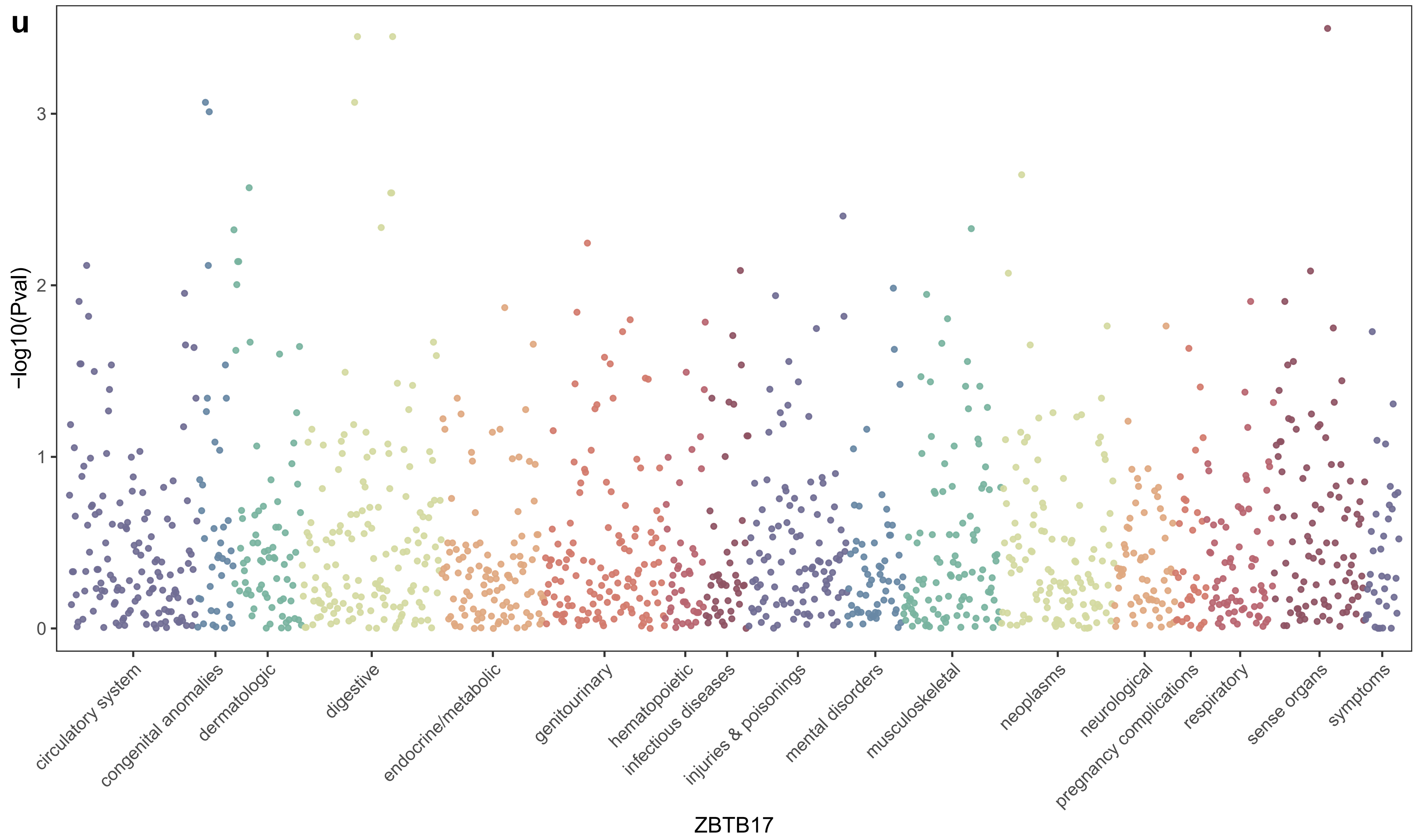

u
−log10(Pval)
0
1
2
3
circulatory system
congenital anomalies
dermatologic
digestive
endocrine/metabolic
genitourinary
hematopoietic
infectious diseases
injuries & poisonings
mental disorders
musculoskeletal
neoplasms
neurological
pregnancy complications
respiratory
sense organs
symptoms
ZBTB17
