## Supplementary Figure 7 for "Exploring novel risk loci for heart failure and the shared genetic etiology with blood lipids, blood pressure, and blood glucose: a large-scale multi-trait association analysis"

a

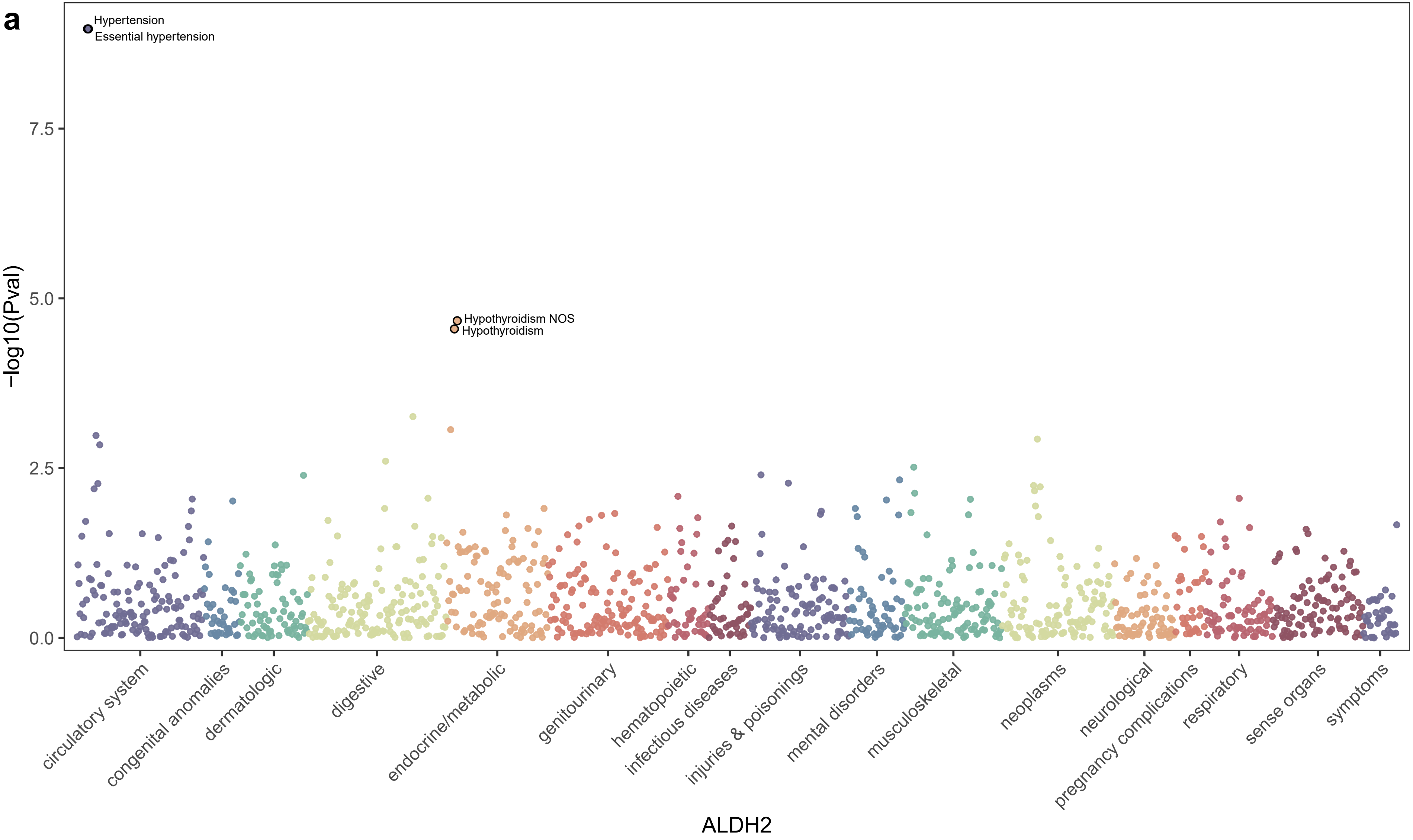

Hypertension
Essential hypertension
Hypothyroidism NOS
Hypothyroidism
7.5
5.0
2.5
0.0
-log10(Pval)
circulatory system
congenital anomalies
dermatologic
digestive
endocrine/metabolic
genitourinary
hematopoietic
infectious diseases
injuries & poisonings
mental disorders
musculoskeletal
neoplasms
neurological
pregnancy complications
respiratory
sense organs
symptoms
ALDH2

b

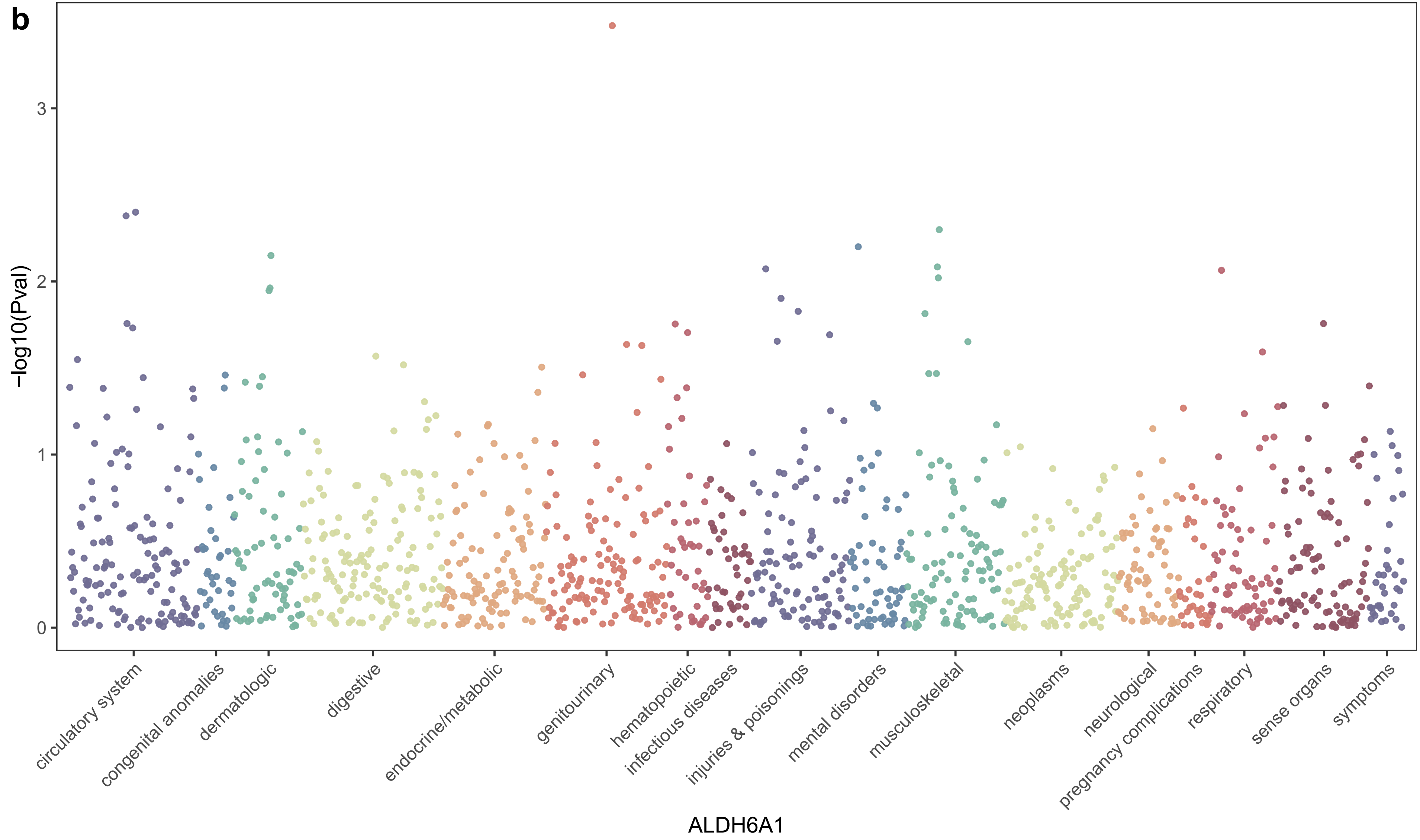

-log10(Pval)
3
2
1
0
circulatory system
congenital anomalies
dermatologic
digestive
endocrine/metabolic
genitourinary
hematopoietic
infectious diseases
injuries & poisonings
mental disorders
musculoskeletal
neoplasms
neurological
pregnancy complications
respiratory
sense organs
symptoms
ALDH6A1

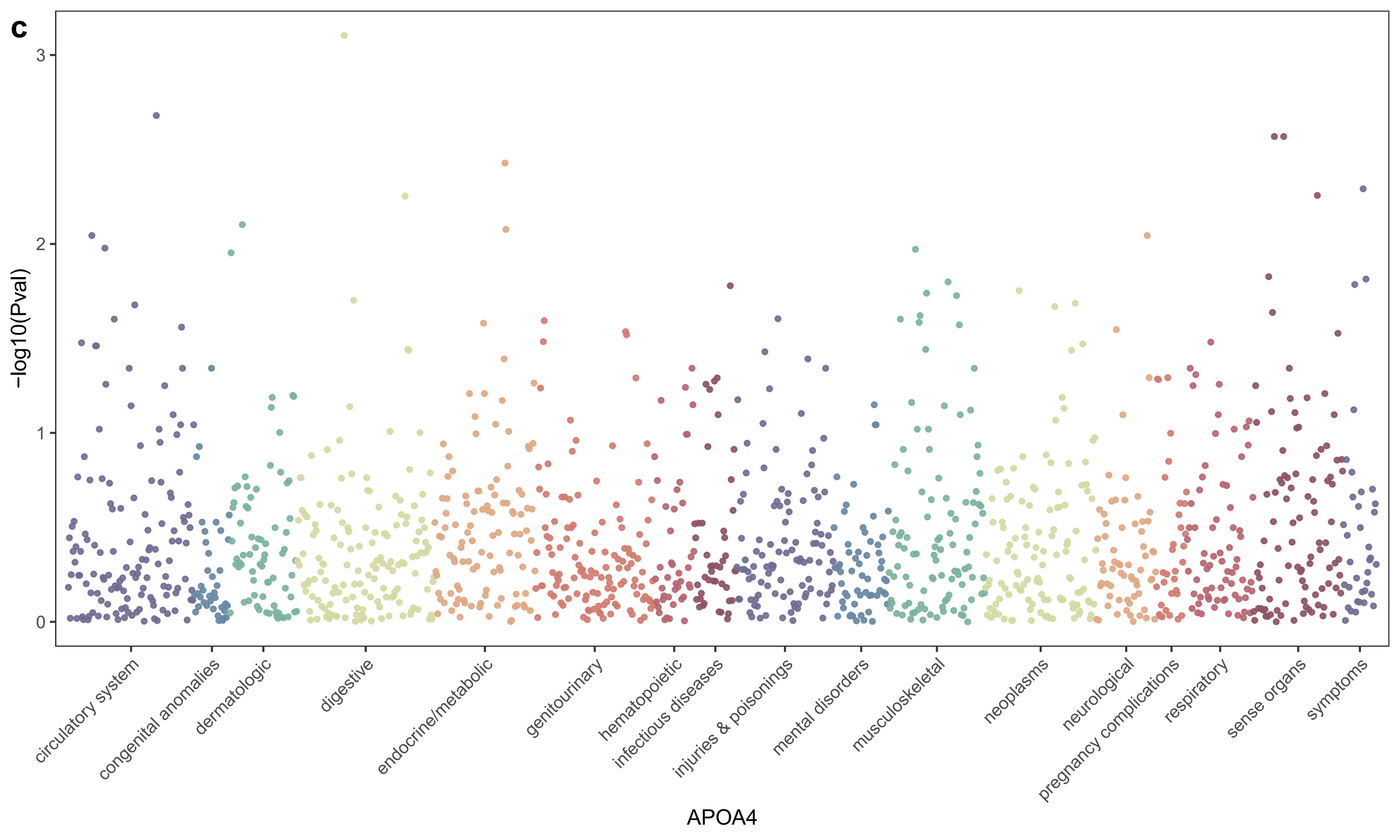
c
3
2
1
0
-log10(Pval)
circulatory system
congenital anomalies
dermatologic
digestive
endocrine/metabolic
genitourinary
hematopoietic
infectious diseases
injuries & poisonings
mental disorders
musculoskeletal
neoplasms
neurological
pregnancy complications
respiratory
sense organs
symptoms
APOA4

**d**

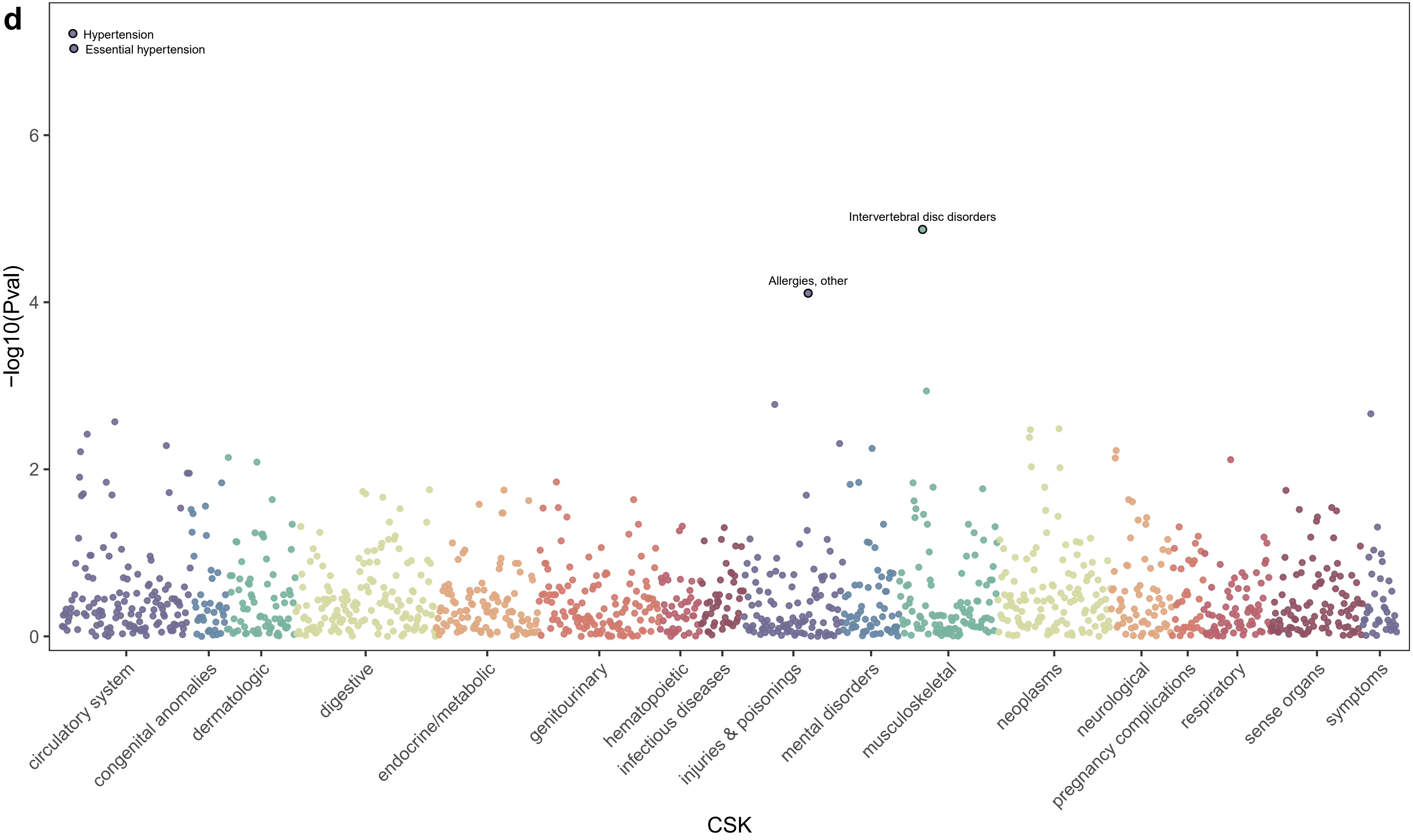
Hypertension
Essential hypertension
Intervertebral disc disorders
Allergies, other
−log10(Pval)
0
2
4
6
circulatory system
congenital anomalies
dermatologic
digestive
endocrine/metabolic
genitourinary
hematopoietic
infectious diseases
injuries & poisonings
mental disorders
musculoskeletal
neoplasms
neurological
pregnancy complications
respiratory
sense organs
symptoms
CSK

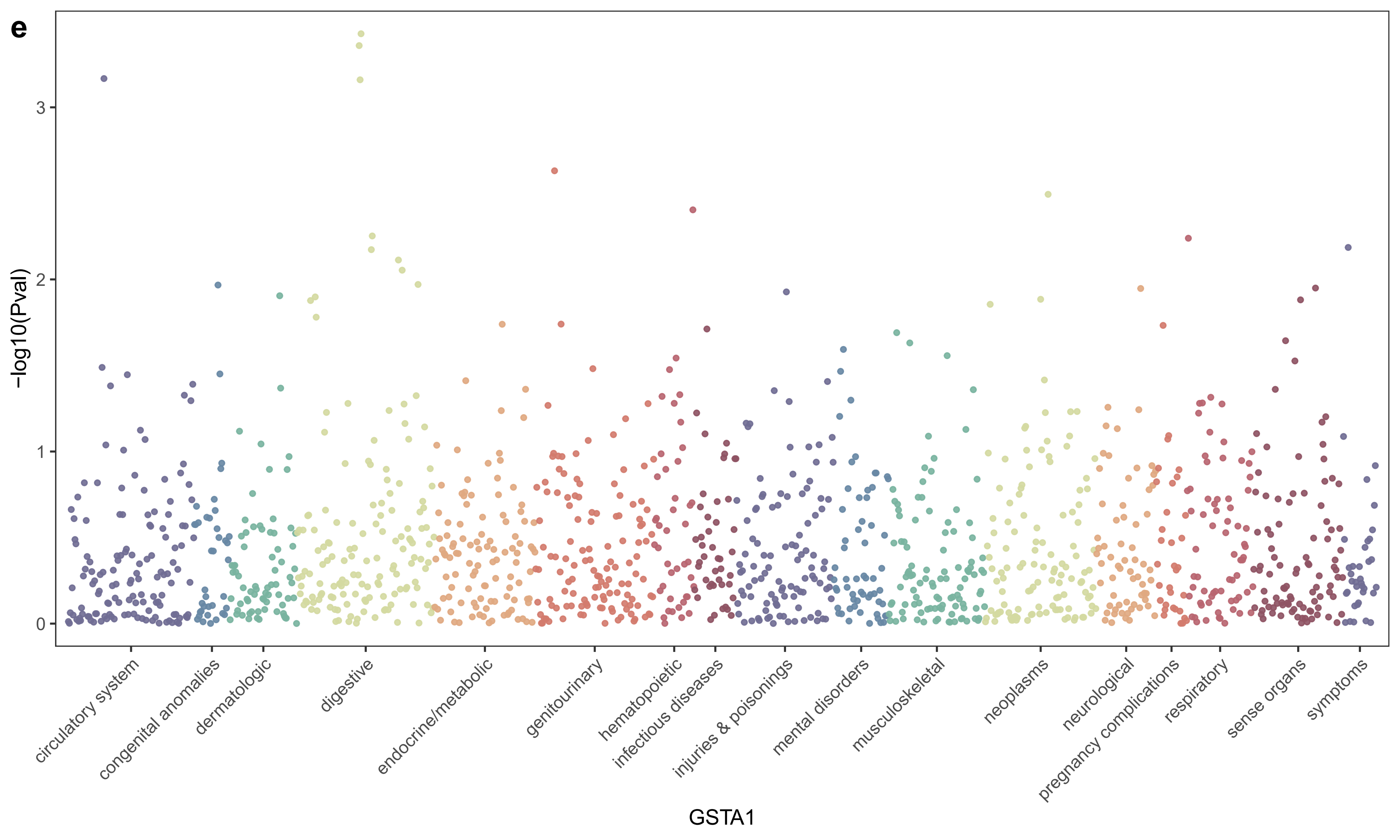
e
-log10(Pval)
0
1
2
3
circulatory system
congenital anomalies
dermatologic
digestive
endocrine/metabolic
genitourinary
hematopoietic
infectious diseases
injuries & poisonings
mental disorders
musculoskeletal
neoplasms
neurological
pregnancy complications
respiratory
sense organs
symptoms
GSTA1

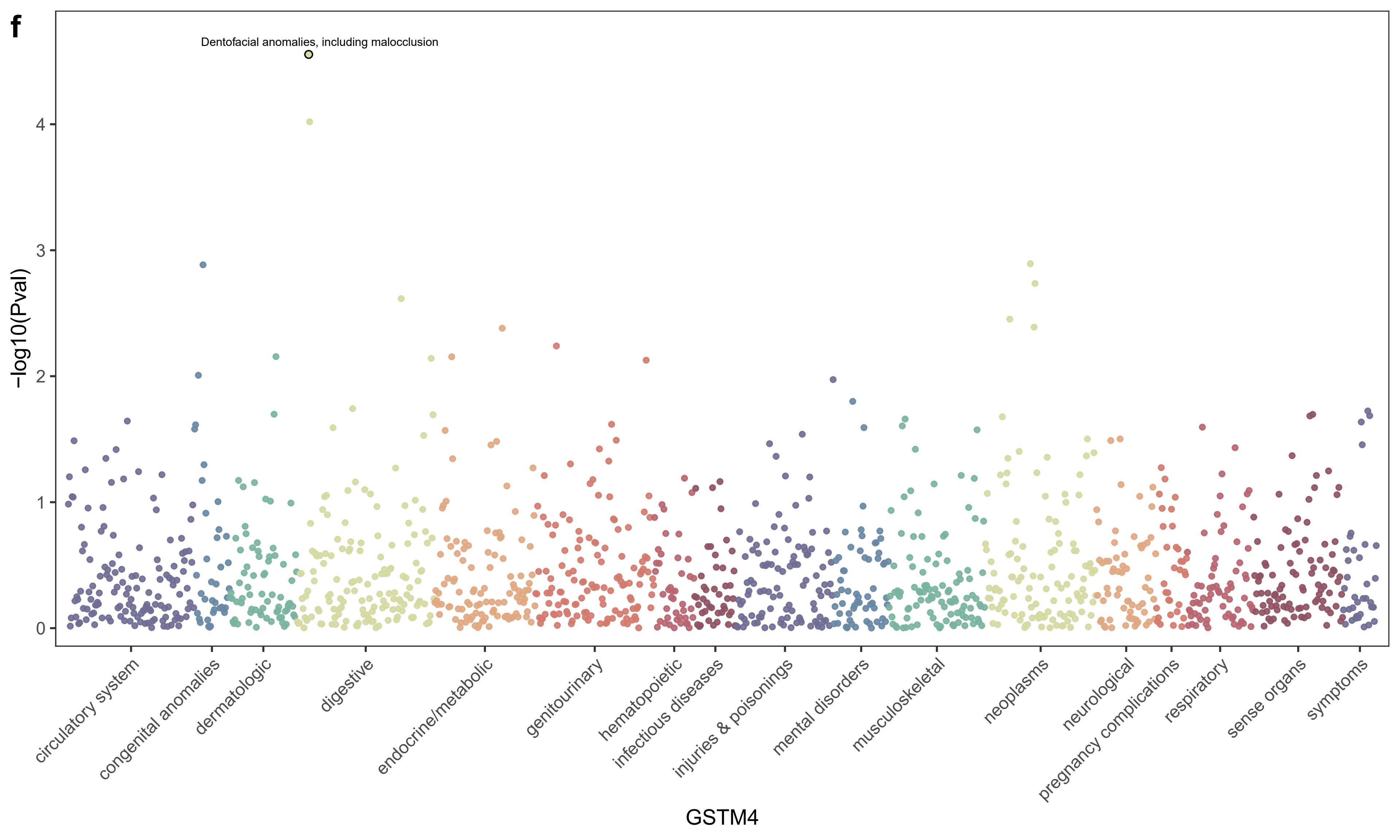
f
Dentofacial anomalies, including malocclusion
-log10(Pval)
0
1
2
3
4
circulatory system
congenital anomalies
dermatologic
digestive
endocrine/metabolic
genitourinary
hematopoietic
infectious diseases
injuries & poisonings
mental disorders
musculoskeletal
neoplasms
neurological
pregnancy complications
respiratory
sense organs
symptoms
GSTM4

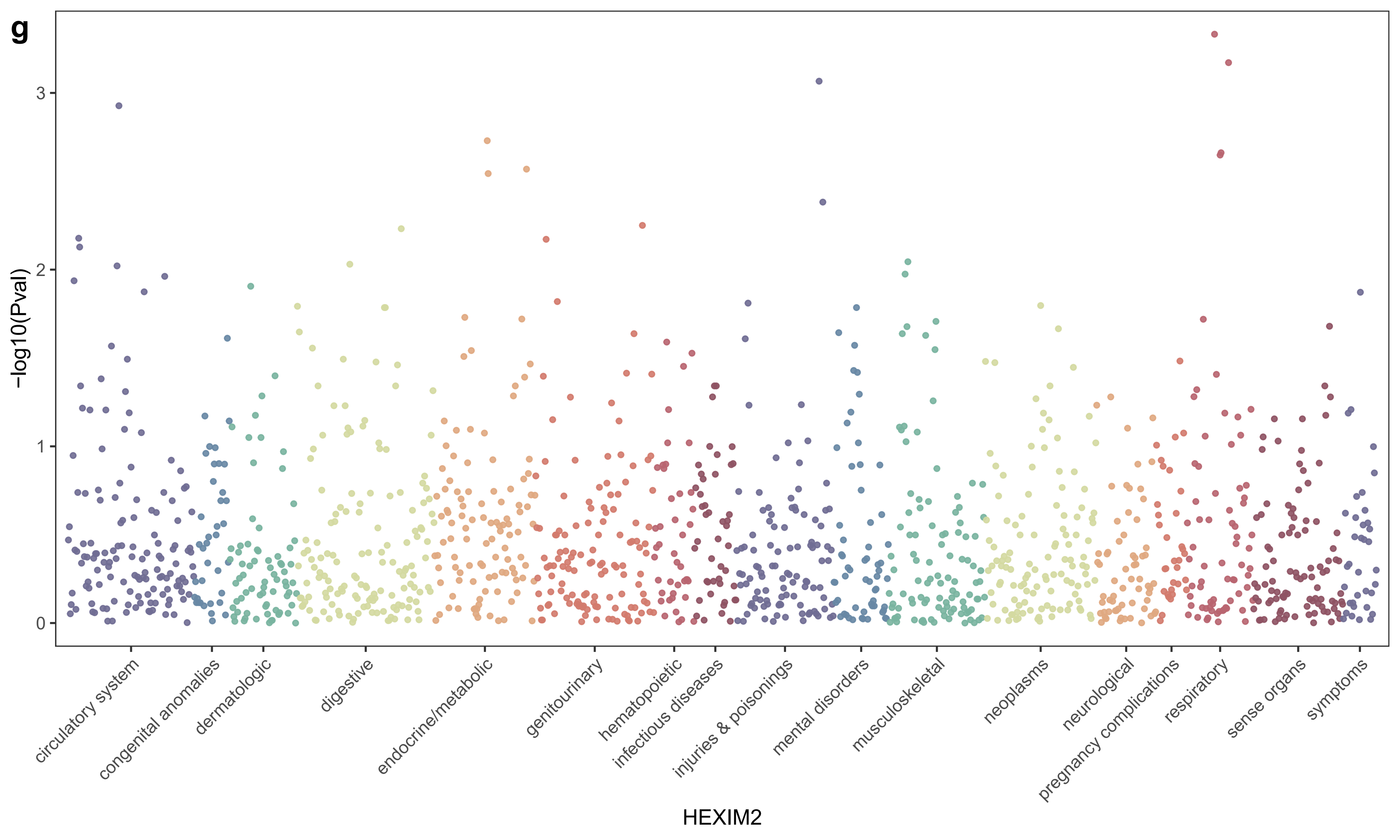
g
-log10(Pval)
0
1
2
3
circulatory system
congenital anomalies
dermatologic
digestive
endocrine/metabolic
genitourinary
hematopoietic
infectious diseases
injuries & poisonings
mental disorders
musculoskeletal
neoplasms
neurological
pregnancy complications
respiratory
sense organs
symptoms
HEXIM2

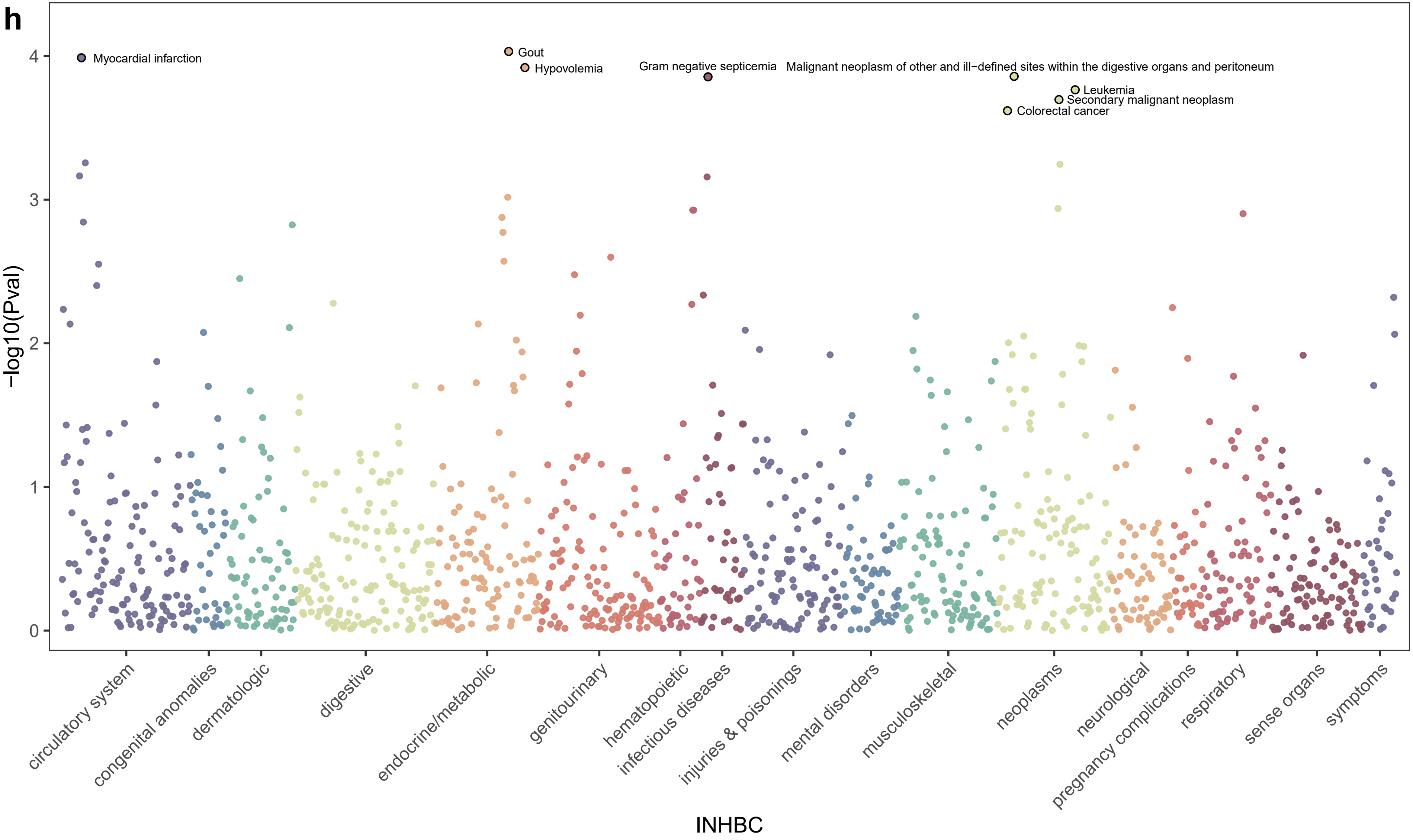
h
Myocardial infarction
Gout
Hypovolemia
Gram negative septicemia
Malignant neoplasm of other and ill−defined sites within the digestive organs and peritoneum
Leukemia
Secondary malignant neoplasm
Colorectal cancer
−log10(Pval)
0
1
2
3
4
circulatory system
congenital anomalies
dermatologic
digestive
endocrine/metabolic
genitourinary
hematopoietic
infectious diseases
injuries & poisonings
mental disorders
musculoskeletal
neoplasms
neurological
pregnancy complications
respiratory
sense organs
symptoms
INHBC

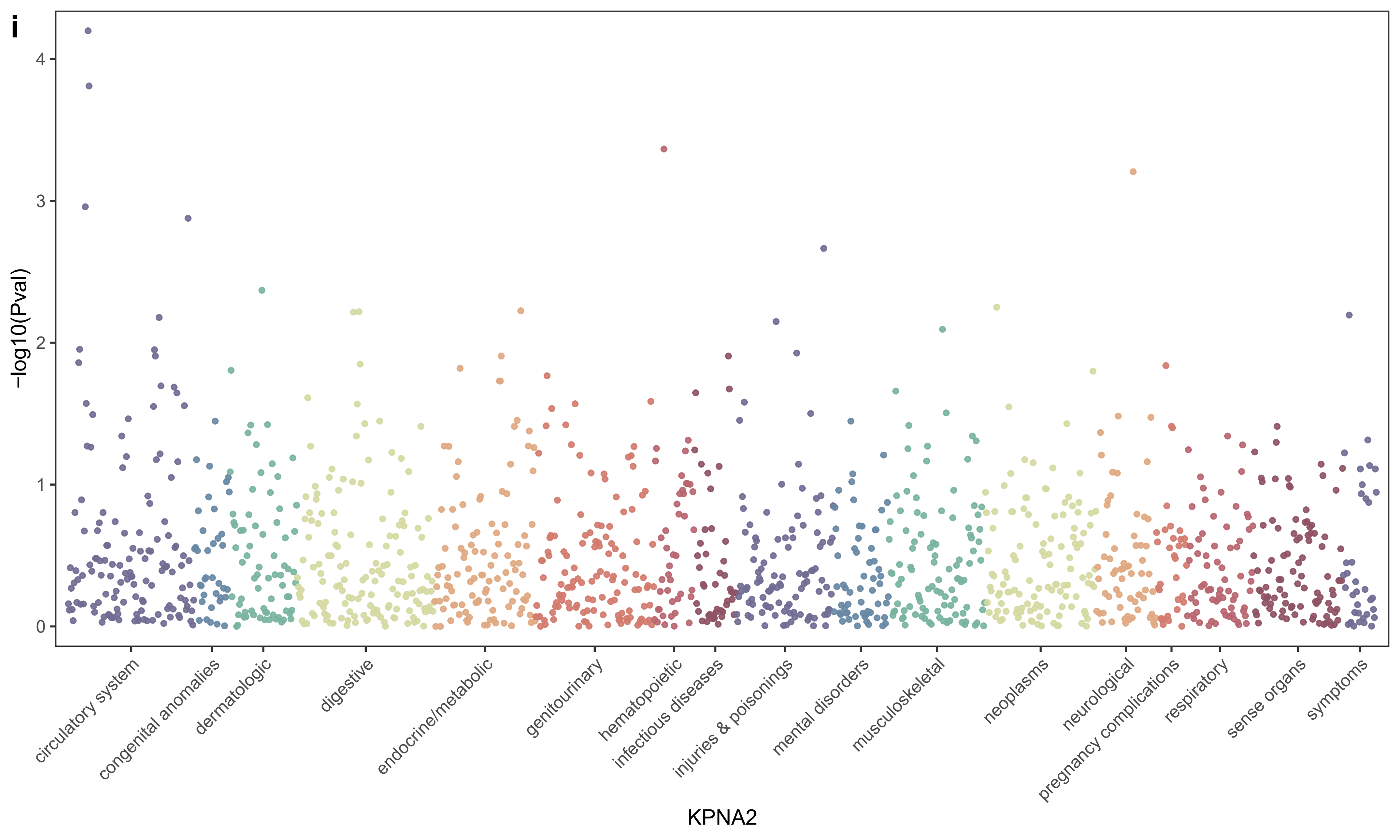
i
-log10(Pval)
0
1
2
3
4
circulatory system
congenital anomalies
dermatologic
digestive
endocrine/metabolic
genitourinary
hematopoietic
infectious diseases
injuries & poisonings
mental disorders
musculoskeletal
neoplasms
neurological
pregnancy complications
respiratory
sense organs
symptoms
KPNA2

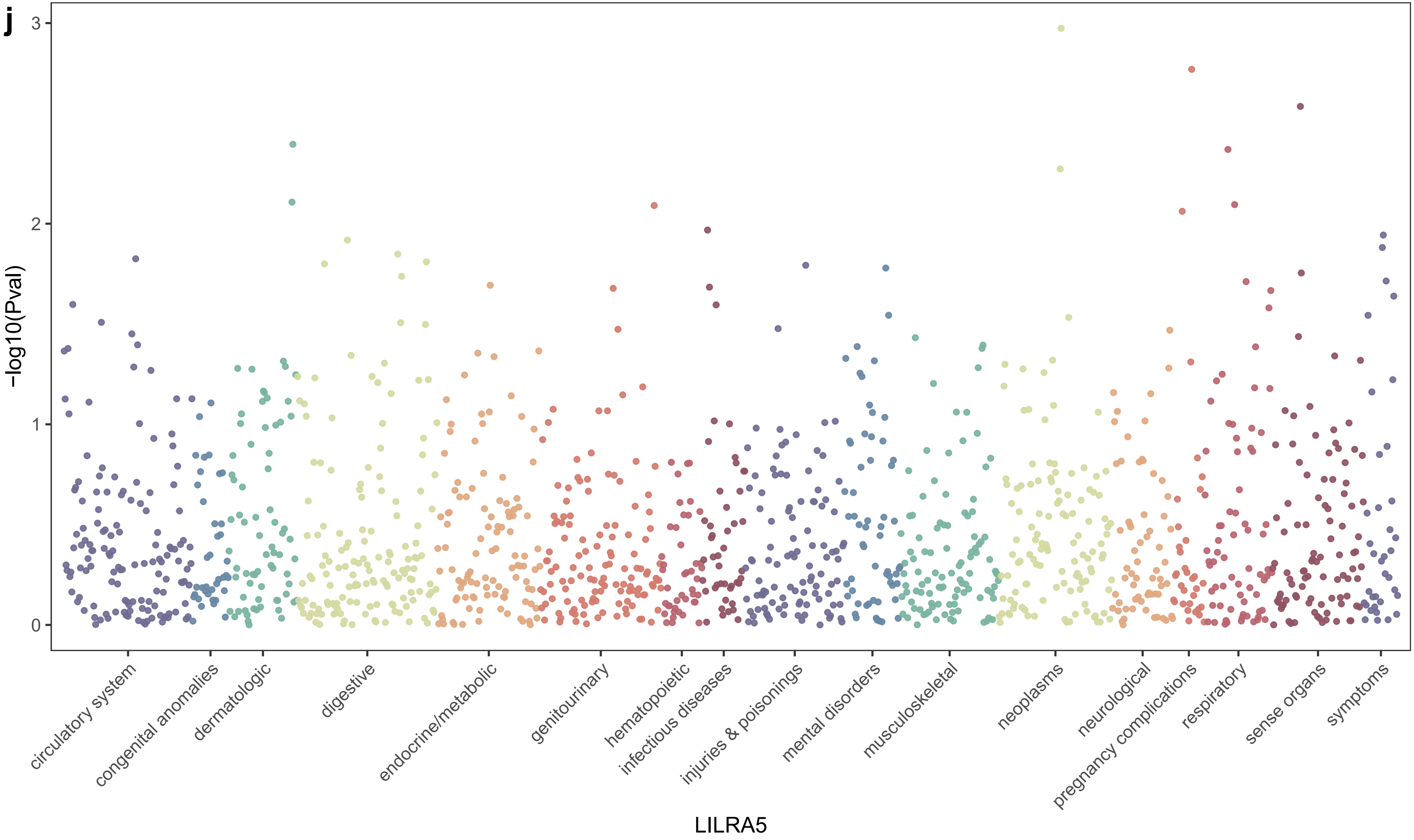
j
−log10(Pval)
0
1
2
3
circulatory system
congenital anomalies
dermatologic
digestive
endocrine/metabolic
genitourinary
hematopoietic
infectious diseases
injuries & poisonings
mental disorders
musculoskeletal
neoplasms
neurological
pregnancy complications
respiratory
sense organs
symptoms
LILRA5

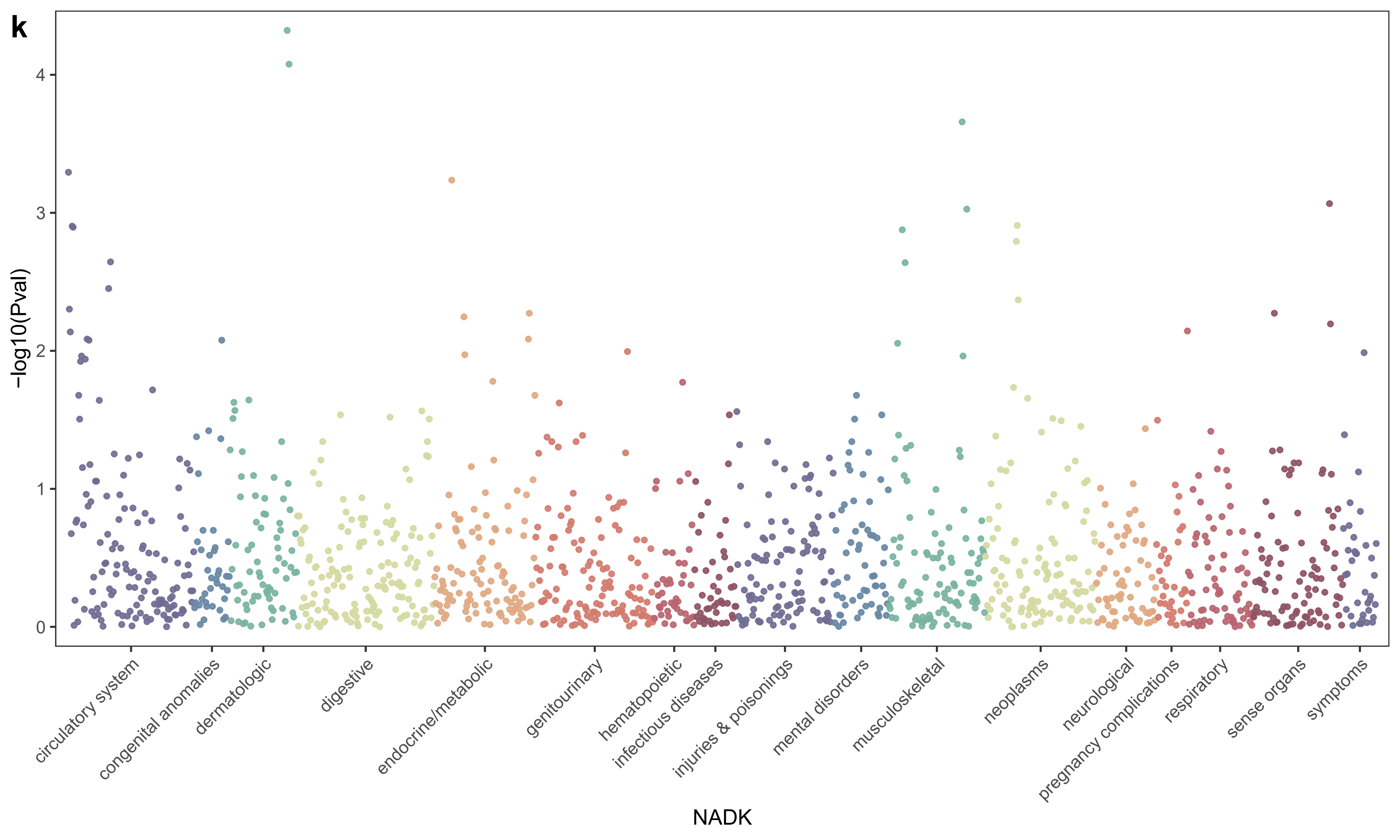
k
−log10(Pval)
4
3
2
1
0
circulatory system
congenital anomalies
dermatologic
digestive
endocrine/metabolic
genitourinary
hematopoietic
infectious diseases
injuries & poisonings
mental disorders
musculoskeletal
neoplasms
neurological
pregnancy complications
respiratory
sense organs
symptoms
NADK

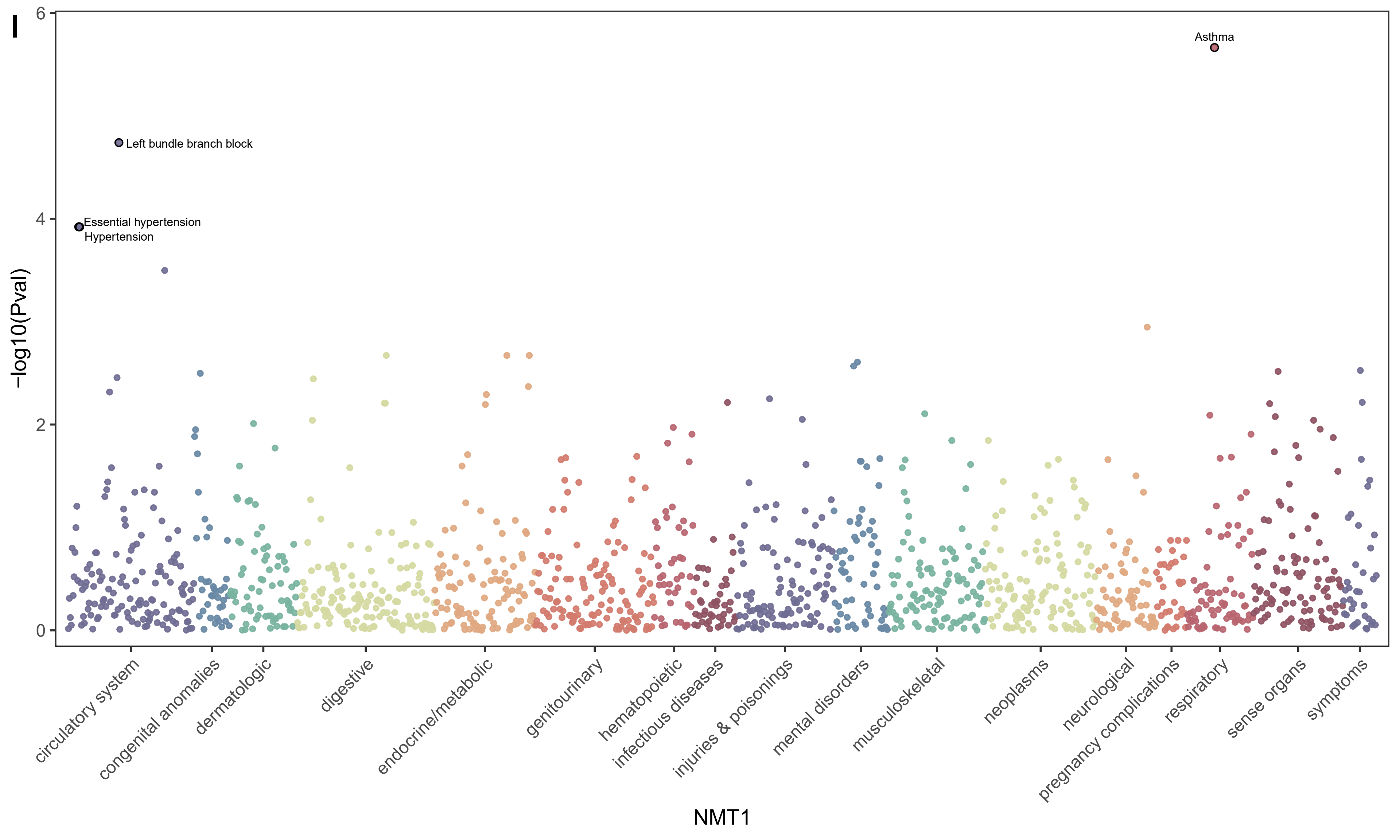
I
Asthma
Left bundle branch block
Essential hypertension
Hypertension
-log10(Pval)
6
4
2
0
circulatory system
congenital anomalies
dermatologic
digestive
endocrine/metabolic
genitourinary
hematopoietic
infectious diseases
injuries & poisonings
mental disorders
musculoskeletal
neoplasms
neurological
pregnancy complications
respiratory
sense organs
symptoms
NMT1

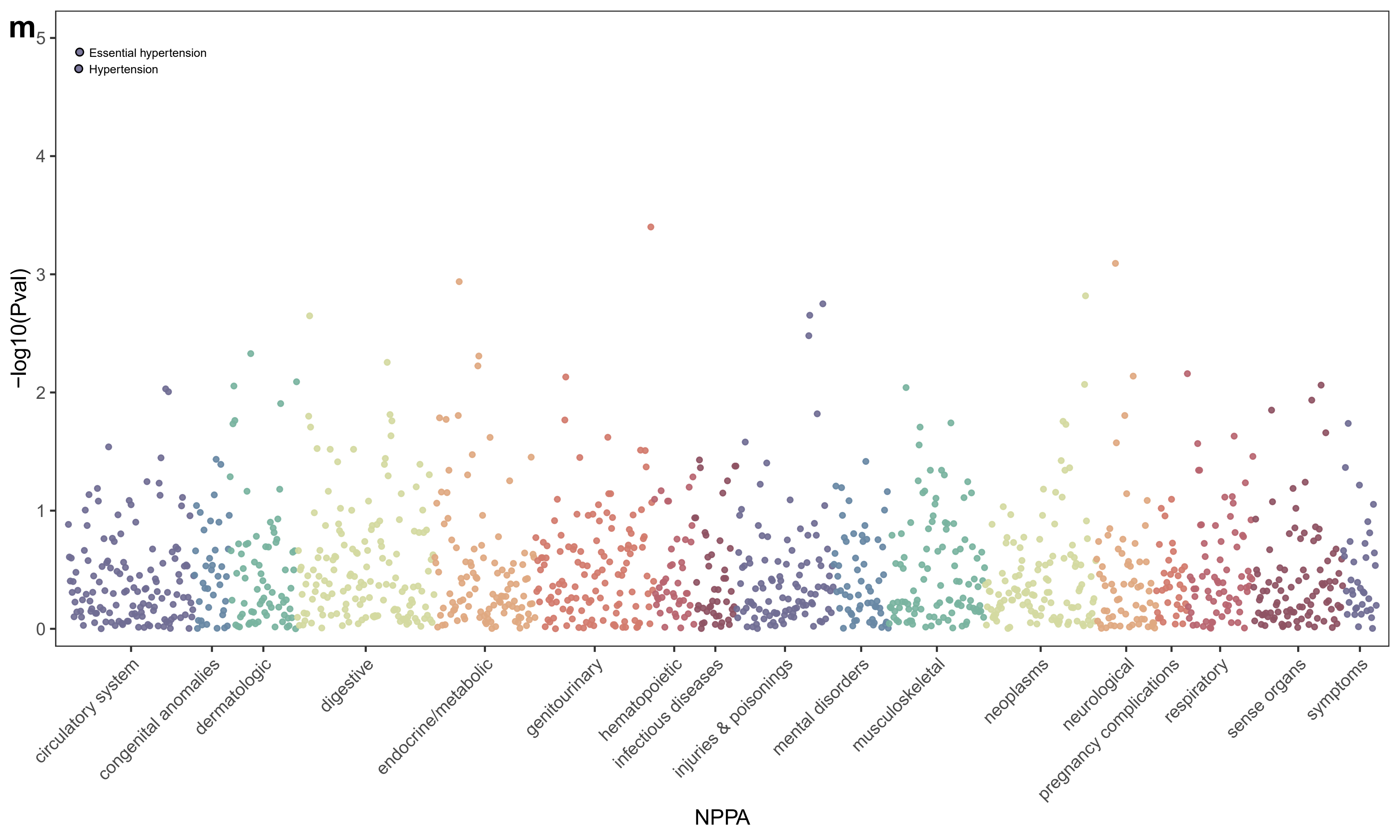

m
Essential hypertension
Hypertension
-log10(Pval)
0
1
2
3
4
5
circulatory system
congenital anomalies
dermatologic
digestive
endocrine/metabolic
genitourinary
hematopoietic
infectious diseases
injuries & poisonings
mental disorders
musculoskeletal
neoplasms
neurological
pregnancy complications
respiratory
sense organs
symptoms
NPPA

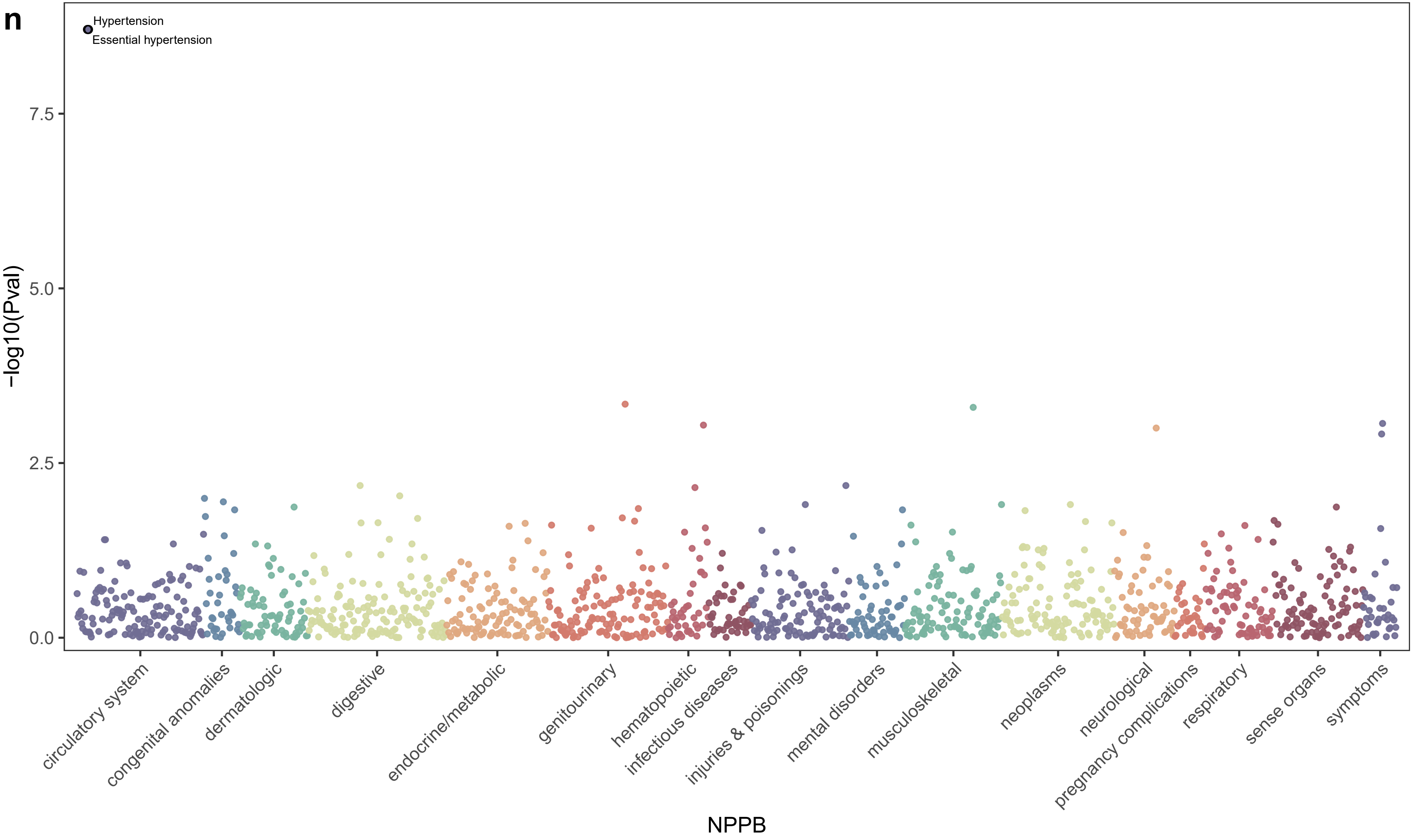

n
Hypertension
Essential hypertension
7.5
5.0
2.5
0.0
-log10(Pval)
circulatory system
congenital anomalies
dermatologic
digestive
endocrine/metabolic
genitourinary
hematopoietic
infectious diseases
injuries & poisonings
mental disorders
musculoskeletal
neoplasms
neurological
pregnancy complications
respiratory
sense organs
symptoms
NPPB

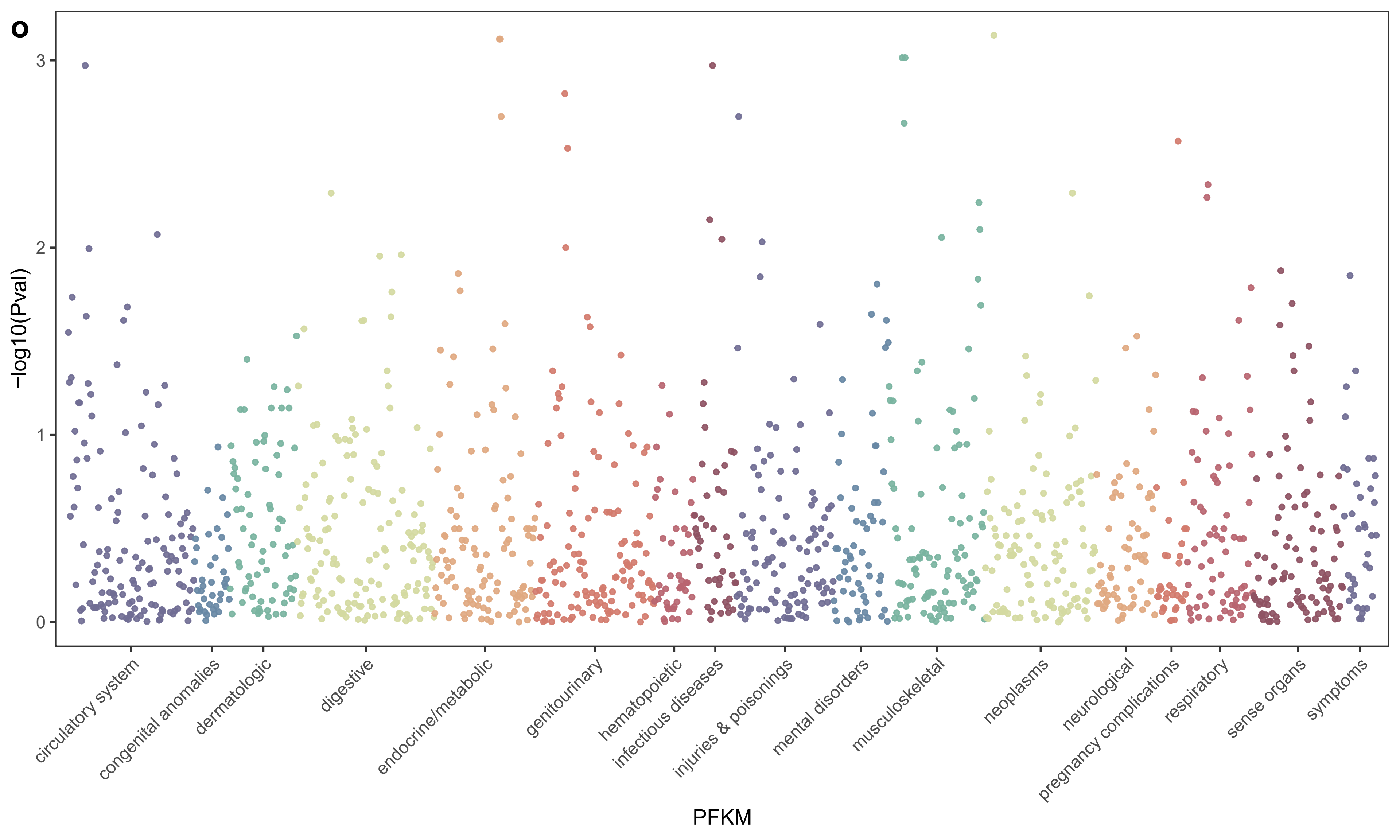
o
-log10(Pval)
3
2
1
0
circulatory system
congenital anomalies
dermatologic
digestive
endocrine/metabolic
genitourinary
hematopoietic
infectious diseases
injuries & poisonings
mental disorders
musculoskeletal
neoplasms
neurological
pregnancy complications
respiratory
sense organs
symptoms
PFKM

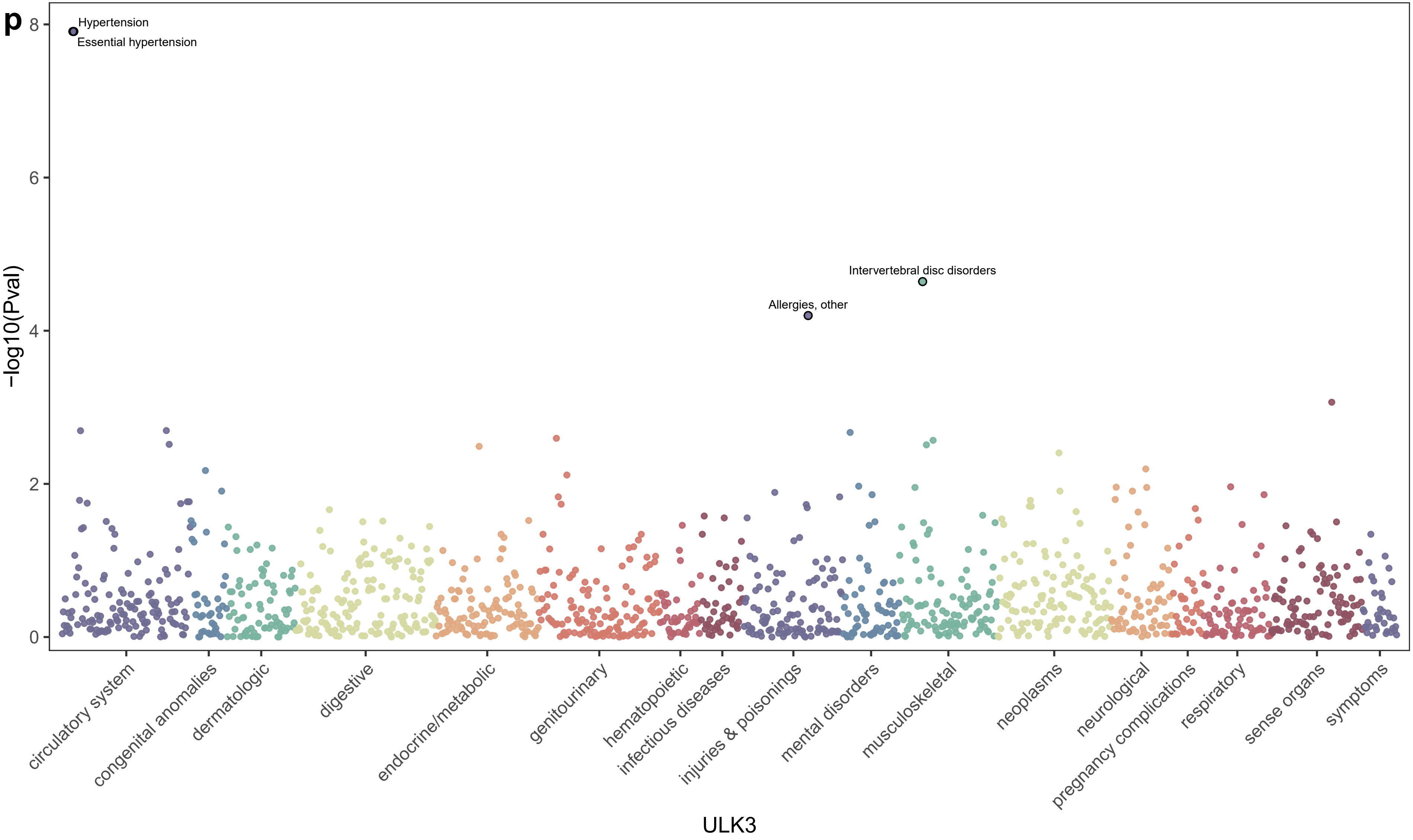
p
Hypertension
Essential hypertension
Intervertebral disc disorders
Allergies, other
-log10(Pval)
0
2
4
6
8
circulatory system
congenital anomalies
dermatologic
digestive
endocrine/metabolic
genitourinary
hematopoietic
infectious diseases
injuries & poisonings
mental disorders
musculoskeletal
neoplasms
neurological
pregnancy complications
respiratory
sense organs
symptoms
ULK3
